# Diagnostic accuracy of rapid point-of-care tests for diagnosis of current SARS-CoV-2 infections in children: A systematic review and meta-analysis

**DOI:** 10.1101/2021.08.11.21261830

**Authors:** Naomi Fujita-Rohwerder, Lars Beckmann, Yvonne Zens, Arpana Verma

**Author notes:** **Correspondence to:** Dr Naomi Fujita-Rohwerder, Institute for Quality and Efficiency in Health Care (IQWiG), Im Mediapark 8, 50670 Cologne, Germany. **Copyright statement:** The Corresponding Author has the right to grant on behalf of all authors and does grant on behalf of all authors, a worldwide licence to the Publishers and its licensees in perpetuity, in all forms, formats and media (whether known now or created in the future), to i) publish, reproduce, distribute, display and store the Contribution, ii) translate the Contribution into other languages, create adaptations, reprints, include within collections and create summaries, extracts and/or, abstracts of the Contribution, iii) create any other derivative work(s) based on the Contribution, iv) to exploit all subsidiary rights in the Contribution, v) the inclusion of electronic links from the Contribution to third party material where-ever it may be located; and, vi) licence any third party to do any or all of the above. **Transparency declaration / Guarantor information:** The lead author (NFR) is the manuscript’s guarantor and affirms that the manuscript is an honest, accurate, and transparent account of the study being reported; that no important aspects of the study have been omitted; and that any discrepancies from the study as planned and registered have been explained. The corresponding author attests that all listed authors meet authorship criteria and that no others meeting the criteria have been omitted. **Authors’ contributions:** NFR: idea/conceptualisation, protocol, literature search (search strategies, screening), data-extraction, quality assessment, data-analysis, manuscript draft [lead author], manuscript review; LB: protocol, data extraction, quality assessment, data-analysis, manuscript draft [contributor], manuscript review; YZ: protocol, literature search (screening), manuscript review; AV: supervision, protocol, manuscript review. All authors read and approved the final manuscript. **Competing interest statement:** All authors have completed the ICMJE disclosure uniform (available on request from the corresponding author) and declare: no support from any organisation for the submitted work; no financial relationships with any organisations that might have an interest in the submitted work in the previous three years; no other relationships or activities that could appear to have influenced the submitted work. **Ethical approval:** Approval of an institutional review board or ethics committee was not required for this systematic review. **Details of funding:** No funding was received for conducting this systematic review. **Details of the role of the study sponsors:** Not applicable. **Statement of independence of researchers from funders:** Not applicable.

## Abstract

**Objective:** To systematically assess the diagnostic accuracy of rapid point-of-care tests for diagnosis of current SARS-CoV-2 infections in children under real-life conditions.

**Design:** Systematic review and meta-analysis.

**Data sources:** MEDLINE, Embase, Cochrane Database for Systematic Reviews, INAHTA HTA database, preprint servers (via Europe PMC), ClinicalTrials.gov, WHO ICTRP from 1 January 2020 to 7 May 2021; NICE Evidence Search, NICE Guidance, FIND Website from 1 January 2020 to 24 May 2021.

**Review methods:** Diagnostic cross-sectional or cohort studies that included paediatric study participants and evaluated rapid point-of care tests for diagnosing current SARS-CoV-2 infections against RT-PCR as the reference standard were eligible for inclusion. QUADAS-2 was used to assess the risk of bias and the applicability of the included studies. Bivariate meta-analyses with random effects were performed. Variability was assessed by subgroup analyses.

**Results:** 17 studies with a total of 6355 paediatric study participants were included. All studies compared antigen tests against RT-PCR. Overall, studies evaluated eight antigen tests from six different brands. Only one study was at low risk of bias. The pooled overall diagnostic sensitivity and specificity in paediatric populations was 64.2% (95% CI: 57.4% to 70.5%) and 99.1% (95% CI: 98.2% to 99.5%), respectively. In symptomatic children, the pooled diagnostic sensitivity was 71.8% (95% CI: 63.6% to 78.8%) and the pooled diagnostic specificity was 98.7% (95% CI: 96.6% to 99.5%). The pooled diagnostic sensitivity in asymptomatic children was 56.2% (95% CI: 47.6% to 64.4%) and the pooled diagnostic specificity was 98.6% (95% CI: 97.3% to 99.3%).

**Conclusions:** The performance of current antigen tests in paediatric populations under real-life conditions varies broadly. Relevant data was only identified for about 1% of antigen tests with market access in Europe. The results should be interpreted with caution since risk of bias in studies was predominantly judged as unclear due to poor reporting. Further, the most common paediatric use cases (e.g., self-testing in schools or toddlers getting tested by their parents before leaving for kindergarten) have not been addressed in clinical performance studies yet. Policymakers should scrutinise whether the observed low diagnostic sensitivity may impact the planned purpose of antigen tests and are encouraged to require the evaluation of tests in the intended setting prior to the broad implementation of testing programmes.

**Systematic review registration:** CRD42021236313 (PROSPERO).

**SUMMARY BOXES:** *What is already known on this topic?:*

- During the current SARS-CoV-2 pandemic, antigen tests are widely used to detect children with current SARS-CoV-2 infection in schools and kindergarten despite an ongoing debate on potential benefits and harms.
- A recent Cochrane review showed that sensitivity estimates of antigen tests in adult populations vary broadly and are substantially lower as reported by manufacturers; however, test performance in paediatric populations remained unknown.

*What this study adds?:*

- A systematic literature search and comprehensive author queries allowed to include 17 studies evaluating the diagnostic accuracy of antigen tests in children.
- Based on the results of the bivariate meta-analysis, the real-life performance of current antigen tests for professional use in paediatric populations is below the minimum performance criteria set by WHO, the US FDA, or the MHRA in the UK.
- Considering the unclear risk of bias in most studies due to poor reporting, the performance of antigen tests for professional use in paediatric populations is similar to what has been reported previously for adult populations.
- There is an urgent need to evaluate the performance of antigen tests for the most common paediatric use cases since the impact of context-specific factors, e.g., sample collection in toddlers by laypersons or self-testing performed by children, on the diagnostic test accuracy – and most notably on the diagnostic sensitivity – remains unknown.

*How might it impact clinical practice in the foreseeable future?:*

- The results presented in this systematic review may support the development and improvement of testing programmes in schools and kindergarten.
- The evidence gaps identified in this systematic review demonstrate current research needs in the context of rapid point-of-care testing for SARS-CoV-2 infection in paediatric populations to support evidence-based decision making.

## INTRODUCTION

Since the beginning of the coronavirus disease 19 (COVID-19) pandemic caused by the novel severe acute respiratory syndrome coronavirus 2 (SARS-CoV-2) in spring 2020, an accurate, fast, and early detection of individuals infected with SARS-CoV-2 followed by effective isolation measures of infected individuals has been considered a cornerstone in the global fight against the spread of SARS-CoV-2. Laboratory-based reverse transcription-polymerase chain reaction (RT-PCR) testing is the standard for diagnosing current infections with SARS-CoV-2. However, limited testing capacities at many laboratories worldwide and limited availability of laboratories in developing countries demonstrated the urgent need for novel diagnostic tests that are easy to use, less expensive, widely available, and suitable for point-of-care use. Today, such tests – and in particular antigen tests – are increasingly used to complement testing with RT-PCR to extend testing capacities or when a short turnaround time is essential [1]. However, the advantages of antigen tests come at the price of lower diagnostic accuracy, most notably a lower diagnostic sensitivity, which increases the risk of missing cases, including pre-symptomatic infected individuals who have yet to enter the most infectious period [2].

Whether a lower sensitivity can be compensated by frequent testing remains controversially discussed [3–6]. Further, the fact that sensitivity and specificity are no inherent test characteristics but are rather affected by various factors, including population characteristics, sample quality, and study design, needs consideration [7]. Data on diagnostic accuracy provided by antigen test manufacturers at market access is often overly optimistic and does not necessarily reflect the test’s performance in practice. A recent Cochrane Review [8] showed that the sensitivity of antigen tests in adult populations varies considerably across brands, with only a few tests meeting the minimum acceptable sensitivity of ≥ 80% as defined by the World Health Organisation (WHO) or the United States Food and Drug Administration (US FDA) [9,10].

Since many countries are implementing public health safety measures that involve the use of antigen tests not only in adults but also in children, such as mass (self-)testing in schools [11], knowledge about how these tests perform in children is of high importance. However, to our knowledge, no systematic review has analysed the diagnostic test accuracy (DTA) of rapid tests in children yet. Therefore, in this systematic review and meta-analysis, we aimed to identify, assess, and summarise the best available evidence on the real-life performance of rapid tests for diagnosing current SARS-CoV-2 infections in paediatric populations at the point of care.

## METHODS

The protocol for this systematic review was registered with PROSPERO, the international prospective register of systematic reviews (ID: CRD42021236313) [12]. The reporting of this systematic review adhered to the Preferred Reporting Items for Systematic reviews and Meta-Analyses of DTA studies (PRISMA-DTA) guideline [13] and two relevant extensions “PRISMA-DTA for Abstracts” [14] and “PRISMA-S for Reporting Literatures Searches in Systematic Reviews” [15].

### Eligibility criteria

We included diagnostic cross-sectional and cohort studies that evaluated the clinical performance of rapid point-of-care tests for detecting current SARS-CoV-2 infections against the reference standard in paediatric or mixed-age populations. Assessing analytical performance parameters such as the analytical sensitivity (limit of detection) or the analytical specificity (cross-reactivity) was not subject of the current review. Diagnostic case-control studies were excluded as they reflect the test’s performance under ideal conditions and, therefore, often overestimate the diagnostic accuracy [16]. Moreover, studies evaluating serological tests were excluded as such tests are not suitable for the initial diagnosis of current SARS-CoV-2 infection [17]. We considered a study as eligible if the study population comprised at least ten paediatric study participants, each identified as positive or negative by the reference standard. In the absence of a true gold standard, laboratory-based RT-PCR alone or in combination with clinical findings or clinical follow-up was defined as the reference standard since it reflects the best available method for diagnosing individuals currently infected with SARS-CoV-2 [7]. Further, we required reporting of data that allowed constructing a complete 2×2 contingency table. The full set of eligibility criteria is shown in Table 1.

**Table 1:**
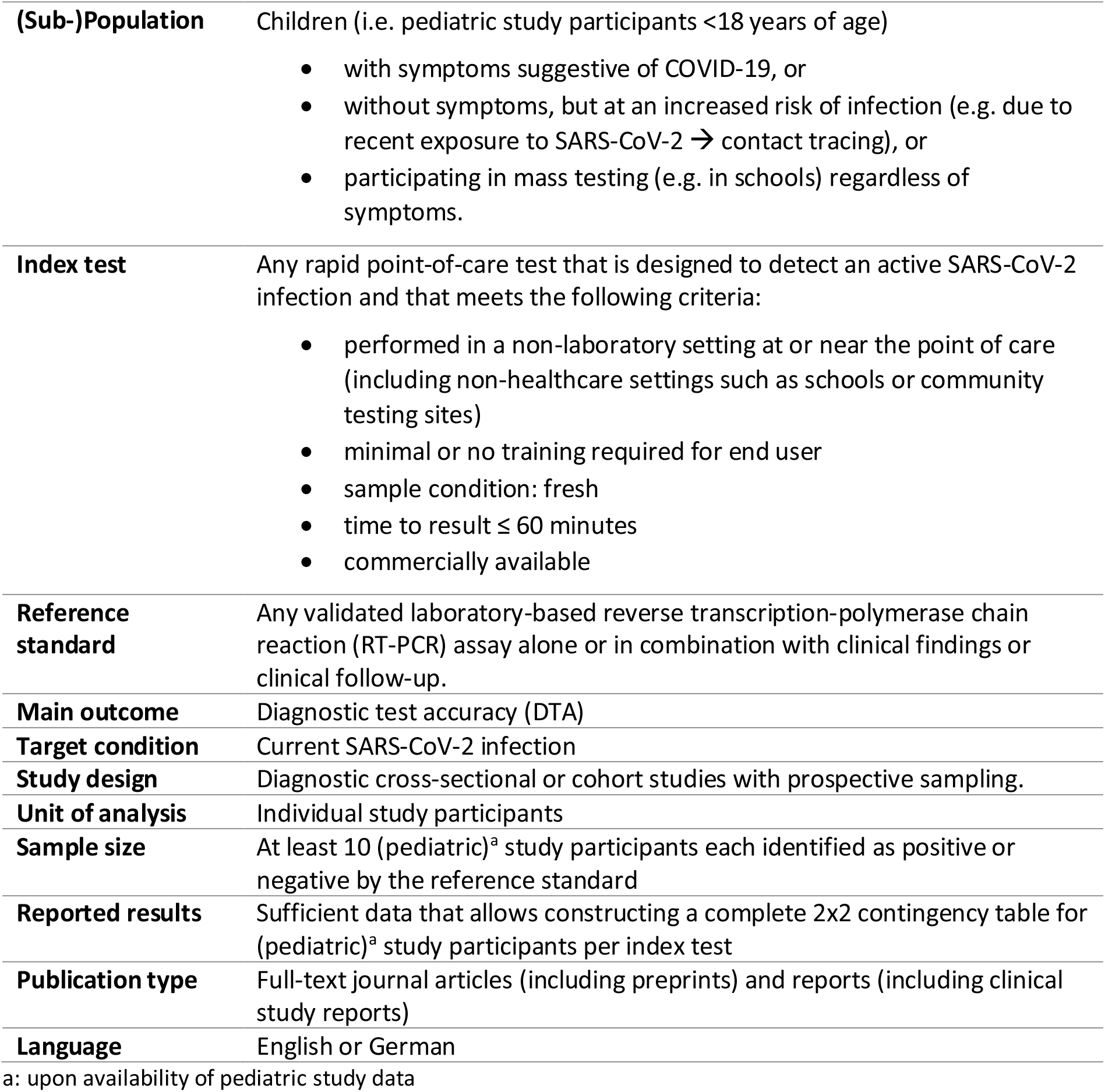
Eligibility criteria for primary studies

|  |  |
| --- | --- |
| **(Sub-)Population** | Children (i.e. pediatric study participants <18 years of age)<br>• with symptoms suggestive of COVID-19, or<br>• without symptoms, but at an increased risk of infection (e.g. due to recent exposure to SARS-CoV-2 → contact tracing), or<br>• participating in mass testing (e.g. in schools) regardless of symptoms. |
| **Index test** | Any rapid point-of-care test that is designed to detect an active SARS-CoV-2 infection and that meets the following criteria:<br>• performed in a non-laboratory setting at or near the point of care (including non-healthcare settings such as schools or community testing sites)<br>• minimal or no training required for end user<br>• sample condition: fresh<br>• time to result ≤ 60 minutes<br>• commercially available |
| **Reference standard** | Any validated laboratory-based reverse transcription-polymerase chain reaction (RT-PCR) assay alone or in combination with clinical findings or clinical follow-up. |
| **Main outcome** | Diagnostic test accuracy (DTA) |
| **Target condition** | Current SARS-CoV-2 infection |
| **Study design** | Diagnostic cross-sectional or cohort studies with prospective sampling. |
| **Unit of analysis** | Individual study participants |
| **Sample size** | At least 10 (pediatric)<sup>a</sup> study participants each identified as positive or negative by the reference standard |
| **Reported results** | Sufficient data that allows constructing a complete 2x2 contingency table for (pediatric)<sup>a</sup> study participants per index test |
| **Publication type** | Full-text journal articles (including preprints) and reports (including clinical study reports) |
| **Language** | English or German |

a: upon availability of pediatric study data

For studies that did not fully meet the inclusion criteria for the population, index test and/or reference standard, we required that at least 80% of the paediatric (sub-)population matched the population we defined for this systematic review. Studies were excluded, if the index test and the reference standard were performed in less than 80% of the paediatric study population. Besides journal articles, reports (including clinical study reports) that adhered to reporting standards such as STARD (Standards for Reporting of Diagnostic Accuracy Studies) [18] or recommendations given by government agencies [10,19] were considered eligible for inclusion. Studies that mentioned the inclusion of paediatric study participants without reporting any corresponding outcome data but otherwise met the eligibility criteria were preliminary included, and study authors were contacted and asked to provide such data. Further, if the study population’s baseline characteristics included information on age, we estimated the proportion of paediatric study participants assuming ages of study participants following a normal distribution and the proportion of PCR-positive paediatric assuming no changes in the PCR positivity rate among age groups. We contacted authors if we estimated at least 10 PCR-positive paediatric study participants in the study population.

### Information sources

We performed a comprehensive search for primary studies and secondary publications (systematic reviews and Health Technology Assessment (HTA) reports) in the following electronic bibliographic databases: MEDLINE (Ovid), Embase (Ovid), the Cochrane Library (Wiley), and preprint servers (Europe PMC) including medRxiv and bioRxiv (see [20] for full list of included preprint servers). Here, secondary publications were solely used as sources for potentially relevant studies. In addition, we searched two study registries (ClinicalTrials.gov and the WHO International Clinical Trials Registry Platform (ICTRP)) for relevant clinical studies. Other information sources comprised the International HTA Database, the Foundation of Innovative Diagnostics (FIND) COVID-19 website, and the Evidence Search and Guidance websites of Britain’s National Institute for Health and Care Excellence (NICE).

### Search strategy

In accordance with the Cochrane Handbook for DTA Reviews [21], the search strategy included concepts addressing the index test and the target condition. The development of the search strategy followed an objective approach that involved text-analytic procedures to identify candidate search terms based on the method described by Hausner et al. [22]. One researcher performed analyses of simple word frequencies and keywords-in-contexts in R using the “quanteda” package [23]. Because of substantial differences between types of tests, separate test sets were used to identify candidate search terms for antigen tests and molecular tests, respectively. Test sets included potentially relevant studies (irrespective of paediatric study participants) from the Cochrane Review by Dinnes et al. [8] and from a frequently updated website that lists DTA studies on antigen tests [24]. Due to a limited number of potentially relevant references addressing rapid molecular tests for point-of-care usage, the draft search strategy was supplemented by search terms derived from a conceptual approach. Furthermore, brand names of tests included in the Cochrane Review were added to increase sensitivity. The final search strategy was tested for completeness against the validation sets and relevant references of studies that included paediatric participants identified via exploratory searches beforehand. Prior to execution, the search strategy was peer-reviewed by a senior information specialist following the Peer Review of Electronic Search Strategies (PRESS) Guideline statement [25].

We only searched for publications published after December 2019, as we were only interested in literature published after the emergence of SARS-CoV-2. Further, we limited our search to publications written in English or German. Since Embase and MEDLINE provided comprehensive search filters for SARS-CoV-2 related literature via Ovid, our concept addressing the target condition was not used in these two searches.

To acknowledge the unprecedented role of preprints in the rapid dissemination of SARS-CoV-2 related research, we also searched for relevant preprints. Due to the direct availability of full texts of preprints and to increase the efficiency of the information retrieval, a further concept addressing the target population was defined and used in addition to the standard search strategy for identifying potentially relevant preprints directly at the full-text level. We assumed that this approach allowed to increase the precision of the overall search without a relevant reduction of its comprehensiveness.

All search strategies are provided in Appendix 1. The last search in bibliographic databases and study registries was conducted on May 7, 2021. Other information sources were last searched on May 24, 2021. Endnote X9.3 was used for citation management. Due to the more specific separate search for preprints at full-text level, any preprint records identified from MEDLINE were removed. Duplicates were initially eliminated via Ovid’s deduplication feature. After exporting all identified references from Ovid, duplicates were identified in R by comparing digital object identifiers (DOIs) of references from MEDLINE and Embase, and the Embase records of duplicates were removed. Remaining duplicates were manually removed in EndNote X9.3 and by using Endnote’s “find duplicates” function. Further, records from ClinicalTrials.gov that were retrieved from the WHO’s ICTRP website were removed since directly accessing ClinicalTrials.gov’s registry data allows for a more comprehensive search for relevant studies.

### Study selection

The screening of literature retrieved from bibliographical databases involved a two-step screening procedure and was performed independently by two researchers using the web-based Trial Selection Database (webTSDB) [26]. In a first step, potentially eligible primary studies and secondary publications were identified from screening titles and abstracts of retrieved citations. In a second step, the full texts of these articles were obtained and evaluated. Publications that met the eligibility criteria were included. Any discrepancies were resolved by consensus between the two researchers before finalizing each screening step. Reference lists of relevant systematic reviews and HTAs (independent of mentioning paediatric study participants) were manually screened to identify further relevant studies. For the screening of records from study registries, both screening steps were combined. Furthermore, documents identified through searching other information sources were screened for eligibility or information about potentially relevant studies.

### Data collection

The individual steps of data collection were performed by one researcher. All output was checked by a second researcher to ensure its validity and completeness. Any disagreements were resolved by consensus. At first, a standardized Excel spreadsheet was developed for data extraction. The spreadsheet was piloted before data extraction commenced. Extracted data included information on the general study characteristics, study participant characteristics, index test, reference standard, flow and timing, and reported outcomes. A complete list of data extraction items is presented in Table 2.

**Table 2:**
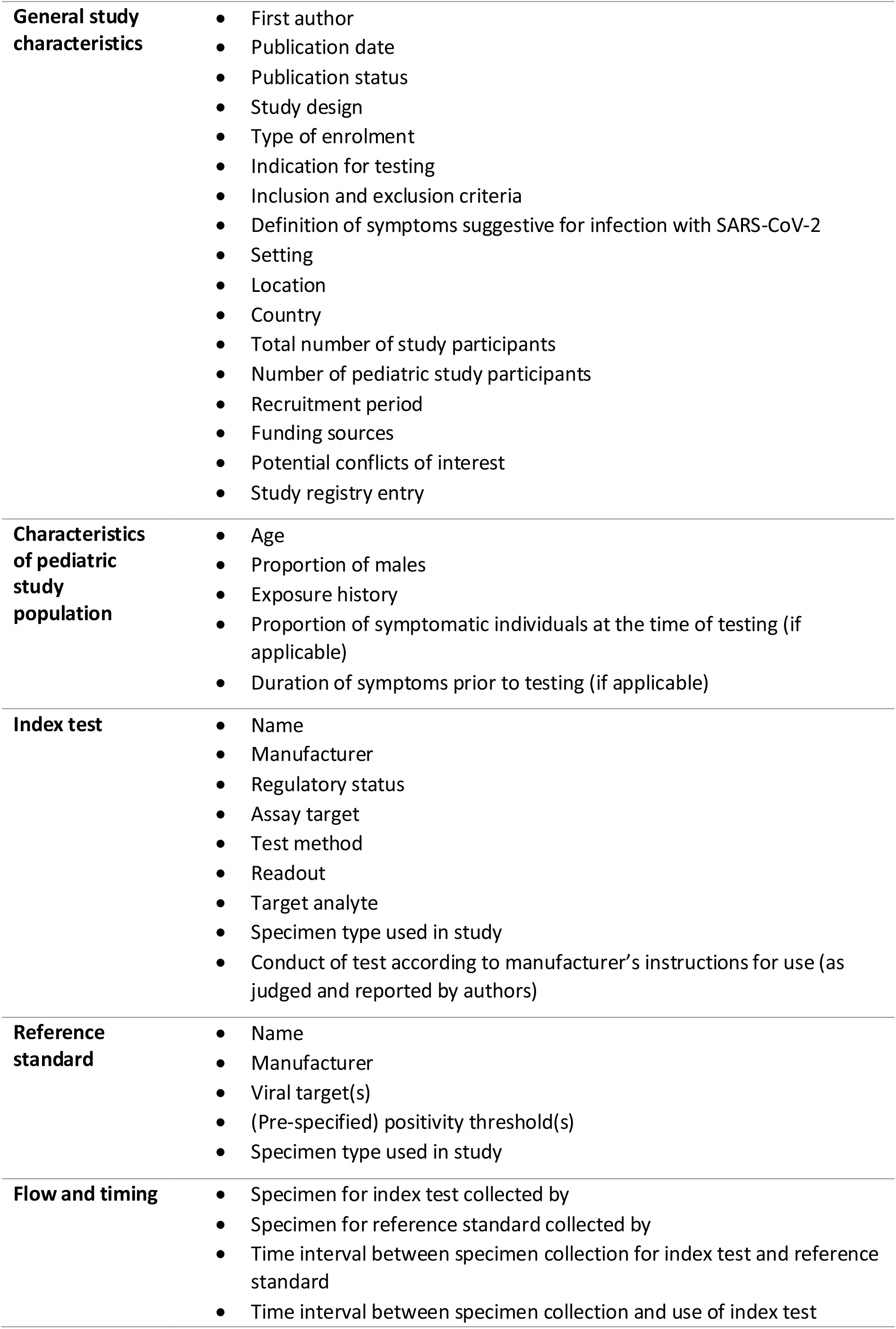
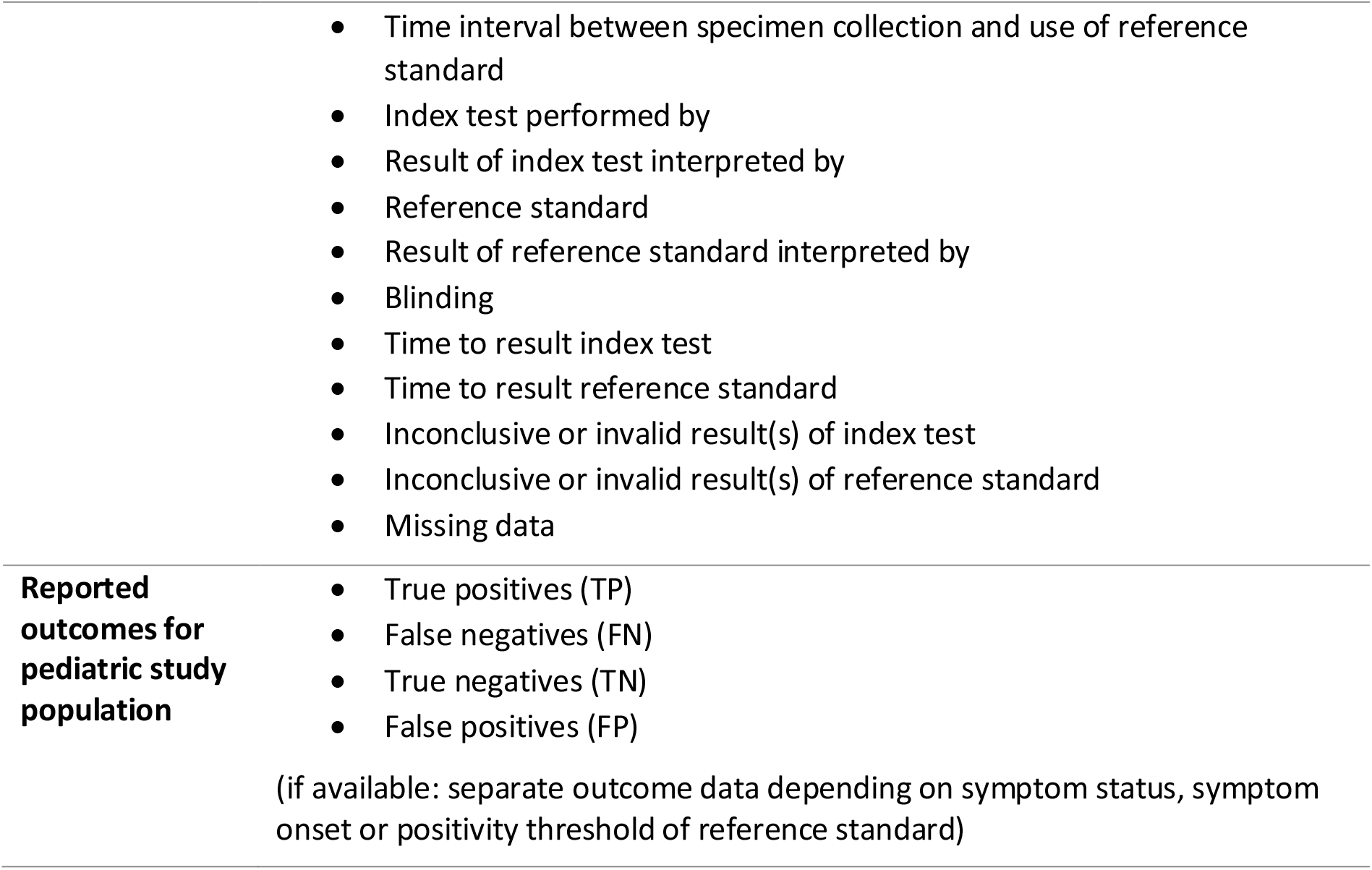
Data extraction items

### Quality assessment

We used the Quality Assessment of Diagnostic Accuracy Studies 2 (QUADAS-2) tool [27] to evaluate the methodological quality and applicability of the included studies at the study level. The tool requires assessing the risk of bias in the four domains patient selection, index test, reference standard, and flow and timing. Further, applicability concerns need to be assessed for the first three domains. Judgments are facilitated by answering pre-defined signalling questions with “yes”, “no”, or “unclear” for each domain. The tool was tailored to our review by adding one signalling question and review-specific guidance was provided to facilitate judgments, see Appendix 2. The quality assessment of each included study was performed by one researcher. A second researcher verified all judgments. Any disagreements were resolved by consensus. The results were summarised in the text and visualized as table and figure.

### Diagnostic accuracy measures and data synthesis

For each included study, diagnostic sensitivity, diagnostic specificity, positive predictive value (PPV) and negative predictive value (NPV) with corresponding 95% confidence intervals (CIs) were calculated based on the extracted 2×2 tables. Individual study participants were used as the unit of analysis throughout this work. If a study reported repeat testing of individuals only the initial test was included in our analyses. If a study evaluated more than one test in the same study population, we reported all test evaluations, but only one randomly chosen test was included in the meta-analyses to avoid the necessity to adjust for multiplicity.

The meta-analyses were based on recommendations provided in the methodological guideline “Meta-analysis of diagnostic test accuracy studies” by the European Network for Health Technology Assessment (EUnetHTA) [28]. Summary estimates for sensitivity and specificity were derived as follows: if sufficient data was available and the level of heterogeneity allowed meaningful statistical pooling, bivariate meta-analysis with random effects following the approach by Reitsma et al [29–31] was performed. Otherwise, separate univariate meta-analysis was performed. The bivariate approach required a continuity correction to handle zero cells in 2×2 tables. Thus, in studies where zero events were observed in one of the four cells, a continuity correction was applied by adding 0.5 to all four cells.

Depending on the availability of suitable data, subgroup analyses were performed to assess variables that could have an impact on a test’s diagnostic accuracy, such as the study participants’ presence of symptoms prior to testing and the duration of symptoms prior to testing. The influence of the publication status (preprint vs. peer-reviewed article) was evaluated as well as subgroup analyses with respect to the type of test (antigen vs. molecular; most commonly used antigen tests), setting (community vs. hospital-based), sample type ((oro-) nasopharyngeal vs. anterior nasal for index test and reference standard, respectively), end-user (layperson (self-testing) vs. trained staff/health care worker), and RT-PCR Ct value (cut-off values of 25 and 30). Differences between subgroups were assessed within the bivariate model and tested for statistical significance using the likelihood ratio test between the standard model and the model, which includes the corresponding variable. In the case of few studies in a subgroup analysis, univariate analysis for sensitivity and specificity were performed as sensitivity analysis and results were reported if remarkable differences between bivariate and univariate analysis were observed.

All statistical analyses were performed using the statistical platform R version 4.1.0 [32]. Bivariate meta-analysis was performed, along with the construction of the corresponding figures, with the package “mada” [33], while univariate meta-analysis was performed with the package “meta” [34] and “PropCIs” [35]. 95% CIs were computed using the approach proposed by Wilson [36].

### Patient and public involvement

Neither patients nor members of the public were actively involved in the conduct of this systematic review. However, the development of the research question was motivated by public discussions about SARS-CoV-2 antigen testing in children. Further, we were – and still are – in continuous exchange with teachers, parents, and children about practical aspects and the impact of current testing programmes in schools and kindergarten.

## RESULTS

### Study selection

Overall, 3011 records were retrieved from five bibliographic databases. The PRISMA flow diagram is shown in Figure 1 and outlines the process of identifying relevant studies from different information sources. References that were excluded at the full-text level can be found in Appendix 3 with the reason for their exclusion. After removing 36 preprint records identified via MEDLINE and 680 duplicate records, 2295 records were screened for eligibility. 2078 records were excluded at the title/abstract level. Full-text publications of 217 records were retrieved for further assessment. Nine studies [37–45] met all eligibility criteria for inclusion. Furthermore, 21 studies [46–66] were identified as eligible for author queries to obtain study data on paediatric subgroups. Authors of nine studies did not respond to our request for data [46,47,52,53,55,57–59,62]. In four cases [49,60,65,66], authors reported that the required number of individuals who tested positive or negative by the reference standard was not reached, and in one case [56] no data on age were recorded. Eventually, author queries led to the inclusion of eight further studies [48,50,51,54,61,63,64,67] resulting in a total of 17 relevant studies for this review (12 peer-reviewed journal articles and five preprints). The full list of included studies is reported in Table 3.

**Figure 1:**
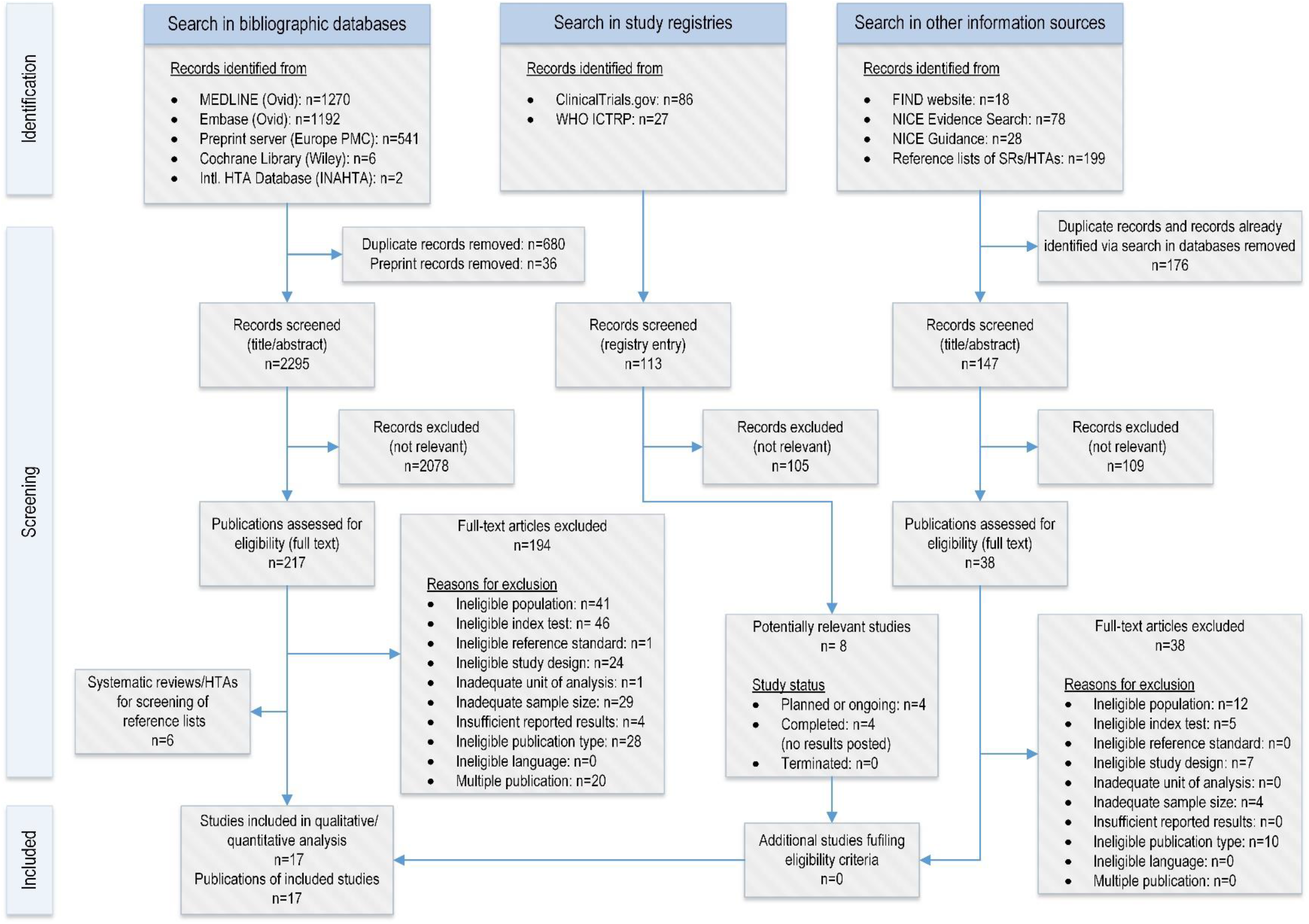
PRISMA Flow diagram illustrating the selection process of primary studies included in this systematic review and meta-analysis.

**Table 3:** Study pool of the systematic review

| **Study identifier** | **Data sources** | **Registry entry** |
| --- | --- | --- |
| Akingba | Preprint [48], unpublished data provided by author | N/A |
| Bianco | Journal article [50], unpublished data provided by author | N/A |
| Drevinek | Preprint [51], unpublished data provided by author | N/A |
| Gonzalez-Donapetry | Journal article [37] | N/A |
| Homza | Journal article [54], unpublished data provided by author | N/A |
| Kiyasu | Preprint [67], unpublished data provided by author | N/A |
| L'Huillier | Preprint [38] | N/A |
| Möckel | Journal article [39] | yes<sup>a</sup> |
| Pilarowski | Journal article [40] | N/A |
| Pollock a | Journal article [41] | N/A |
| Pollock b | Preprint [42] | N/A |
| Prince-Guerra | Journal article [43] | N/A |
| Sood | Journal article [44] | N/A |
| Shah | Journal article [61], supplementary file provided by authors (published as part of preprint [74] after finalization of the study pool) | N/A |
| Takeuchi | Journal article [63], unpublished data provided by author | N/A |
| Torres | Journal article [64], unpublished data provided by author | N/A |
| Villaverde | Journal article [45] | N/A |

N/A: not available
a: Study registry identifier: DRKS00019207

Furthermore, we screened 113 records identified from study registries and 323 records identified from other information sources. The search for studies in study registries allowed to identify four planned or ongoing and four completed studies with no results posted, see Table 4 for further details. Information retrieval from other information sources included screening 18 records retrieved from the FIND website, 78 records from NICE Evidence Search, 28 records from NICE, and 23 records from reference lists of six systematic reviews [8,68–72] identified via searching bibliographic databases. As a result, no additional study that met the inclusion criteria was identified.

**Table 4:** Potentially relevant studies identified through searching clinical study registries

| **Study registry identifier** | **Country** | **Enrollment** | **Ages eligible for study** | **Name of index test** | **Recruitment status<sup>a</sup> (study completion date)** |
| --- | --- | --- | --- | --- | --- |
| NCT04513990 | USA | 1,500 | not specified (children included) | unspecified point-of-care test | Recruiting (estimated: Apr 2021) |
| NCT04557046 | USA | 400 | any | LumiraDx SARS-CoV-2 Ag Test | Recruiting (estimated: Dec 2021) |
| NCT04583189 | France | 500 | ≤18 years | Biosynex Covid-19 Ag BSS Rapid test | Completed (Nov 2020) |
| NCT04720235 | USA | 304 | 14-75 years | Lucira COVID-19 All-In-One Test Kit | Completed (Mar 2021) |
| NCT04750629 | USA | 100 | ≥ 1 year | CovidX™ Rapid Antigen Test | Not yet recruiting (estimated: Mar 2021) |
| NCT04808921 | USA | 151 | any | Xiamen Wiz Biotech SARS-CoV-2 Antigen Rapid Test | Completed (Apr 2021) |
| NCT04859023 | France | 10,000 | ≥10 years | unspecified SARS-CoV-2 antigen test | Completed (Feb 2021) |
| NCT04878068 | not reported | 300 | ≥ 12 years | Therma COVID-19 Rapid Antigen Test | Not yet recruiting (estimated: June 2021) |

a: Status as of May 24, 2021

### Study characteristics

All 17 included studies (6355 paediatric study participants) evaluated the performance of antigen tests against the reference standard RT-PCR. The main study characteristics for each individual study are summarised in Table 5, further details are reported in Table 6 and Table 7. While 14 studies evaluated the test performance in mixed-age populations, including 24 to 928 paediatric study participants, three studies with a sample size between 440 and 1620 individuals exclusively recruited children. In eight studies, the purpose of testing included diagnostic testing of symptomatic individuals suggestive of SARS-CoV-2 infection. Six studies reported the inclusion of asymptomatic individuals at an increased risk of infection due to previous exposure to SARS-CoV-2. Here, “asymptomatic” refers to any individual who is healthy, infected but pre-symptomatic or infected but without symptoms. Evaluating the performance of antigen tests in a screening setting (e.g. community mass testing) was the main objective of six studies. Eight antigen tests (six lateral flow immunochromatographic assays and two fluorescent immunoassays) from six different brands were used in 18 test evaluations, whereas antigen tests by Abbott were most investigated (Panbio COVID-19 Ag Rapid Test n=6, BinaxNOW COVID-19 Ag Card n=5). In more than half of the test evaluations (n=11), nasopharyngeal samples were collected for the index test. Six test evaluations used anterior nasal specimens for the index test. In all studies, the reference standard was RT-PCR performed in a laboratory setting.

**Table 5:** Characteristics of included studies. All studies aimed at identifying individuals currently infected with SARS-CoV-2. The reference standard was RT-PCR performed in the laboratory setting.

| Study identifier | Publication status (year) | Study design | Setting | Purpose of testing | Location (recruitment period) | Name of index test | Total number of study participants | Pediatric study participants |  |  |  | Funding / potential COI |
| --- | --- | --- | --- | --- | --- | --- | --- | --- | --- | --- | --- | --- |
|  |  |  |  |  |  |  |  | n | male (%) | symptomatic (%) | age (years) |  |
| **Akingba<sup>a</sup> [48]** | preprint (2021) | cross-sectional | Community testing site (mobile clinic) | D | South Africa (Nov 20) | Panbio COVID-19 Ag Rapid Test | 667 | 41 | 39.0 | 100<sup>b</sup> | median: 13<sup>c</sup><br>IQR: 10-16<sup>c</sup><br>range: 3-17<sup>c</sup> | none / none |
| **Bianco<sup>a</sup> [50]** | published (2021) | cross-sectional | Hospital ED, Hospital unit | n.r. | Italy (Oct-Dec 20) | LumiraDx SARS-CoV-2 Ag Test | 907<sup>d</sup> | 165 | n.r. | 9.7 | mean: 7.2<br>range: 0-18 | none / none |
| **Drevinek<sup>a</sup> [51]** | preprint (2020) | cross-sectional | Hospital TC | D, A, S | Czech Republic (Oct 20) | 1: Panbio COVID-19 Ag Rapid Test<br>2: Standard F Covid-19 FIA Ag | 591 | 31 | 38.7<sup>c</sup> | 32.3<sup>c</sup> | median: 15<sup>c</sup><br>IQR: 13.5-16<sup>c</sup><br>range: 11-17<sup>c</sup> | public / none |
| **Gonzalez-Donapetry [37]** | published (2021) | cross-sectional | Hospital ED | D | Spain (Sept-Oct 20) | Panbio COVID-19 Ag Rapid Test | 440 | 440 | 59.1 | 100<sup>b</sup> | median: 3<br>IQR: 1-7<br>range: 0-15 | none / none |
| **Homza<sup>a</sup> [54]** | published (2021) | cross-sectional | Hospital TC | D, A | Czech Republic (not reported) | ECOTEST COVID-19 Antigen Rapid Test | 494 | 24 | 58.3 | 45.8 | mean: 13.17<br>SD: 2.79<br>range: 7-17 | public / none |
| **Kiyasu<sup>a,g</sup> [67]** | preprint (2021) | cohort | Hospital TC | n.r. | Japan (Oct 20-Jan 21) | QuickNavi COVID19 Ag | 1881 | 90 | 68.9 | 4.4 | median: 12<sup>c</sup><br>IRQ: 6-15<sup>c</sup><br>range: 0-17<sup>c</sup> | private / yes |
| **L'Huillier [38]** | preprint (2021) | cross-sectional | Hospital TC | D, A, S | Switzerland (Nov 20-Mar 21) | Panbio COVID-19 Ag Rapid Test | 885 | 885 | 50.1 | 64.8 | median: 11.8<br>IQR: 9.0-14.3<br>range: 0-16 | public / none |
| **Möckel [39]** | published (2021) | cross-sectional | Hospital ED | D | Germany (Oct-Nov 20) | Roche SARS-CoV-2 Rapid Antigen Test | 483 | 196<sup>h</sup> | 55 | 87.1 | median: 3<br>IQR: 1-9 | public / none |
| **Pilarowski [40]** | published (2020) | cross-sectional | Community testing site | S | USA (Nov-Dec 20) | BinaxNOW COVID-19 Ag Card | 3320 | 209 | n.r. | n.r.<br>30.9<sup>e</sup> | ≤18<br>(47% < 13) | public, private / yes |
| **Pollock a [41]** | published (2021) | cross-sectional | Community testing site | S | USA (Oct-Dec 20) | BinaxNOW COVID-19 Ag Card | 2482 | 928 | 47.0<sup>c</sup> | 10.7 | ≤18 (69%<sup>c</sup> ≤ 13) | public / none |
| **Pollock b [42]** | preprint (2021) | cross-sectional | Community testing site | S | USA (Jan 21) | CareStart COVID-19 Antigen test | 1603 | 253 | 46.2<sup>c</sup> | 12.6 | ≤18 (62%<sup>c</sup> ≤ 13) | public / none |
| **Prince-Guerra [43]** | published (2021) | cross-sectional | Community testing site | S | USA (Nov 20) | BinaxNOW COVID-19 Ag Card | 3419 | 236 | n.r. | n.r. 24.2<sup>e</sup> | range: 10-17 | n.r. / none |
| **Shah<sup>a</sup> [61,74]** | published (2021) | cohort | Community testing site | S | USA (Nov-Dec 20) | BinaxNOW COVID-19 Ag Card | 2024 | 217 | n.r. | 53.4<sup>c</sup> | range: 5-17 | public / none |
| **Sood [44]** | published (2021) | cross-sectional | Community testing site | S, A | USA (Nov-Dec 20) | BinaxNOW COVID-19 Ag Card | 1429 | 783 | 49.5 | 23.5 | range: 5-17<br>65%: 5-12<br>35%: 13-17 | public, private / yes |
| **Takeuchi<sup>a,g</sup> [63]** | published (2021) | cohort | Hospital TC | D, A | Japan (Oct-Dec 20) | QuickNavi COVID19 Ag | 1208 | 164 | 61.6 | 54.9 | median: 10<sup>c</sup><br>IQR: 5-14<sup>c</sup><br>range: 0-17<sup>c</sup> | private / yes |
| **Torres<sup>a</sup> [64]** | published (2021) | cross-sectional | Hospital TC | A | Spain (Oct-Nov 20) | Panbio COVID-19 Ag Rapid Test | 634 | 73 | 47.9 | 0<sup>f</sup> | median: 13<br>range: 9-17 | none / none |
| **Villaverde [45]** | published (2021) | cohort | Hospital ED | D | Spain (Sept-Oct 20) | Panbio COVID-19 Ag Rapid Test | 1620 | 1620 | n.r. | 100<sup>a</sup> | range: 0-16 | public / none |

**Table 6:** Characteristics of the studies’ index test and reference standard

| Study identifier | Name (manufacturer) | Index test |  |  | RT-PCR assay | Reference standard |  |  |
| --- | --- | --- | --- | --- | --- | --- | --- | --- |
|  |  | Test method<sup>a</sup> / readout<sup>a</sup> | Target analyte<sup>a</sup> | Specimen type used in study |  | Viral target | Positivity threshold | Specimen type used in study |
| **Akingba** | Panbio COVID-19 Ag Rapid Test (Abbott) | LFA / visual | N protein | NP | Seegene nCoV assay | 3 targets (not specified) | at least 1 target ct-value < 38<br>inconclusive: 2 targets negative and 1 target positive with ct-value ≥38 | same swab used for antigen test |
| **Bianco** | LumiraDx SARS-CoV-2 Ag Test (LumiraDx) | microfluidic FIA / automated | N protein | nasal | XpertXpress SARS-CoV-2 assay (Cepheid)<sup>b</sup> | n.r. | n.r. | NP |
| **Drevinek** | 1: Panbio COVID-19 Ag Rapid Test (Abbott)<br>2: Standard F Covid-19 FIA Ag (SD Biosensor) | 1: LFA / visual<br>2: FIA / automated | 1: N protein<br>2: N protein | 1: NP<br>2: NP | Allplex SARS-nCoV-2 (Seegene) | N, E and RdRP/S genes | at least one target with ct-value ≤ 40 | NP+OP |
| **Gonzalez-Donapetry** | Panbio COVID-19 Ag Rapid Test (Abbott) | LFA / visual | N protein | NP | Viracell SARS-CoV-2 real-time PCR kit (Viracell) | N and E gene | both targets with ct-value ≤ 40 | NP |
| **Homza** | ECOTEST COVID-19 Antigen Rapid Test (Assure Tech) | LFA / visual | N protein | NP (nostril 1) | COVID-19 Multiplex RT-PCR Kit (Diana Biotechnologies) | S gene and gene coding the EndoRNase | n.r. | NP (nostril 2) |
| **Kiyasu** | QuickNavi COVID19 Ag (Denka) | LFA / visual | N protein | NP | RT-PCR (National Institute of Infectious Diseases, Japan)<sup>c</sup> | n.r. | n.r. | NP |
| **L'Huillier** | Panbio COVID-19 Ag Rapid Test (Abbott) | LFA / visual | N protein | NP | 1: Cobas SARS-CoV-2 assay (Roche) or 2: Nimbus RT-PCR assay | 1: n.r.<br>2: n.r. | 1: unclear<br>2: unclear | NP |
| **Möckel** | Roche SARS-CoV-2 Rapid Antigen Test (SD Biosensors) | LFA / visual | N protein | ONP | 1: Cobas SARS-CoV-2 assay (Roche) or 2: SARS-CoV-2 E-gene assay (TibMolbiol) | 1: n.r.<br>2: E gene | 1: unclear<br>2: unclear | ONP |
| **Pilarowski** | BinaxNOW COVID-19 Ag Card (Abbott) | LFA / visual | N protein | AN (bilateral) | RenegadeXP<sup>d</sup> (RenegadeBio) | N gene | "none", ct cut-off value =30 or 35 | AN (bilateral) |
| **Pollock a** | BinaxNOW COVID-19 Ag Card (Abbott) | LFA / visual | N protein | AN (bilateral) | CRSP SARS-CoV-2 Real-time RT-PCR Diagnostic Assay (MIT/Harvard) | N2 gene | ct cut-off value = 40 (in addition: 25,30, 35) | AN (bilateral) |
| **Pollock b** | CareStart COVID-19 Antigen test (Access Bio) | LFA / visual | N protein | AN (bilateral) | CRSP SARS-CoV-2 Real-time RT-PCR Diagnostic Assay (MIT/Harvard) | N2 gene | ct cut-off value = 40 (in addition: 25,30, 35) | AN (bilateral) |
| **Prince-Guerra** | BinaxNOW COVID-19 Ag Card (Abbott) | LFA / visual | N protein | AN (bilateral) | 1: CDC 2019-nCoV Real-time RT-PCR Diagnostic Panel or 2: Fosun COVID-19 RT-PCR Detection Kit | 1: n.r.<br>2: n.r. | 1: n.r.<br>2: n.r. | NP (bilateral) |
| **Shah** | BinaxNOW COVID-19 Ag Card (Abbott) | LFA / visual | N protein | AN (nostril 1) | TaqPath SARS-CoV-2 Combo Kit (Thermo Fisher Scientific) | S, N and Orf1Ab genes | ct-value $\leq 37$ for at least two targets; inconclusive: 1 target positive | AN (nostril 2) |
| **Sood** | BinaxNOW COVID-19 Ag Card (Abbott) | LFA / visual | N protein | AN | Curative SARS-Cov-2 Assay (EUA for testing at KorvaLabs) | n.r. | ct-value $\leq 40$ | oral fluid<sup>e</sup> |
| **Takeuchi** | QuickNavi COVID19 Ag (Denka) | LFA / visual | N protein | NP | RT-PCR (National Institute of Infectious Diseases, Japan)<sup>f</sup> | n.r. | n.r. | NP |
| **Torres** | Panbio COVID-19 Ag Rapid Test (Abbott) | LFA / visual | N protein | NP (left nostril) | TaqPath SARS-CoV-2 Combo Kit (Thermo Fisher Scientific) | N gene | ct-value $\leq 35$ (in addition: $\leq 30$, $\leq 25$, $\leq 20$) | NP (right nostril) |
| **Villaverde** | Panbio COVID-19 Ag Rapid Test (Abbott) | LFA / visual | N protein | NP | RT-PCR (not further specified) | E and RdRp genes | n.r. | NP |

**Table 7:**
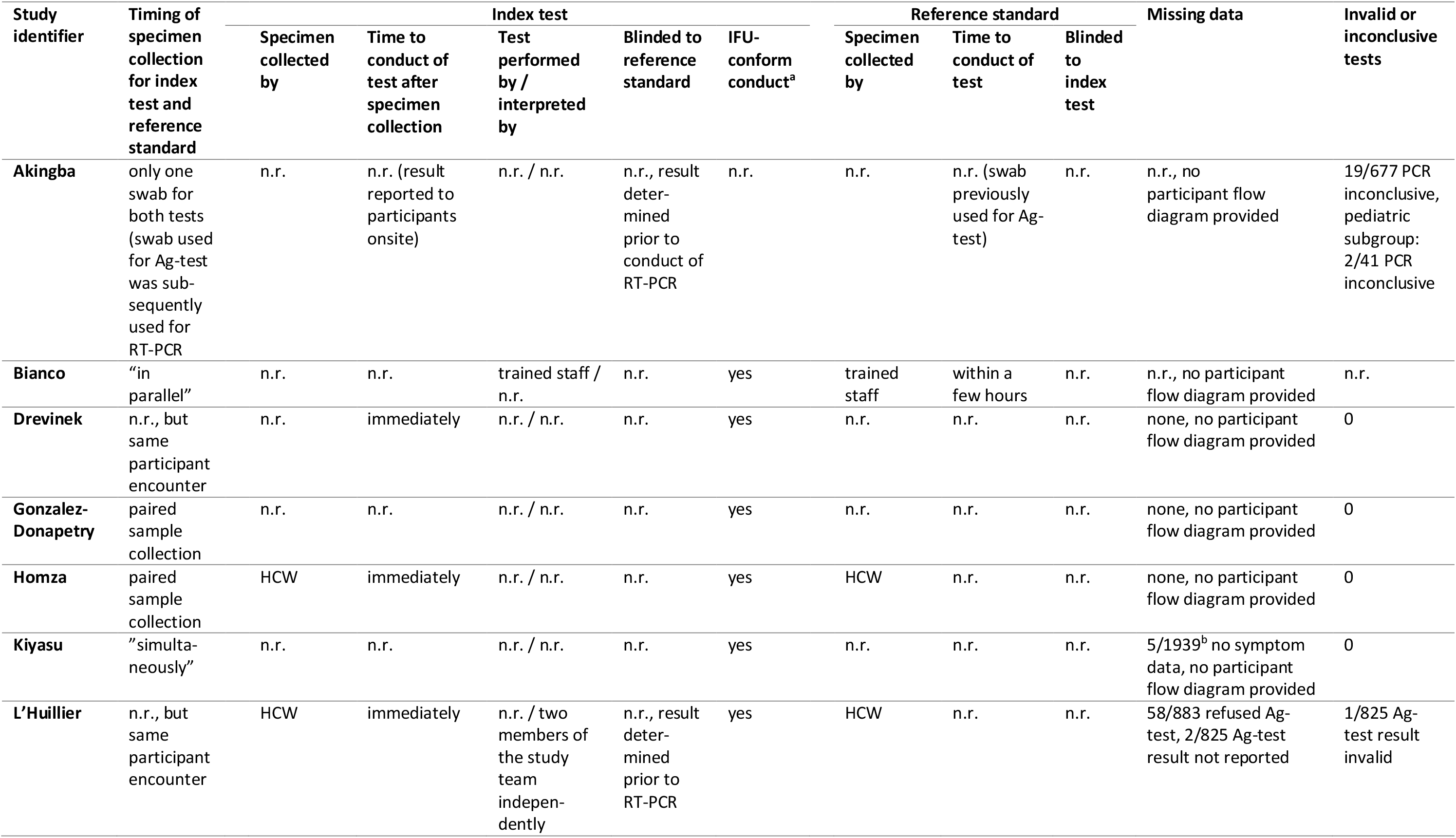
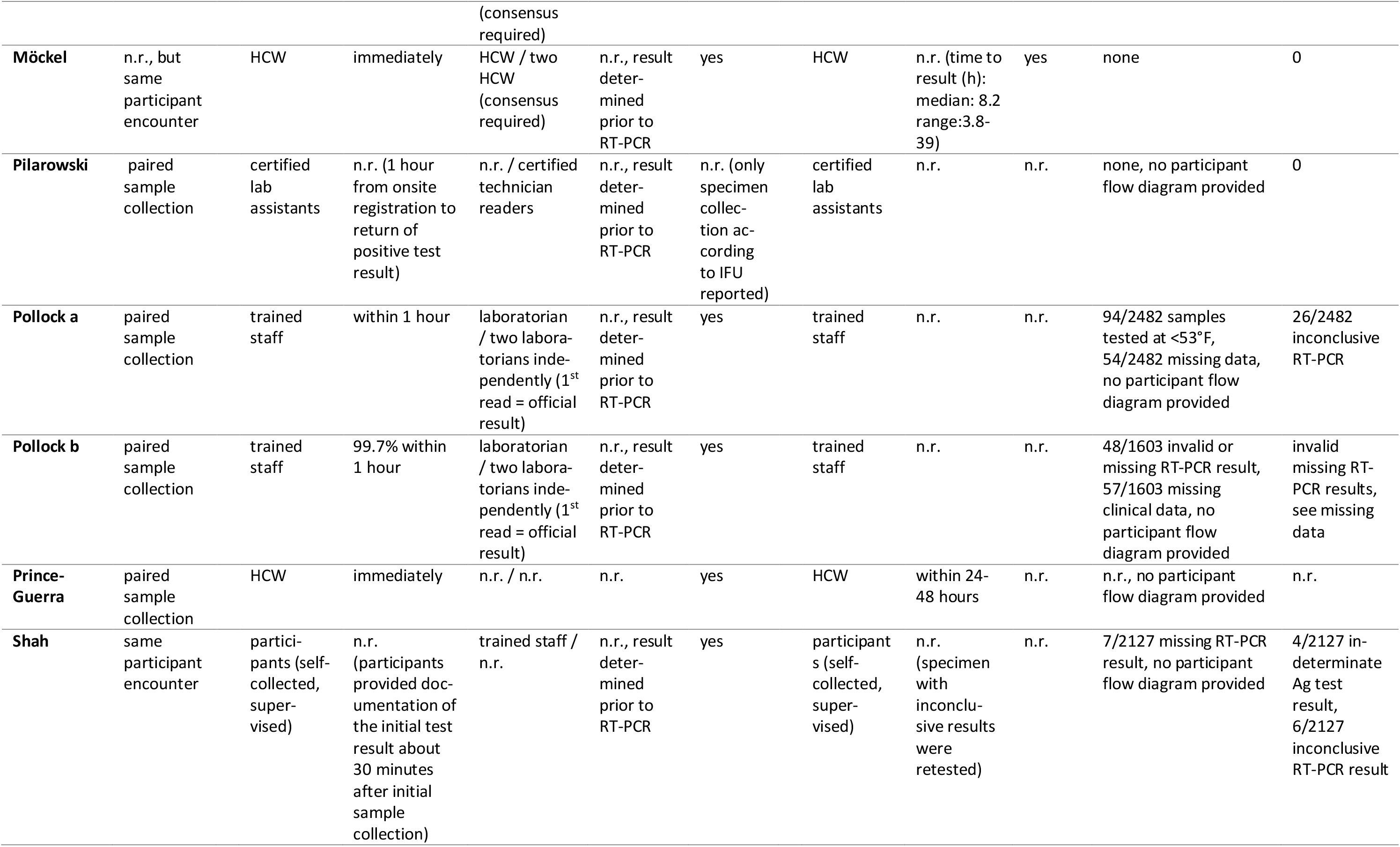
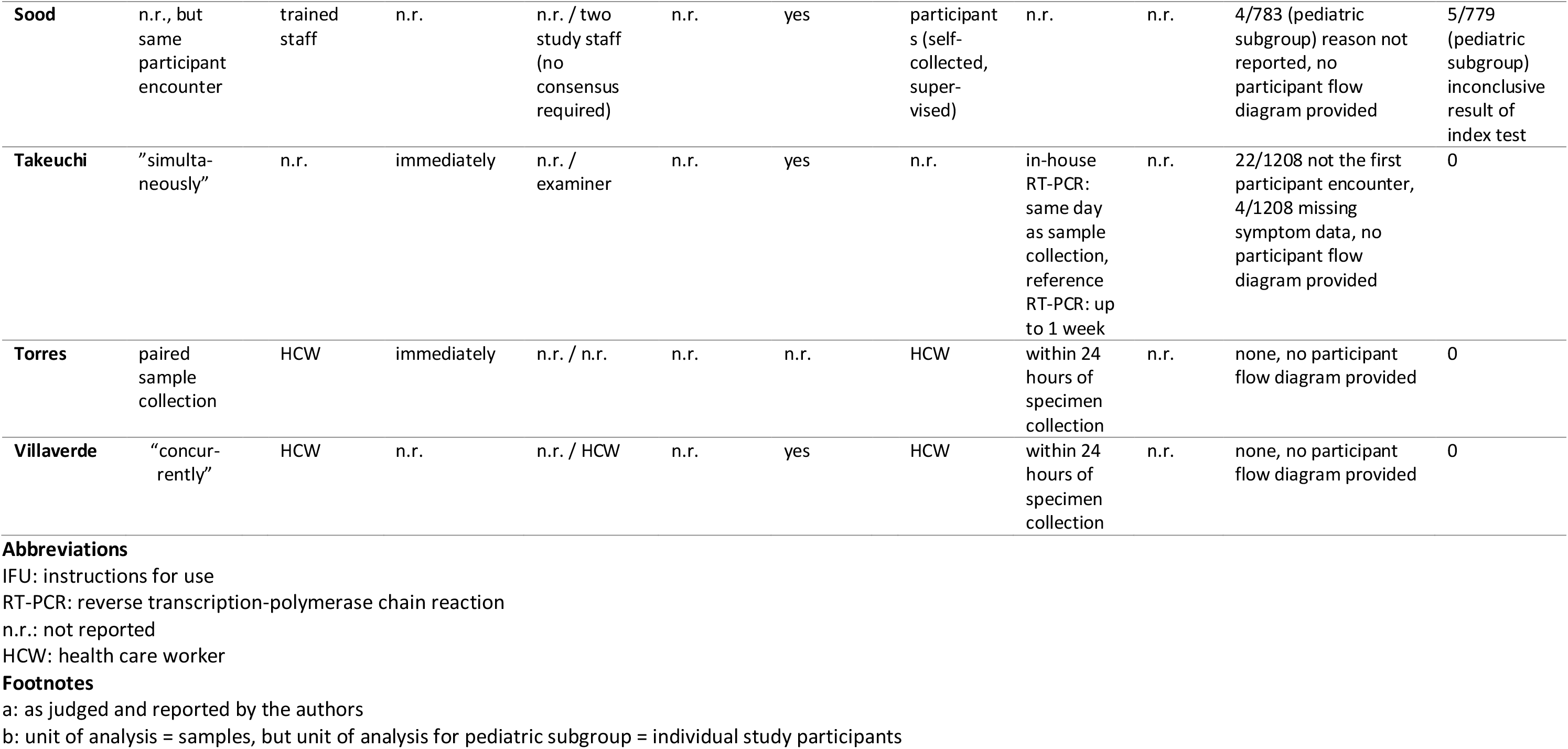
Conduct, flow and timing, and interpretation of index test and reference standard

| Study identifier | Timing of specimen collection for index test and reference standard | Index test |  |  |  |  | Reference standard |  |  | Missing data | Invalid or inconclusive tests |
| --- | --- | --- | --- | --- | --- | --- | --- | --- | --- | --- | --- |
|  |  | Specimen collected by | Time to conduct of test after specimen collection | Test performed by / interpreted by | Blinded to reference standard | IFU-conform conduct<sup>a</sup> | Specimen collected by | Time to conduct of test | Blinded to index test |  |  |
| **Akingba** | only one swab for both tests (swab used for Ag-test was subsequently used for RT-PCR) | n.r. | n.r. (result reported to participants onsite) | n.r. / n.r. | n.r., result determined prior to conduct of RT-PCR | n.r. | n.r. | n.r. (swab previously used for Ag-test) | n.r. | n.r., no participant flow diagram provided | 19/677 PCR inconclusive, pediatric subgroup: 2/41 PCR inconclusive |
| **Bianco** | "in parallel" | n.r. | n.r. | trained staff / n.r. | n.r. | yes | trained staff | within a few hours | n.r. | n.r., no participant flow diagram provided | n.r. |
| **Drevinek** | n.r., but same participant encounter | n.r. | immediately | n.r. / n.r. | n.r. | yes | n.r. | n.r. | n.r. | none, no participant flow diagram provided | 0 |
| **Gonzalez-Donapetry** | paired sample collection | n.r. | n.r. | n.r. / n.r. | n.r. | yes | n.r. | n.r. | n.r. | none, no participant flow diagram provided | 0 |
| **Homza** | paired sample collection | HCW | immediately | n.r. / n.r. | n.r. | yes | HCW | n.r. | n.r. | none, no participant flow diagram provided | 0 |
| **Kiyasu** | "simultaneously" | n.r. | n.r. | n.r. / n.r. | n.r. | yes | n.r. | n.r. | n.r. | 5/1939<sup>b</sup> no symptom data, no participant flow diagram provided | 0 |
| **L'Huillier** | n.r., but same participant encounter | HCW | immediately | n.r. / two members of the study team independently (consensus required) | n.r., result determined prior to RT-PCR | yes | HCW | n.r. | n.r. | 58/883 refused Ag-test, 2/825 Ag-test result not reported | 1/825 Ag-test result invalid |
| **Möckel** | n.r., but same participant encounter | HCW | immediately | HCW / two HCW (consensus required) | n.r., result determined prior to RT-PCR | yes | HCW | n.r. (time to result (h): median: 8.2 range:3.8-39) | yes | none | 0 |
| **Pilarowski** | paired sample collection | certified lab assistants | n.r. (1 hour from onsite registration to return of positive test result) | n.r. / certified technician readers | n.r., result determined prior to RT-PCR | n.r. (only specimen collection according to IFU reported) | certified lab assistants | n.r. | n.r. | none, no participant flow diagram provided | 0 |
| **Pollock a** | paired sample collection | trained staff | within 1 hour | laboratorian / two laboratorians independently (1<sup>st</sup> read = official result) | n.r., result determined prior to RT-PCR | yes | trained staff | n.r. | n.r. | 94/2482 samples tested at <53°F, 54/2482 missing data, no participant flow diagram provided | 26/2482 inconclusive RT-PCR |
| **Pollock b** | paired sample collection | trained staff | 99.7% within 1 hour | laboratorian / two laboratorians independently (1<sup>st</sup> read = official result) | n.r., result determined prior to RT-PCR | yes | trained staff | n.r. | n.r. | 48/1603 invalid or missing RT-PCR result, 57/1603 missing clinical data, no participant flow diagram provided | invalid missing RT-PCR results, see missing data |
| **Prince-Guerra** | paired sample collection | HCW | immediately | n.r. / n.r. | n.r. | yes | HCW | within 24-48 hours | n.r. | n.r., no participant flow diagram provided | n.r. |
| **Shah** | same participant encounter | participants (self-collected, supervised) | n.r. (participants provided documentation of the initial test result about 30 minutes after initial sample collection) | trained staff / n.r. | n.r., result determined prior to RT-PCR | yes | participants (self-collected, supervised) | n.r. (specimen with inconclusive results were retested) | n.r. | 7/2127 missing RT-PCR result, no participant flow diagram provided | 4/2127 indeterminate Ag test result, 6/2127 inconclusive RT-PCR result |
| **Sood** | n.r., but same participant encounter | trained staff | n.r. | n.r. / two study staff (no consensus required) | n.r. | yes | participants (self-collected, supervised) | n.r. | n.r. | 4/783 (pediatric subgroup) reason not reported, no participant flow diagram provided | 5/779 (pediatric subgroup) inconclusive result of index test |
| **Takeuchi** | "simultaneously" | n.r. | immediately | n.r. / examiner | n.r. | yes | n.r. | in-house RT-PCR: same day as sample collection, reference RT-PCR: up to 1 week | n.r. | 22/1208 not the first participant encounter, 4/1208 missing symptom data, no participant flow diagram provided | 0 |
| **Torres** | paired sample collection | HCW | immediately | n.r. / n.r. | n.r. | n.r. | HCW | within 24 hours of specimen collection | n.r. | none, no participant flow diagram provided | 0 |
| **Villaverde** | "concurrently" | HCW | n.r. | n.r. / HCW | n.r. | yes | HCW | within 24 hours of specimen collection | n.r. | none, no participant flow diagram provided | 0 |

### Risk of bias and applicability

The results of the quality assessment are summarised in Table 8 and Figure 2. Quality among studies varied. Only one study was at low risk of bias in all four domains of the QUADAS-2 tool. For patient selection, more than half of the studies were at high (n=1) or unclear (n=12) risk of bias because inadequate exclusion of participants occurred, or it was not clear whether a consecutive or random sample was enrolled into the study. All but one study was judged as having an unclear risk of bias for the reference standard due to insufficient reporting of blinding. Risk of bias in the flow and timing domain was high in three studies due to more than 5% of missing outcome data.

**Figure 2:**
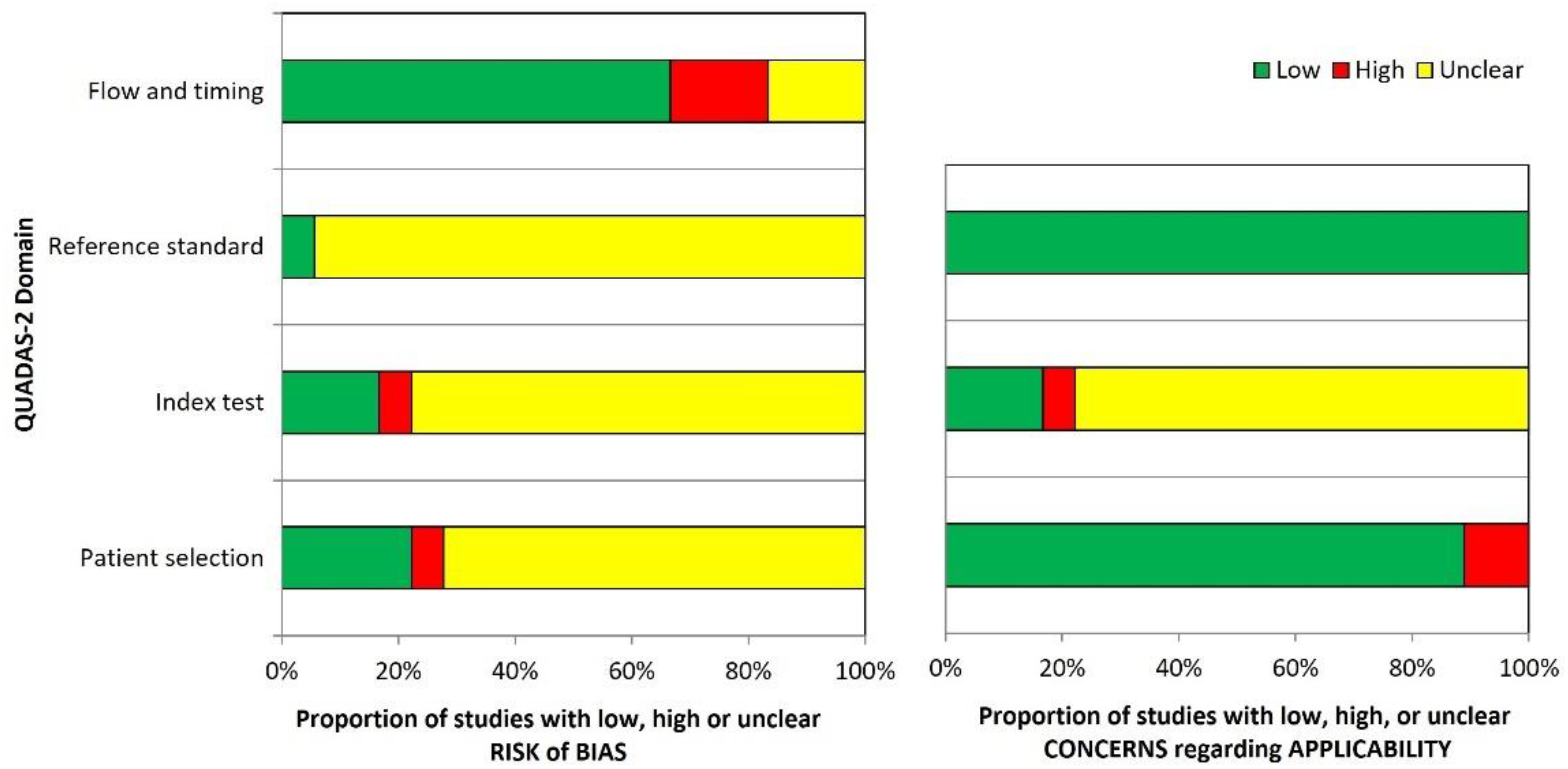
QUADAS-2 risk of bias and applicability concerns – graphical summary showing the review authors’ judgment about each domain as percentages across 18 test evaluations reported in 17 included studies.

**Table 8:** QUADAS-2 risk of bias and applicability concerns summary – review authors’ judgment about each domain for 18 test evaluations reported in 17 included studies.

| Study identifier | Risk of bias |  |  |  | Applicability concerns |  |  |
| --- | --- | --- | --- | --- | --- | --- | --- |
|  | Patient selection | Index test | Reference standard | Flow and timing | Patient selection | Index test | Reference standard |
| **Akingba** | unclear | unclear | unclear | low | low | unclear | low |
| **Bianco** | unclear | unclear | unclear | unclear | low | unclear | low |
| **Drevinek (IT1)** | unclear | unclear | unclear | low | low | unclear | low |
| **Drevinek (IT2)** | unclear | low | unclear | low | low | unclear | low |
| **Gonzalez-D.** | unclear | unclear | unclear | low | high | unclear | low |
| **Homza** | low | unclear | unclear | low | low | unclear | low |
| **Kiyasu** | unclear | unclear | unclear | low | low | unclear | low |
| **L’Huillier** | low | low | unclear | high | low | low | low |
| **Möckel** | low | low | low | low | low | unclear | low |
| **Pilarowski** | unclear | unclear | unclear | low | low | unclear | low |
| **Pollock a** | unclear | unclear | unclear | high | low | unclear | low |
| **Pollock b** | unclear | unclear | unclear | high | low | low | low |
| **Prince-G.** | unclear | unclear | unclear | unclear | low | low | low |
| **Shah** | unclear | unclear | unclear | low | low | unclear | low |
| **Sood** | unclear | high | unclear | unclear | low | high | low |
| **Takeuchi** | unclear | unclear | unclear | low | low | unclear | low |
| **Torres** | low | unclear | unclear | low | low | unclear | low |
| **Villaverde** | high | unclear | unclear | low | high | unclear | low |

IT1: Index test 1; IT 2: Index test 2

Overall applicability concerns were high in three studies due to high concerns in either the patient selection or index test domain. Three studies were of low concern and the remaining 11 were rated unclear due to insufficient reporting in at least one domain.

### Results of individual studies

RT-PCR positivity rate, diagnostic sensitivity, diagnostic specificity as well as PPV and NPV (and their 95% CIs) of individual studies based on data from 2×2 contingency tables for paediatric populations are reported in Table 9 and Figure 3. The RT-PCR positivity rate, which corresponds to the SARS-CoV-2 prevalence in the sample population, varied between 4.1% and 50%, with a median of 14,5% over n=17 studies. The sensitivity and specificity ranged from 33.3% to 85.7% and 91.7% to 100%, respectively. PPV and NPV ranged from 60.0% to 98.7% and 73.3% to 98.9%, respectively.

**Figure 3:**
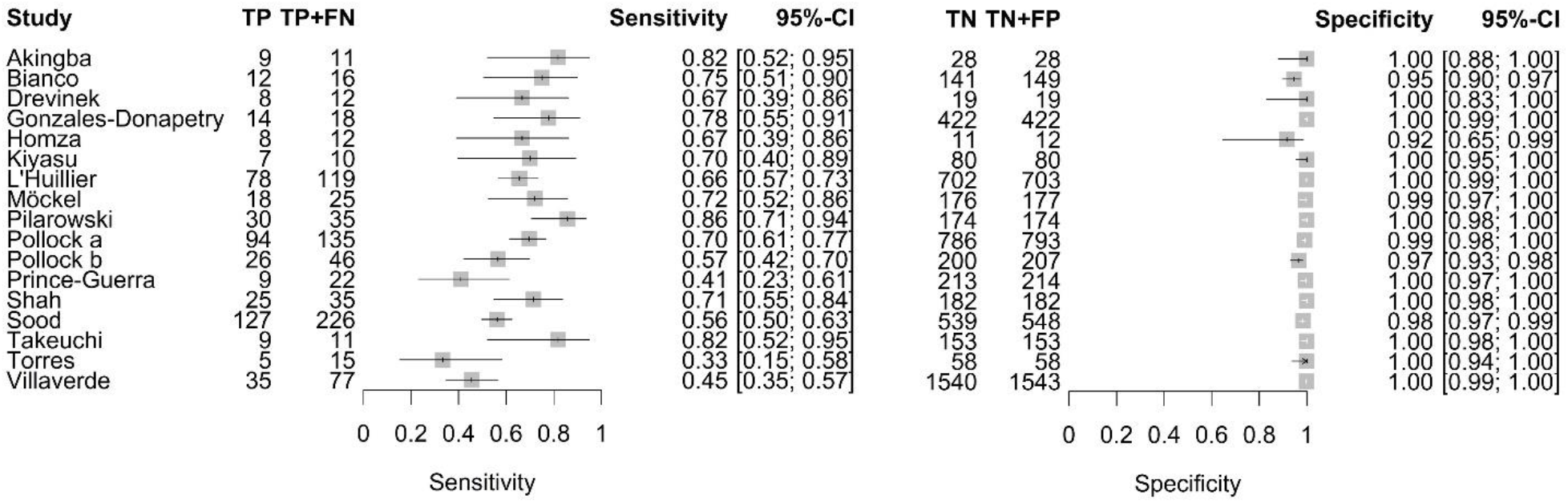
Forest plot of sensitivity and specificity of antigen tests in entire paediatric study populations irrespective of symptoms. The point estimates of sensitivity and specificity from each study (identified by name of first author) are shown as squares, the corresponding 95% confidence intervals (95% CIs) are represented as horizontal lines. TP: true positive, FN: false negative, TN: true negative, FP: false positive.

**Table 9:** Calculated RT-PCR positivity rate, sensitivity, specificity, positive predictive value (PPV) and negative predictive value (NPV) with 95% confidence interval (CI) for each included study based on the 2×2 contingency tables extracted for the entire pediatric study populations irrespective of symptom status.

| Study identifier | TP | FP | TN | FN | RT-PCR<br>positivity<br>rate | Sensitivity<br>(95% CI) | Specificity<br>(95% CI) | PPV<br>(95% CI) | NPV<br>(95% CI) |
| --- | --- | --- | --- | --- | --- | --- | --- | --- | --- |
| Akingba | 9 | 0 | 28 | 2 | 28.2 | 81.8 (52.3-94.9) | 100 (87.9-100) | 95.0 (65.6-99.5) | 91.9 (77.2-97.5) |
| Bianco | 12 | 8 | 141 | 4 | 9.7 | 75.0 (50.5-89.8) | 94.6 (89.8-97.3) | 60.0 (38.7-78.1) | 97.2 (93.1-98.9) |
| Drevinek (IT1) | 8 | 0 | 19 | 4 | 38.7 | 66.7 (39.1-86.2) | 100 (83.2-100) | 94.4 (62.9-99.4) | 81.2 (61.8-92.1) |
| Drevinek (IT2) | 8 | 0 | 19 | 4 | 38.7 | 66.7 (39.1-86.2) | 100 (83.2-100) | 94.4 (62.9-99.4) | 81.2 (61.8-92.1) |
| Gonzales-Donapetry | 14 | 0 | 422 | 4 | 4.1 | 77.8 (54.8-91.0) | 100 (99.1-100) | 96.7 (74.7-99.7) | 98.9 (97.4-99.6) |
| Homza | 8 | 1 | 11 | 4 | 50.0 | 66.7 (39.1-86.2) | 91.7 (64.6-98.5) | 88.9 (56.5-98.0) | 73.3 (48.0-89.1) |
| Kiyasu | 7 | 0 | 80 | 3 | 11.1 | 70.0 (39.7-89.2) | 100 (95.4-100) | 93.8 (59.8-99.3) | 95.8 (89.2-98.5) |
| L'Huillier | 78 | 1 | 702 | 41 | 14.5 | 65.5 (56.6-73.5) | 99.9 (99.2-100) | 98.7 (93.2-99.8) | 94.5 (92.6-95.9) |
| Möckel | 18 | 1 | 176 | 7 | 12.4 | 72.0 (52.4-85.7) | 99.4 (96.9-99.9) | 94.7 (75.4-99.1) | 96.2 (92.3-98.1) |
| Pilarowski | 30 | 0 | 174 | 5 | 16.7 | 85.7 (70.6-93.7) | 100 (97.8-100) | 98.4 (86.3-99.8) | 96.9 (93.3-98.6) |
| Pollock a | 94 | 7 | 786 | 41 | 14.5 | 69.6 (61.4-76.8) | 99.1 (98.2-99.6) | 93.1 (86.4-96.6) | 95.0 (93.3-96.3) |
| Pollock b | 26 | 7 | 200 | 20 | 18.2 | 56.5 (42.2-69.8) | 96.6 (93.2-98.4) | 78.8 (62.3-89.3) | 90.9 (86.4-94.0) |
| Prince-Guerra | 9 | 1 | 213 | 13 | 9.3 | 40.9 (23.3-61.3) | 99.5 (97.4-99.9) | 90.0 (59.6-98.2) | 94.2 (90.4-96.6) |
| Shah | 25 | 0 | 182 | 10 | 16.1 | 71.4 (54.9-83.7) | 100 (97.9-100) | 98.1 (84.0-99.8) | 94.6 (90.4-97.0) |
| Sood | 127 | 9 | 539 | 99 | 29.2 | 56.2 (49.7-62.5) | 98.4 (96.9-99.1) | 93.4 (87.9-96.5) | 84.5 (81.5-87.1) |
| Takeuchi | 9 | 0 | 153 | 2 | 6.7 | 81.8 (52.3-94.9) | 100 (97.6-100) | 95.0 (65.6-99.5) | 98.4 (95.0-99.5) |
| Torres | 5 | 0 | 58 | 10 | 20.5 | 33.3 (15.2-58.3) | 100 (93.8-100) | 91.7 (51.7-99.1) | 84.8 (74.5-91.4) |
| Villaverde | 35 | 3 | 1540 | 42 | 4.8 | 45.5 (34.8-56.5) | 99.8 (99.4-99.9) | 92.1 (79.2-97.3) | 97.3 (96.4-98.0) |

TP: true positive, FP: false positive, TN: true negative, FN: false negative
RT-PCR: reverse transcription-polymerase chain reaction
IT1: Index test 1; IT 2: Index test 2

For individual studies, separate analyses for subgroups based on symptom status are reported in Tables 10-12 and Figure 4. Here, populations were defined as symptomatic or asymptomatic if at least 80% of paediatric study participants were reported as being symptomatic or asymptomatic, respectively. Mixed populations refer to populations with no predominant symptom status. RT-PCR positivity rates of the primary analysis population and the different subgroups based on symptom status are presented in Figure 5. Two studies [51,54] were performed in high-prevalence populations with RT-PCR positivity rates of 38.7% and 50.0%, respectively. The median RT-PCR positivity rate was 13.2% (n=10 test evaluations) in asymptomatic populations, 13.8% in mixed populations (n=3 test evaluations), and 25.7% in symptomatic populations (n=13 test evaluations). Thus, we observed a slight trend in the RT-PCR positivity rate with respect to the proportion of symptomatic subjects. Although the bivariate meta-analysis might be influenced by the difference in the prevalence [73], we did not directly analyse the impact of the prevalence within the bivariate model. Instead, we performed a subgroup analysis with respect to symptom status (see below).

**Figure 4:**
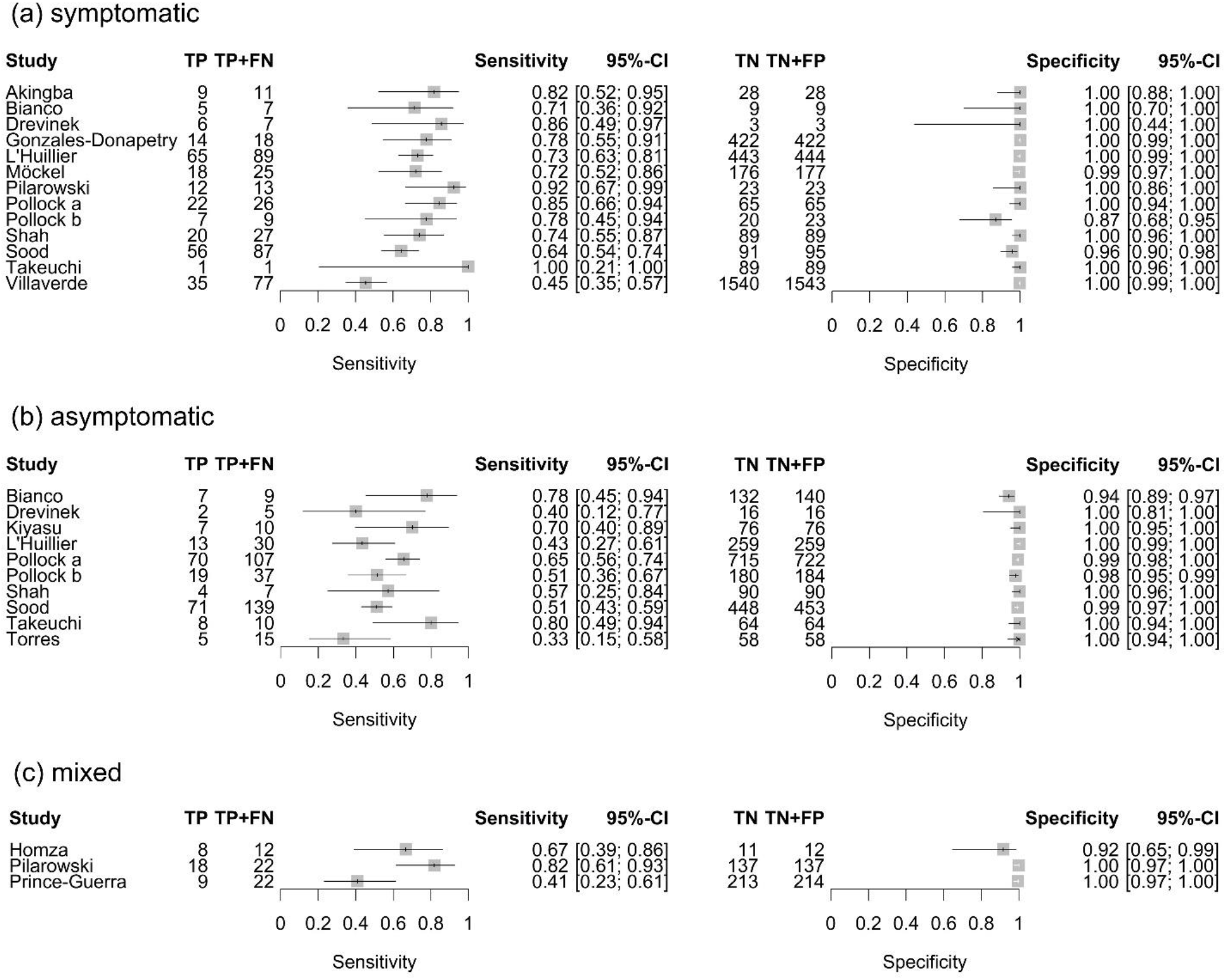
Forest plot of sensitivity and specificity of antigen tests in (a) symptomatic, (b) asymptomatic, and (c) mixed paediatric study populations. The point estimates of sensitivity and specificity from each study (identified by name of first author) are shown as squares, the corresponding 95% confidence intervals (95% CIs) are represented as horizontal lines. TP: true positive, FN: false negative, TN: true negative, FP: false positive.

**Figure 5:**
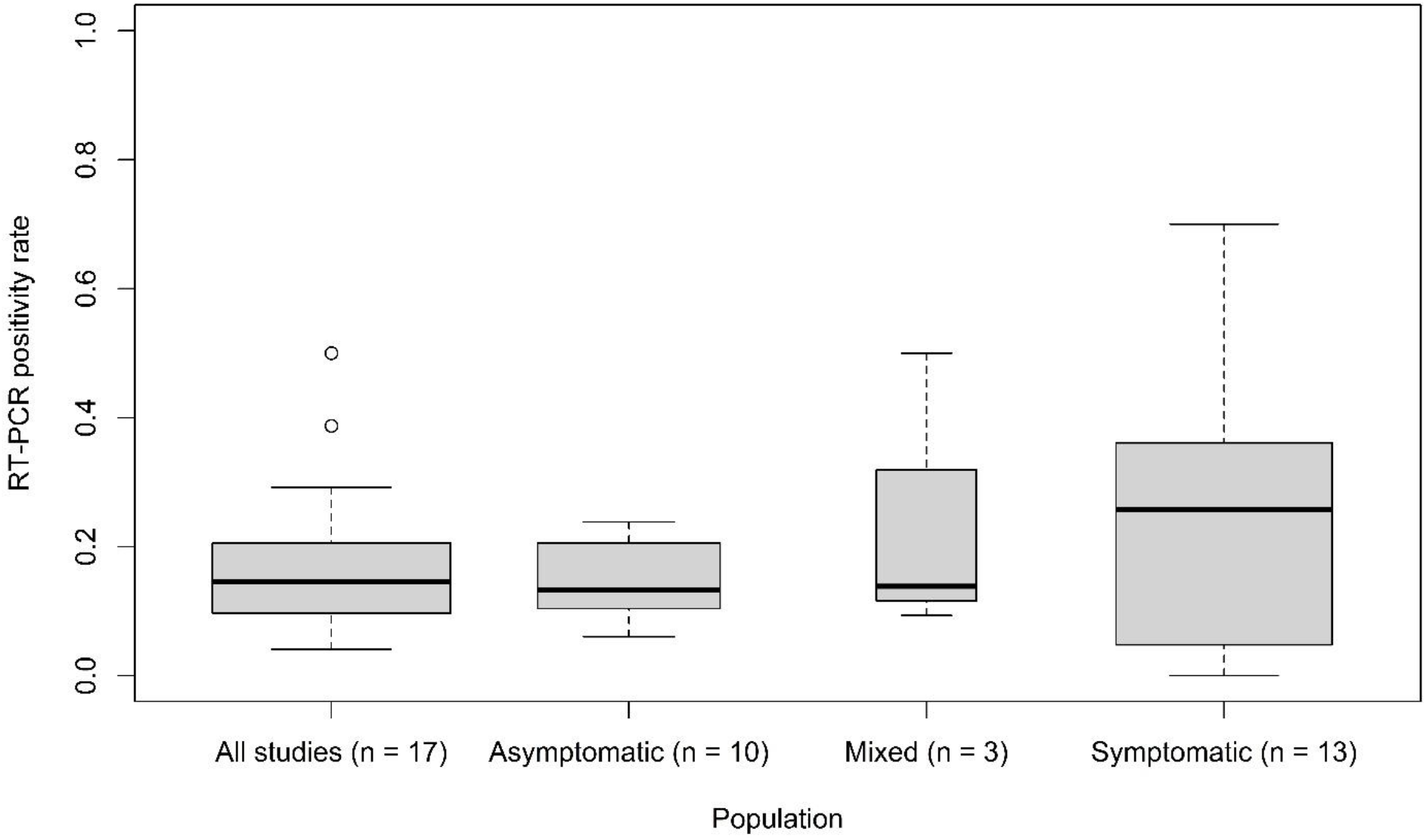
Box and whisker plots of the RT-PCR positivity rates in entire paediatric study populations irrespective of symptoms and in asymptomatic, mixed, and symptomatic paediatric study populations. The box is drawn from the first to the third quartile, defined via hinges. The bold horizontal line in the box denotes the median. The whiskers extend out of the box by 1.5 times the difference between the hinges. Outliers are plotted as small circle. The width of the box corresponds to the number of considered studies.

**Table 10:** Calculated RT-PCR positivity rate, sensitivity, specificity, positive predictive value (PPV) and negative predictive value (NPV) with 95% confidence interval (CI) for each included study based on the 2×2 contingency tables extracted for *symptomatic* pediatric study populations.

| Study identifier | TP | FP | TN | FN | RT-PCR<br>positivity<br>rate | Sensitivity<br>(95% CI) | Specificity<br>(95% CI) | PPV<br>(95% CI) | NPV<br>(95% CI) |
| --- | --- | --- | --- | --- | --- | --- | --- | --- | --- |
| Akingba | 9 | 0 | 28 | 2 | 28.2 | 81.8 (52.3-94.9) | 100 (87.9-100) | 95.0 (65.6-99.5) | 91.9 (77.2-97.5) |
| Bianco | 5 | 0 | 9 | 2 | 43.8 | 71.4 (35.9-91.8) | 100 (70.1-100) | 91.7 (51.7-99.1) | 79.2 (50.9-93.3) |
| Drevinek (IT1) | 6 | 0 | 3 | 1 | 70.0 | 85.7 (48.7-97.4) | 100 (43.9-100) | 92.9 (56.1-99.2) | 70.0 (29.9-92.7) |
| Drevinek (IT2) | 6 | 0 | 3 | 1 | 70.0 | 85.7 (48.7-97.4) | 100 (43.9-100) | 92.9 (56.1-99.2) | 70.0 (29.9-92.7) |
| Gonzales-Donapetry | 14 | 0 | 422 | 4 | 4.1 | 77.8 (54.8-91.0) | 100 (99.1-100) | 96.7 (74.7-99.7) | 98.9 (97.4-99.6) |
| L'Huillier | 65 | 1 | 443 | 24 | 16.7 | 73.0 (63.0-81.2) | 99.8 (98.7-100) | 98.5 (91.9-99.7) | 94.9 (92.5-96.5) |
| Möckel | 18 | 1 | 176 | 7 | 12.4 | 72.0 (52.4-85.7) | 99.4 (96.9-99.9) | 94.7 (75.4-99.1) | 96.2 (92.3-98.1) |
| Pilarowski | 12 | 0 | 23 | 1 | 36.1 | 92.3 (66.7-98.6) | 100 (85.7-100) | 96.2 (71.7-99.6) | 94.0 (77.7-98.6) |
| Pollock a | 22 | 0 | 65 | 4 | 28.6 | 84.6 (66.5-93.8) | 100 (94.4-100) | 97.8 (82.2-99.8) | 93.6 (85.3-97.3) |
| Pollock b | 7 | 3 | 20 | 2 | 28.1 | 77.8 (45.3-93.7) | 87 (67.9-95.5) | 70.0 (39.7-89.2) | 90.9 (72.2-97.5) |
| Shah | 20 | 0 | 89 | 7 | 23.3 | 74.1 (55.3-86.8) | 100 (95.9-100) | 97.6 (80.8-99.8) | 92.3 (85.2-96.1) |
| Sood | 56 | 4 | 91 | 31 | 47.8 | 64.4 (53.9-73.6) | 95.8 (89.7-98.4) | 93.3 (84.1-97.4) | 74.6 (66.2-81.5) |
| Takeuchi | 1 | 0 | 89 | 0 | 1.1 | 100 (20.7-100) | 100 (95.9-100) | 75.0 (19.8-97.3) | 99.4 (94.9-99.9) |
| Villaverde | 35 | 3 | 1540 | 42 | 4.8 | 45.5 (34.8-56.5) | 99.8 (99.4-99.9) | 92.1 (79.2-97.3) | 97.3 (96.4-98.0) |

TP: true positive, FP: false positive, TN: true negative, FN: false negative
RT-PCR: reverse transcription-polymerase chain reaction
IT1: Index test 1; IT 2: Index test 2

**Table 11:** Calculated RT-PCR positivity rate, sensitivity, specificity, positive predictive value (PPV) and negative predictive value (NPV) with 95% confidence interval (CI) for each included study based on the 2×2 contingency tables extracted for *asymptomatic* pediatric study populations.

| Study identifier | TP | FP | TN | FN | RT-PCR<br>positivity<br>rate | Sensitivity<br>(95% CI) | Specificity<br>(95% CI) | PPV<br>(95% CI) | NPV<br>(95% CI) |
| --- | --- | --- | --- | --- | --- | --- | --- | --- | --- |
| **Bianco** | 7 | 8 | 132 | 2 | 6.0 | 77.8 (45.3-93.7) | 94.3 (89.1-97.1) | 46.7 (24.8-69.9) | 98.5 (94.7-99.6) |
| **Drevinek (IT1)** | 2 | 0 | 16 | 3 | 23.8 | 40.0 (11.8-76.9) | 100 (80.6-100) | 83.3 (31.0-98.2) | 82.5 (61.1-93.4) |
| **Drevinek (IT2)** | 2 | 0 | 16 | 3 | 23.8 | 40.0 (11.8-76.9) | 100 (80.6-100) | 83.3 (31.0-98.2) | 82.5 (61.1-93.4) |
| **Kiyasu** | 7 | 0 | 76 | 3 | 11.6 | 70.0 (39.7-89.2) | 100 (95.2-100) | 93.8 (59.8-99.3) | 95.6 (88.7-98.4) |
| **L'Huillier** | 13 | 0 | 259 | 17 | 10.4 | 43.3 (27.4-60.8) | 100 (98.5-100) | 96.4 (73.2-99.6) | 93.7 (90.2-96.0) |
| **Pollock a** | 70 | 7 | 715 | 37 | 12.9 | 65.4 (56.0-73.8) | 99.0 (98.0-99.5) | 90.9 (82.4-95.5) | 95.1 (93.3-96.4) |
| **Pollock b** | 19 | 4 | 180 | 18 | 16.7 | 51.4 (35.9-66.6) | 97.8 (94.5-99.2) | 82.6 (62.9-93.0) | 90.9 (86.1-94.2) |
| **Shah** | 4 | 0 | 90 | 3 | 7.2 | 57.1 (25.0-84.2) | 100 (95.9-100) | 90.0 (46.3-99.0) | 96.3 (90.3-98.6) |
| **Sood** | 71 | 5 | 448 | 68 | 23.5 | 51.1 (42.9-59.2) | 98.9 (97.4-99.5) | 93.4 (85.5-97.2) | 86.8 (83.6-89.5) |
| **Takeuchi** | 8 | 0 | 64 | 2 | 13.5 | 80.0 (49.0-94.3) | 100 (94.3-100) | 94.4 (62.9-99.4) | 96.3 (88.7-98.8) |
| **Torres** | 5 | 0 | 58 | 10 | 20.5 | 33.3 (15.2-58.3) | 100 (93.8-100) | 91.7 (51.7-99.1) | 84.8 (74.5-91.4) |

TP: true positive, FP: false positive, TN: true negative, FN: false negative
RT-PCR: reverse transcription-polymerase chain reaction
IT1: Index test 1; IT 2: Index test 2

**Table 12:** Calculated RT-PCR positivity rate, sensitivity, specificity, positive predictive value (PPV) and negative predictive value (NPV) with 95% confidence interval (CI) for each included study based on the 2×2 contingency tables extracted for *mixed* pediatric study populations.

| Study identifier | TP | FP | TN | FN | RT-PCR<br>positivity<br>rate | Sensitivity<br>(95% CI) | Specificity<br>(95% CI) | PPV<br>(95% CI) | NPV<br>(95% CI) |
| --- | --- | --- | --- | --- | --- | --- | --- | --- | --- |
| **Homza** | 8 | 1 | 11 | 4 | 50.0 | 66.7 (39.1-86.2) | 91.7 (64.6-98.5) | 88.9 (56.5-98.0) | 73.3 (48.0-89.1) |
| **Pilarowski** | 18 | 0 | 137 | 4 | 13.8 | 81.8 (61.5-92.7) | 100 (97.3-100) | 97.4 (79.1-99.7) | 96.8 (92.5-98.7) |
| **Prince-Guerra** | 9 | 1 | 213 | 13 | 9.3 | 40.9 (23.3-61.3) | 99.5 (97.4-99.9) | 90.0 (59.6-98.2) | 94.2 (90.4-96.6) |

TP: true positive, FP: false positive, TN: true negative, FN: false negative
RT-PCR: reverse transcription-polymerase chain reaction

### Synthesis of results

In our primary meta-analyses, we used data from 17 studies evaluating the diagnostic accuracy of antigen tests in paediatric participants. Estimated pooled sensitivity and specificity were 64.2% (95% CI: 57.4% to 70.5%) and 99.1% (95% CI: 98.2% to 99.5%), respectively. While the estimates for the sensitivity revealed high heterogeneity and thus justified the application of the bivariate model with random effects, the estimates for the specificity were limited to a small range, as shown in Figure 6. As a consequence, the estimated summary receiver operating characteristic (SROC) curve cannot be meaningfully interpreted. As pre-specified in the protocol, we performed subgroup analysis evaluating the diagnostic accuracy according to symptom status. Estimated pooled sensitivity and specificity in asymptomatic children was 56.2% (95% CI: 47.6% to 64.4%) and 98.6% (95% CI: 97.3% to 99.3%), respectively, based on data from 2439 asymptomatic children in 10 studies. Estimated pooled sensitivity and specificity in symptomatic children was 71.8% (95% CI: 63.6% to 78.8%) and 98.7% (95% CI: 96.6% to 99.5%), respectively, based on data from 3413 symptomatic children in 13 studies. Estimated pooled sensitivity and specificity in the mixed population of symptomatic and asymptomatic children from three studies including 419 children was 63.4% (95% CI: 37.3% to 83.5%) and 98.7% (95% CI: 90.8% to 99.8%), respectively. The corresponding SROC curves are shown in Figure 7. The likelihood ratio test (LRT) for differences between the three groups revealed a p-value of p_LRT_=0.066. Results for the other subgroup analyses did not show relevant differences in the pooled estimates (setting p_LRT_=0.400; index test sample type p_LRT_=0.303; reference standard sample type p_LRT_=0.723; RT-PCR Ct cut-off value p_LRT_=0.105; publication status p_LRT_=0.551; test type (most used) p_LRT_=0.146). Due to insufficient data, we did not perform subgroup analysis with respect to test type (antigen vs. molecular) and end-user (layperson (self-testing) vs. trained staff/health care worker). Except for one study [61,74] where the testing procedure involved (supervised) self-collection of samples by study participants, in all other studies, testing was conducted with trained staff and/or health care workers (if reported). Univariate meta-analysis with random effects for sensitivity and specificity in cases where only a few studies were included (mixed population of symptomatic and asymptomatic children) did not show remarkable differences to the bivariate analysis. The results of all bivariate meta-analyses are summarised in Table 15.

**Figure 6:**
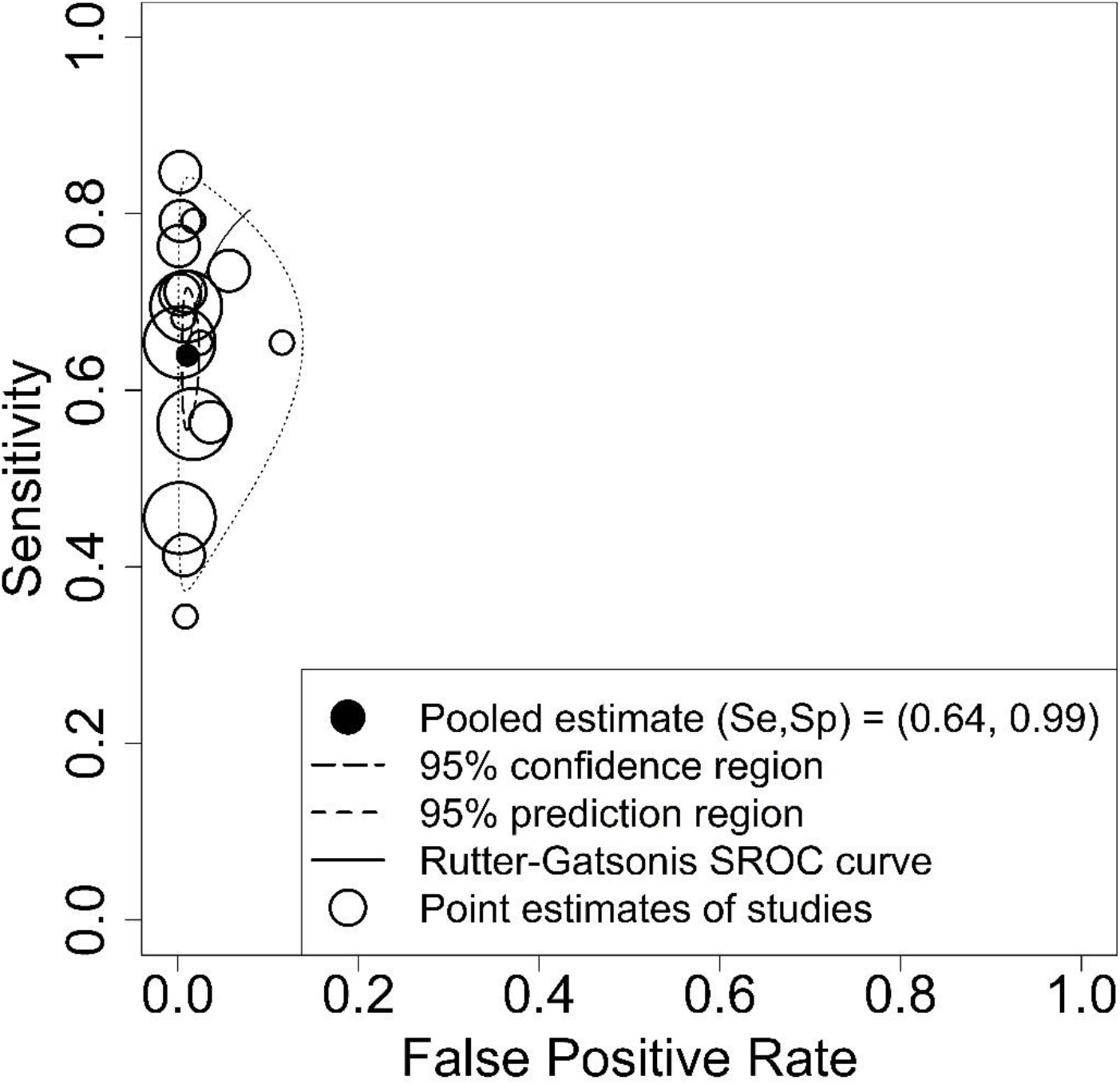
Summary receiver operating characteristic (SROC) plot of sensitivity and specificity of antigen tests for diagnosis of current SARS-CoV-2 infections in entire paediatric study populations irrespective of symptoms. Each circle represents the point estimate of an individual study, whereas the size of the circle correlates with the number of paediatric study participants (small circle: <100 participants, medium circle: between 100 and 500 participants, large circle: >500 participants). The pooled estimate (black dot) of the pair of sensitivity (Se) and specificity (Sp) is surrounded by its 95% confidence region (closed curve with short dashes) and prediction region (closed curve with long dashes). The estimation of the SROC curve is based on the bivariate approach by Rutter and Gatsonis [29].

**Figure 7:**
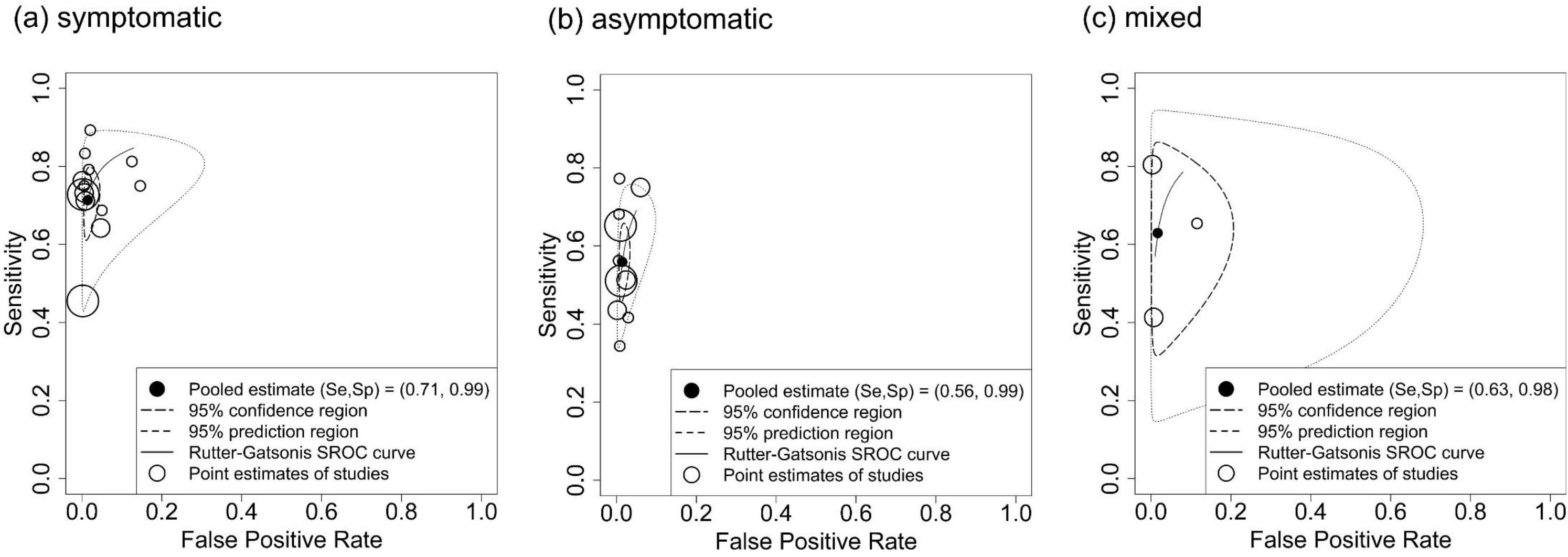
Summary receiver operating characteristic (SROC) plot of sensitivity and specificity of antigen tests for diagnosis of current SARS-CoV-2 infections in (a) symptomatic, (b) asymptomatic, and (c) mixed symptomatic and asymptomatic paediatric study populations. Each circle represents the point estimate of an individual study, whereas the size of the circle correlates with the number of paediatric study participants (small circle: <100 participants, medium circle: between 100 and 500 participants, large circle: >500 participants). The pooled estimate (black dot) of the pair of sensitivity (Se) and specificity (Sp) is surrounded by its 95% confidence region (closed curve with short dashes) and prediction region (closed curve with long dashes). The estimation of the SROC curve is based on the bivariate approach by Rutter and Gatsonis [29].

## DISCUSSION

To our knowledge, this is the first systematic review that focused on evaluating the diagnostic accuracy of rapid point-of-care tests for current SARS-CoV-2 infections in paediatric populations. Our current review comprises 17 studies with 18 evaluations of eight different antigen tests in children, whereas comprehensive author queries allowed us to include eight studies that did not provide sufficient data on paediatric study participants in their original study publication. We did not identify any evaluations of molecular-based tests that met our inclusion criteria confirming the current dominant role of antigen tests for rapid point-of-care usage.

Sensitivity estimates of antigen tests varied broadly between studies and were substantially lower as reported by manufacturers. Less variation and only minor discrepancies to performance claims by manufacturers were observed for specificity estimates across studies. Taking into account test-specific pooled results, no test included in this review fully satisfied the minimum performance requirements as recommended by the WHO [9] (minimum sensitivity of ≥ 80% and minimum specificity of ≥ 97%), the US FDA [10] (minimum sensitivity of ≥ 80%, whereas a lower bound of the two-sided 95% CI above 70% is required for over-the-counter use self-tests [75]) or the Medicines and Healthcare products Regulatory Agency (MHRA) in the United Kingdom (UK) [76] (minimum acceptable sensitivity of ≥ 80% with two-sided 95% CI entirely above 70% and minimum acceptable specificity of 95% with two-sided 95% CI entirely above 90%).

While specificities were similar high in symptomatic (98.7% with 95% CI: 96.6% to 99.5%) and asymptomatic (98.6% with 95% CI: 97.3% to 99.3%) populations, we observed a drop in sensitivity by about 15 percentage points in asymptomatic populations (56.2% with 95% CI: 47.6% to 64.4%) compared to symptomatic populations (71.8% with 95% CI 63.6% to 78.8%). The better performance in symptomatic populations might be explained by changes in the viral load over the course of infection and the timing of the test: most symptomatic individuals were tested within seven days of symptom onset in contrast to asymptomatic individuals with more variable disease onset, including individuals in the early (pre-symptomatic) or late stages of infection when viral loads are relatively low [77]. Further, since test performance can also be influenced by prevalence, one should note that median prevalence in symptomatic study populations was about 12 percentage points higher than in asymptomatic study populations.

As expected, the sensitivity increased when the positivity threshold of the reference standard was set to a lower Ct cut-off value of 30 or 25. However, such analyses should not be over-interpreted since Ct values are not standardized across systems or laboratories, making it difficult to directly compare results between different studies. Furthermore, while the Ct value from RT-PCR is a strong indicator of viral load, there is no specific cut-off viral load which allows distinguishing individuals as being infectious or not. As shown in Table 13 and Table 14, an increase in sensitivity comes at the cost of a decrease in specificity as antigen tests also identify individuals with moderate or low viral loads, who would then be considered as false positives.

**Table 13:** Calculated RT-PCR positivity rate, sensitivity, specificity, positive predictive value (PPV) and negative predictive value (NPV) with 95% confidence interval (CI) for each included study based on the 2×2 contingency tables extracted for entire pediatric study populations irrespective of symptom status when the RT-PCR cycle threshold cut-off value is set to 30.

| Study identifier | TP | FP | TN | FN | RT-PCR<br>positivity<br>rate | Sensitivity<br>(95% CI) | Specificity<br>(95% CI) | PPV<br>(95% CI) | NPV<br>(95% CI) |
| --- | --- | --- | --- | --- | --- | --- | --- | --- | --- |
| **Akingba** | 9 | 0 | 30 | 0 | 23.1 | 100 (70.1-100) | 100 (88.6-100) | 95.0 (65.6-99.5) | 98.4 (86.3-99.8) |
| **Pilarowski** | 24 | 6 | 179 | 0 | 11.5 | 100 (86.2-100) | 96.8 (93.1-98.5) | 79.0 (61.9-89.7) | 99.7 (97.4-100) |
| **Pollock a** | 84 | 17 | 818 | 9 | 10.0 | 90.3 (82.6-94.8) | 98.0 (96.8-98.7) | 83.2 (74.7-89.2) | 98.9 (97.9-99.4) |
| **Pollock b** | 24 | 9 | 215 | 5 | 11.5 | 82.8 (65.5-92.4) | 96.0 (92.5-97.9) | 72.7 (55.8-84.9) | 97.7 (94.8-99.0) |
| **Sood** | 42 | 91 | 625 | 11 | 6.9 | 79.2 (66.5-88.0) | 87.3 (84.7-89.5) | 31.6 (24.3-39.9) | 98.3 (96.9-99.0) |
| **Torres** | 5 | 0 | 60 | 8 | 17.8 | 38.5 (17.7-64.5) | 100 (94.0-100) | 91.7 (51.7-99.1) | 87.7 (77.9-93.5) |

TP: true positive, FP: false positive, TN: true negative, FN: false negative
RT-PCR: reverse transcription-polymerase chain reaction

**Table 14:** Calculated RT-PCR positivity rate, sensitivity, specificity, positive predictive value (PPV) and negative predictive value (NPV) with 95% confidence interval (CI) for each included study based on the 2×2 contingency tables extracted for entire pediatric study populations irrespective of symptom status when the RT-PCR cycle threshold cut-off value is set to 25.

| Study identifier | TP | FP | TN | FN | RT-PCR<br>positivity<br>rate | Sensitivity<br>(95% CI) | Specificity<br>(95% CI) | PPV<br>(95% CI) | NPV<br>(95% CI) |
| --- | --- | --- | --- | --- | --- | --- | --- | --- | --- |
| **Akingba** | 4 | 5 | 30 | 0 | 10.3 | 100 (51.0-100) | 85.7 (70.6-93.7) | 45.0 (20.1-72.6) | 98.4 (86.3-99.8) |
| **Pollock a** | 61 | 40 | 826 | 1 | 6.7 | 98.4 (91.4-99.7) | 95.4 (93.8-96.6) | 60.4 (50.6-69.4) | 99.9 (99.3-100) |
| **Pollock b** | 20 | 13 | 220 | 0 | 7.9 | 100 (83.9-100) | 94.4 (90.7-96.7) | 60.3 (43.6-74.9) | 99.8 (97.9-100) |
| **Sood** | 15 | 118 | 635 | 1 | 2.1 | 93.8 (71.7-98.9) | 84.3 (81.6-86.8) | 11.3 (7.0-17.8) | 99.8 (99.1-100) |
| **Torres** | 4 | 1 | 65 | 3 | 9.6 | 57.1 (25.0-84.2) | 98.5 (91.9-99.7) | 80.0 (37.5-96.4) | 95.6 (87.8-98.5) |

TP: true positive, FP: false positive, TN: true negative, FN: false negative
RT-PCR: reverse transcription-polymerase chain reaction

**Table 15:** Results of the bivariate meta-analyses: pooled sensitivity and specificity with 95% confidence interval (CI).

|  | **Sensitivity**<br>**(95% CI)** | **Specificity**<br>**(95% CI)** |
| --- | --- | --- |
| **All studies** | 64.2 (57.4-70.5) | 99.1 (98.2-99.5) |
| **Symptom status: symptomatic** | 71.8 (63.6-78.8) | 98.7 (96.6-99.5) |
| **Symptom status: asymptomatic** | 56.2 (47.6-64.4) | 98.6 (97.3-99.3) |
| **Symptom status: mixed** | 63.4 (37.3-83.5) | 98.7 (90.8-99.8) |
| **Setting: community testing site** | 64.1 (54.7-72.6) | 98.7 (97.6-99.3) |
| **Setting: hospital test center / emergency department** | 64.1 (53.8-73.2) | 99.4 (98.2-99.8) |
| **Sample type (index test): nasopharyngeal** | 64.3 (54.7-73.0) | 99.4 (98.5-99.8) |
| **Sample type (index test): not nasopharyngeal** | 64.6 (54.4-73.7) | 98.5 (96.7-99.3) |
| **Sample type (reference standard): nasopharyngeal** | 65.4 (56.3-73.5) | 99.1 (97.7-99.7) |
| **Sample type (reference standard): not nasopharyngeal** | 64.2 (53.1-74.0) | 98.9 (97.6-99.5) |
| **RT-PCR Ct cut-off value = 25** | 92.4 (72.7-98.2) | 92.7 (85.4-96.5) |
| **RT-PCR Ct cut-off value = 30** | 83.3 (63.9-93.4) | 96.1 (91.8-98.2) |
| **Publication status: preprint** | 63.2 (55.6-70.3) | 98.9 (95.9-99.7) |
| **Publication status: peer-reviewed** | 64.3 (54.8-72.7) | 99.1 (98.1-99.6) |
| **Test type (most used): Panbio** | 60.8 (46.9-73.1) | 99.7 (99.2-99.9) |
| **Test type (most used): BinaxNOW** | 66.3 (53.0-77.5) | 99.1 (98.2-99.5) |

RT-PCR: reverse transcription-polymerase chain reaction
Ct: cycle threshold

Despite some methodological differences (such as the stringency of inclusion criteria) and neglecting differences between included studies (e.g. settings), the findings of our review are similar to those in the recent Cochrane Review by Dinnes et al. [8]. These similarities between paediatric and adult populations might be explained by the findings by Jones et al. [78], who only identified minor differences in viral loads across age groups in a comprehensive analysis of more than 25,000 individuals who tested positive for SARS-CoV-2 by RT-PCR.

For the current version of our review, publication bias is not considered as relevant due to the novelty of the topic. Non-publication of studies, in particular underreporting of studies with unfavourable outcomes, may introduce bias and threaten the validity of systematic reviews. It is now commonly expected to publish results of clinical studies within 12 months of completion [79–81]. No study included in our review was published before November 2020. All four completed studies that were identified through searching study registries were completed within the last nine months. Of note is that for all but one study [39] included in our review no entry in a study registry was reported.

Despite the roll-out of vaccines, testing continues to be a key to pandemic control. As experts predict new surges of infections in autumn, early identification of outbreaks will remain vital for controlling the spread of SARS-CoV-2, particularly in populations with low vaccination rates. Consequently, multi-layered mitigation strategies will continue involving screening testing of children in schools and kindergarten to avoid further closures. The high specificity of antigen tests and the corresponding PPVs calculated for the paediatric study populations suggest that antigen testing might be a valuable tool to rapidly identify children with SARS-CoV-2 infection in moderate to high prevalence settings. However, at the same time, it is important to raise awareness that antigen tests should not be used to rule out SARS-CoV-2 infection (or infectiousness) due to their limited sensitivity. Whether increasing the frequency of antigen-based testing leads to an improved overall diagnostic accuracy that allows to effectively reduce transmission of SARS-CoV-2, has yet to be demonstrated in practice [82]. The latter two aspects and the urgent need for high quality screening tests likely led to the recent publication of a new target product profile by the MHRA in the UK [83], which includes increased performance requirements for self-tests to be used in national testing programs that aim at detecting current SARS-CoV-2 infections in individuals without symptoms. Here, the minimum acceptable sensitivity for tests to “rule out” a current infection is ≥97% with two-sided 95% CI entirely above 95%. The minimum acceptable specificity is ≥99.5% with two-sided 95% CI entirely above 97%. Further, it is stated that performance claims of repeated testing strategies require adequate clinical evidence rather than evidence from modelling studies only.

Other screening testing methods such as molecular-based pool testing, which involves RT-PCR testing of pooled samples and so-called deconvolution testing of individuals belonging to pools tested positive are currently under investigation [84–86] and may complement mere antigen testing, in particular in low prevalence settings. Further, novel tests, for example, lateral flow tests based on clustered regularly interspaced short palindromic repeat (CRISPR), hold great promise for a highly sensitive direct detection of SARS-CoV-2 [87] but have yet to demonstrate their value upon market access.

### Limitations

The results presented in this review should be considered in light of the following limitations:

1. As visualized in the QUADAS-2 summary graph, the majority of studies were at unclear risk of bias in at least three of the four domains of the QUADAS-2 tool because of poor reporting by authors. Therefore, the results reported in this review should be interpreted with caution. The issue of insufficient reporting has been addressed previously and led to the publication of comprehensive guidance to authors evaluating the clinical performance of tests for SARS-CoV-2 [88].
2. We acknowledge that infectiousness as target condition is of higher practical relevance than current SARS-CoV-2 infection, which was chosen as the target condition of this review. While RT-PCR as the corresponding reference standard is a highly sensitive method that is used to detect the presence of viral ribonucleic acid (RNA) in a specimen, this does not necessarily indicate that infectious virus is present. Therefore, the actual transmission risk from individuals who tested RT-PCR positive remains unknown. Testing for infectiousness would allow to identify (and isolate) exclusively individuals who could pass the virus to others. However, while there have been attempts to use viral load (estimated from Ct values) or virus viability in cell culture as a proxy to determine infectious individuals, up to now, there is no adequate reference standard for infectiousness [82].
3. Screening testing programs of children in schools and kindergarten using antigen tests have been implemented in many countries. Specific aspects that define these use cases, such as sample collection in toddlers by laypersons or self-testing in schools performed by children, are likely to influence the real-life test performance but were not addressed in any of the studies included in our review. Furthermore, one should keep in mind that diagnostic accuracy is only one factor affecting the effectiveness of testing programs [89].
4. Only eight different antigen tests were included in our review, and four of these were only evaluated in one study each. Thus, the performance of most antigen tests under real-life conditions remains unknown. While the number of antigen tests is still small in the US due to more rigorous approval processes, the situation in Europe is different, since the current regulatory framework for market access in the Europe Union (EU) [90] allows self-declaration of regulatory conformity of SARS-CoV-2 antigen tests for professional use by manufacturers without the involvement of a third party. As of July 6, 2021, more than 500 antigen tests gained market access in the EU [91]. A recent laboratory-based study evaluated the sensitivity of 122 of these antigen tests using common SARS-CoV-2 specimens with varying viral concentrations [92]. Even under such ideal conditions, a wide range of sensitivities was observed, whereas 26 tests missed the study’s sensitivity criteria of 75% for specimens with high SARS-CoV-2 concentrations of around 10^6^ SARS-CoV-2 RNA/ml and higher corresponding to a Ct value of <25.
5. Diagnostic accuracy estimates reported in this review may not apply to future variants of SARS-CoV-2. Considering the reported recruitment periods, most individuals identified as RT-PCR positive were likely infected with the wild-type of SARS-CoV-2. The impact of new emerging variants of SARS-CoV-2 on the diagnostic accuracy of SARS-CoV-2 tests is unknown and requires thorough investigation.

## CONCLUSION

Our findings confirm the urgent need for independent validations of rapid tests in paediatric populations under real-life conditions. Estimates of sensitivity and specificity and their 95% CIs from the bivariate meta-analyses indicate a subpar real-life performance of current antigen tests below the minimum performance criteria set by WHO, the US FDA, or the MHRA in the UK. Policymakers should especially be aware of the limited diagnostic sensitivity of current antigen tests and should require performance data from diagnostic accuracy studies in the intended use setting prior to the broad implementation of testing programs. Results presented in this systematic review should be interpreted with caution since risk of bias was predominantly judged as unclear due to poor reporting. We encourage study authors to put more emphasis on the reporting of their studies as stronger adherence to reporting standards would highly increase the value of their research. Future studies should investigate whether screening testing in paediatric populations is indeed effective in reducing the spread of SARS-CoV-2 and whether the benefits outweigh the harms.

## Supporting information

PRISMA Checklists (DTA for Abstracts, DTA, S)

Appendix 1: Search strategies

Appendix 2: QUADAS-2 Guidance

Appendix 3: List of excluded studies

## Data Availability

Any data supporting the findings of this study are available within the article or its supplementary files.

## ACKNOWLEDGMENTS

We thank Dr. Siw Waffenschmidt (IQWiG, Germany) for her valuable guidance during the development of the search strategies, the peer-review of the final search strategy, and the literature screening technical support. Further, we are grateful to all study authors who replied to our author queries and who provided us with separate data on the paediatric study participants included in their studies.

## DATA SHARING STATEMENT

The data supporting the findings of this review are available within the article or its appendices The R code used for this review is available from the corresponding author (NFR) on request.

