## Supplementary material for "Diagnostic accuracy of rapid point-of-care tests for diagnosis of current SARS-CoV-2 infections in children: A systematic review and meta-analysis": PRISMA Checklists (DTA for Abstracts, DTA, S)

### a) PRISMA-DTA for Abstracts Checklist

| Section/topic | # | PRISMA-DTA for Abstracts Checklist item | Reported on page # |
|---|---|---|---|
| **TITLE and PURPOSE** | | | |
| **Title** | 1 | Identify the report as a systematic review (+/- meta-analysis) of diagnostic test accuracy (DTA) studies. | 1 |
| **Objectives** | 2 | Indicate the research question, including components such as participants, index test, and target conditions. | 3 |
| **METHODS** | | | |
| **Eligibility criteria** | 3 | Include study characteristics used as criteria for eligibility. | 3 |
| **Information sources** | 4 | List the key databases searched and the search dates. | 3 |
| **Risk of bias & applicability** | 5 | Indicate the methods of assessing risk of bias and applicability. | 3 |
| **Synthesis of results** | A1 | Indicate the methods for the data synthesis. | 3 |
| **RESULTS** | | | |
| **Included studies** | 6 | Indicate the number and type of included studies and the participants and relevant characteristics of the studies (including the reference standard). | 3 |
| **Synthesis of results** | 7 | Include the results for the analysis of diagnostic accuracy, preferably indicating the number of studies and participants. Describe test accuracy including variability; if meta-analysis was done, include summary results and confidence intervals. | 3 |
| **DISCUSSION** | | | |
| **Strengths and limitations** | 9 | Provide a brief summary of the strengths and limitations of the evidence | 3 |
| **Interpretation** | 10 | Provide a general interpretation of the results and the important implications. | 3 |
| **OTHER** | | | |
| **Funding** | 11 | Indicate the primary source of funding for the review. | 2 |
| **Registration** | 12 | Provide the registration number and the registry name | 3 |

Adapted From: McInnes MDF, Moher D, Thombs BD, McGrath TA, Bossuyt PM, The PRISMA-DTA Group (2018). Preferred Reporting Items for a Systematic Review and Meta-analysis of Diagnostic Test Accuracy Studies: The PRISMA-DTA Statement. JAMA. 2018 Jan 23;319(4):388-396. doi: 10.1001/jama.2017.19163.

b) PRISMA-DTA Checklist

| Section/topic | # | PRISMA-DTA Checklist Item | Reported on page # |
|---|---|---|---|
| **TITLE / ABSTRACT** | | | |
| **Title** | 1 | Identify the report as a systematic review (+/- meta-analysis) of diagnostic test accuracy (DTA) studies. | 1 |
| **Abstract** | 2 | Abstract: See PRISMA-DTA for abstracts. | 3 |
| **INTRODUCTION** | | | |
| **Rationale** | 3 | Describe the rationale for the review in the context of what is already known. | 4, 5 |
| **Clinical role of index test** | D1 | State the scientific and clinical background, including the intended use and clinical role of the index test, and if applicable, the rationale for minimally acceptable test accuracy (or minimum difference in accuracy for comparative design). | 5, minimal performance requirements: 14, 15 |
| **Objectives** | 4 | Provide an explicit statement of question(s) being addressed in terms of participants, index test(s), and target condition(s). | 3 |
| **METHODS** | | | |
| **Protocol and registration** | 5 | Indicate if a review protocol exists, if and where it can be accessed (e.g., Web address), and, if available, provide registration information including registration number. | 3,6 |
| **Eligibility criteria** | 6 | Specify study characteristics (participants, setting, index test(s), reference standard(s), target condition(s), and study design) and report characteristics (e.g., years considered, language, publication status) used as criteria for eligibility, giving rationale. | 6, 7, Table 1 |
| **Information sources** | 7 | Describe all information sources (e.g., databases with dates of coverage, contact with study authors to identify additional studies) in the search and date last searched. | 7, Appendix 1 |
| **Search** | 8 | Present full search strategies for all electronic databases and other sources searched, including any limits used, such that they could be repeated. | Appendix 1 |
| **Study selection** | 9 | State the process for selecting studies (i.e., screening, eligibility, included in systematic review, and, if applicable, included in the meta-analysis). | 8 |
| **Data collection process** | 10 | Describe method of data extraction from reports (e.g., piloted forms, independently, in duplicate) and any processes for obtaining and confirming data from investigators. | 9, Table 2 |
| **Definitions for data extraction** | 11 | Provide definitions used in data extraction and classifications of target condition(s), index test(s), reference standard(s) and other characteristics (e.g. study design, clinical setting). | 6, 9, Table 1 |
| **Risk of bias and applicability** | 12 | Describe methods used for assessing risk of bias in individual studies and concerns regarding the applicability to the review question. | 9, Appendix 2 |

| Section/topic | # | PRISMA-DTA Checklist Item | Reported on page # |
|---|---|---|---|
| **Diagnostic accuracy measures** | 13 | State the principal diagnostic accuracy measure(s) reported (e.g. sensitivity, specificity) and state the unit of assessment (e.g. per-patient, per-lesion). | 9 |
| **Synthesis of results** | 14 | Describe methods of handling data, combining results of studies and describing variability between studies. This could include, but is not limited to: a) handling of multiple definitions of target condition. b) handling of multiple thresholds of test positivity, c) handling multiple index test readers, d) handling of indeterminate test results, e) grouping and comparing tests, f) handling of different reference standards | 9, 10 |
| **Meta-analysis** | D2 | Report the statistical methods used for meta-analyses, if performed. | 9, 10 |
| **Additional analyses** | 16 | Describe methods of additional analyses (e.g., sensitivity or subgroup analyses, meta-regression), if done, indicating which were pre-specified. | 10 |
| **RESULTS** | | | |
| **Study selection** | 17 | Provide numbers of studies screened, assessed for eligibility, included in the review (and included in meta-analysis, if applicable) with reasons for exclusions at each stage, ideally with a flow diagram. | 10, 11, Figure 1, Appendix 3, Table 3-4 |
| **Study characteristics** | 18 | For each included study provide citations and present key characteristics including: a) participant characteristics (presentation, prior testing), b) clinical setting, c) study design, d) target condition definition, e) index test, f) reference standard, g) sample size, h) funding sources | 11, 12, Table 5-7 |
| **Risk of bias and applicability** | 19 | Present evaluation of risk of bias and concerns regarding applicability for each study. | 12, Table 8, Figure 2 |
| **Results of individual studies** | 20 | For each analysis in each study (e.g. unique combination of index test, reference standard, and positivity threshold) report 2x2 data (TP, FP, FN, TN) with estimates of diagnostic accuracy and confidence intervals, ideally with a forest or receiver operator characteristic (ROC) plot. | 12, Table 9, Figure 3+6 |
| **Synthesis of results** | 21 | Describe test accuracy, including variability; if meta-analysis was done, include results and confidence intervals. | 13, Figure 6, Table 15 |
| **Additional analysis** | 23 | Give results of additional analyses, if done (e.g., sensitivity or subgroup analyses, meta-regression; analysis of index test: failure rates, proportion of inconclusive results, adverse events). | 13, Table 10-15, Figure 4-5+7 |
| **DISCUSSION** | | | |
| **Summary of evidence** | 24 | Summarize the main findings including the strength of evidence. | 13, 14 |
| **Limitations** | 25 | Discuss limitations from included studies (e.g. risk of bias and concerns regarding applicability) and from the review process (e.g. incomplete retrieval of identified research). | 16, 17 |
| **Conclusions** | 26 | Provide a general interpretation of the results in the context of other evidence. Discuss implications for | 17 |

| Section/topic | # | PRISMA-DTA Checklist Item | Reported on page # |
|---|---|---|---|
| | | future research and clinical practice (e.g. the intended use and clinical role of the index test). | |
| **FUNDING** | | | |
| **Funding** | 27 | For the systematic review, describe the sources of funding and other support and the role of the funders. | 2 |

Adapted From: McInnes MDF, Moher D, Thombs BD, McGrath TA, Bossuyt PM, The PRISMA-DTA Group (2018). Preferred Reporting Items for a Systematic Review and Meta-analysis of Diagnostic Test Accuracy Studies: The PRISMA-DTA Statement. JAMA. 2018 Jan 23;319(4):388-396. doi: 10.1001/jama.2017.19163.

### c) PRISMA-S Checklist

| Section/topic | # | Checklist item | Location(s) Reported |
|---|---|---|---|
| **INFORMATION SOURCES AND METHODS** | | | |
| **Database name** | 1 | Name each individual database searched, stating the platform for each. | 7, Appendix 1 |
| **Multi-database searching** | 2 | If databases were searched simultaneously on a single platform, state the name of the platform, listing all of the databases searched. | not applicable |
| **Study registries** | 3 | List any study registries searched. | 7, Appendix 1 |
| **Online resources and browsing** | 4 | Describe any online or print source purposefully searched or browsed (e.g., tables of contents, print conference proceedings, web sites), and how this was done. | 7, 8, Appendix 1 |
| **Citation searching** | 5 | Indicate whether cited references or citing references were examined, and describe any methods used for locating cited/citing references (e.g., browsing reference lists, using a citation index, setting up email alerts for references citing included studies). | 7 |
| **Contacts** | 6 | Indicate whether additional studies or data were sought by contacting authors, experts, manufacturers, or others. | 7 |
| **Other methods** | 7 | Describe any additional information sources or search methods used. | 7 |
| **SEARCH STRATEGIES** | | | |
| **Full search strategies** | 8 | Include the search strategies for each database and information source, copied and pasted exactly as run. | Appendix 1 |
| **Limits and restrictions** | 9 | Specify that no limits were used, or describe any limits or restrictions applied to a search (e.g., date or time period, language, study design) and provide justification for their use. | 6, 7, Appendix 1 |
| **Search filters** | 10 | Indicate whether published search filters were used (as originally designed or modified), and if so, cite the filter(s) used. | Appendix 1 |

| | | | |
|---|---|---|---|
| **Prior work** | 11 | Indicate when search strategies from other literature reviews were adapted or reused for a substantive part or all of the search, citing the previous review(s). | not applicable |
| **Updates** | 12 | Report the methods used to update the search(es) (e.g., rerunning searches, email alerts). | not applicable |
| **Dates of searches** | 13 | For each search strategy, provide the date when the last search occurred. | Appendix 1 |
| **PEER REVIEW** | | | |
| **Peer review** | 14 | Describe any search peer review process. | 7 |
| **MANAGING RECORDS** | | | |
| **Total Records** | 15 | Document the total number of records identified from each database and other information sources. | 10, Figure 1 |
| **Deduplication** | 16 | Describe the processes and any software used to deduplicate records from multiple database searches and other information sources. | 8 |

PRISMA-S: An Extension to the PRISMA Statement for Reporting Literature Searches in Systematic Reviews Rethlefsen ML, Kirtley S, Waffenschmidt S, Ayala AP, Moher D, Page MJ, Koffel JB, PRISMA-S Group. Last updated February 27, 2020.
