## Appendix 1: Search strategies for "Diagnostic accuracy of rapid point-of-care tests for diagnosis of current SARS-CoV-2 infections in children: A systematic review and meta-analysis"

### 1) Bibliographic databases

#### MEDLINE

- Search interface: Ovid
- Segment: Ovid MEDLINE(R) and Epub Ahead of Print, In-Process, In-Data-Review & Other Non-Indexed Citations, Daily and Versions(R) <1946 to May 06, 2021>
- COVID-19 Ovid Filter for MEDLINE: https://ospguides.ovid.com/OSPguides/medline.htm
- Last search: 07/05/2021

| # | Search |
|---|---|
| 1 | Point-of-Care Testing/ |
| 2 | ((molecular or antigen*) adj5 (test* or detect* or diagnos* or assay*)).ti,ab. |
| 3 | (nucleic acid or isothermal or crispr).mp. and (test* or detect* or diagnos* or assay*).ti,ab. |
| 4 | 2 or 3 |
| 5 | (rapid or fast or short* or quick*).ti,ab. or point of care.mp. |
| 6 | (id now or accula or xpert xpress or covid nudge or samba ii or binaxnow or covid-viro or panbio or veritor or nowcheck or biosynex or respi-strip or dart or espline or innova or strongstep or sofia or biocredit or standard q or standard f or bioeasy).mp. |
| 7 | 1 or (4 and 5) or 6 |
| 8 | 7 and (english or german).lg. |
| 9 | limit 8 to yr="2020 -Current" |
| 10 | 9 not (exp Animals/ not exp Humans/) |
| 11 | limit 10 to covid-19 |
| 12 | remove duplicates from 11 |

**Embase**

- Search interface: Ovid
- Segment: Embase <1974 to 2021 May 06>
- COVID-19 Ovid Filter for Embase: https://ospguides.ovid.com/OSPguides/embase.htm
- Last search: 07/05/2021

| # | Search |
|---|---|
| 1 | rapid test/ |
| 2 | ((molecular or antigen*) adj5 (test* or detect* or diagnos* or assay*)).ti,ab. |
| 3 | (nucleic acid or isothermal or crispr).mp. and (test* or detect* or diagnos* or assay*).ti,ab. |
| 4 | 2 or 3 |
| 5 | (rapid or fast or short* or quick*).ti,ab. or point of care.mp. |
| 6 | (id now or accula or xpert xpress or covid nudge or samba ii or binaxnow or covid-viro or panbio or veritor or nowcheck or biosynex or respi-strip or dart or espline or innova or strongstep or sofia or biocredit or standard q or standard f or bioeasy).mp. |
| 7 | 1 or (4 and 5) or 6 |
| 8 | 7 and (english or german).lg. |
| 9 | limit 8 to yr="2020 -Current" |
| 10 | 9 not (exp animal/ not exp human/) |
| 11 | limit 10 to covid-19 |
| 12 | remove duplicates from 11 |
| 13 | 12 not medline.cr. |
| 14 | 13 not (Conference Abstract or Conference Review or Editorial).pt. |

**Cochrane Library**

- Search interface: Wiley
- Database segment: Cochrane Database of Systematic Reviews, Issue 5 of 12, May 2021
- Last Search: 07/05/2021

| ID | Search |
|---|---|
| #1 | [mh ^"Point-of-Care Testing"] |
| #2 | ((molecular or antigen*) NEAR/5 (test* or detect* or diagnos* or assay*)):ti,ab |
| #3 | ("nucleic acid" or isothermal or crispr) and (test* or detect* or diagnos* or assay*):ti,ab |
| #4 | #2 or #3 |
| #5 | (rapid or fast or short* or quick*):ti,ab or "point of care" |
| #6 | ("id now" or accula or "xpert xpress" or "covid nudge" or "samba ii" or binaxnow or covid-viro or panbio or veritor or nowcheck or biosynex or respi-strip or dart or espline or innova or strongstep or sofia or biocredit or "standard q" or "standard f" or bioeasy):ti,ab,kw |
| #7 | #1 or (#4 and #5) or #6 |
| #8 | [mh COVID-19] |
| #9 | ("COVID-19" or "SARS-CoV-2" or "SARS-2" or "SARS2" or "coronavir*" or "corona vir*" or "ncov19" or "ncov-19" or "2019-ncov"):ti,ab |
| #10 | #8 or #9 |

| | |
|---|---|
| #11 | #7 and #10 with Cochrane Library publication date Between Jan 2020 and Jun 2021, in Cochrane Reviews |
| #12 | #11 not ((language next (afr or ara or aze or bos or bul or car or cat or chi or cze or dan or dut or es or est or fin or fre or gre or heb or hrv or hun or ice or ira or ita or jpn or ko or kor or lit or nor or peo or per or pol or por or pt or rom or rum or rus or slo or slv or spa or srp or swe or tha or tur or ukr or urd or uzb)) not (language near/2 (en or eng or english or ger or german or mul or unknown))) |

**International HTA Database**

- Provided by the International Network of Agencies for Health Technology Assessment (INAHTA)
- https://database.inahta.org
- Search interface: Advanced Search
- Filter: Year 2020 to 2021
- Last Search 07/05/2021

| **Search** |
|---|
| (("Point-of-Care Testing"[mh]) OR ((antigen* OR "nucleic acid" OR molecular OR isothermal OR crispr*) AND (diagnos* OR test* OR detect* OR assay*) AND (rapid OR fast OR short* OR quick* OR “point of care”))) AND ((“SARS Virus”[mh]) OR (“Coronavirus Infections”[mh]) OR ("COVID-19" OR "SARS-CoV-2" OR "SARS-2" OR "SARS2" OR "coronavir*" OR "corona vir*" OR "ncov19" OR "ncov-19" OR "2019-ncov")) |

**2) Preprints**

**Europe PMC**

- Search interface: https://europepmc.org
- Filter: Preprints
- Last search: 07/05/2021

| **Search** |
|---|
| ("COVID-19" OR "SARS-CoV-2" OR "SARS-2" OR "SARS2" OR "coronavir*" OR "corona vir*" OR "ncov19" OR "ncov-19" OR "2019-ncov")<br>AND<br>(<br>TITLE:(((antigen* OR molecular OR "nucleic acid" OR crispr OR isothermal) AND (test* OR diagnos* OR detect* OR assay*) AND (rapid OR fast OR quick* OR short* OR “point of care”)) OR (“id now” OR accula OR “xpert xpress” OR “covid nudge” OR “samba ii” OR binaxnow OR covid-viro OR panbio OR veritor OR nowcheck OR biosynex OR respi-strip OR dart OR espline OR innova OR strongstep OR sofia OR biocredit OR “standard q” OR “standard f” OR bioeasy))<br>OR |

| |
|---|
| ABSTRACT:(((antigen* OR molecular OR "nucleic acid" OR crispr OR isothermal) AND (test* OR diagnos* OR detect* OR assay*) AND (rapid OR fast OR quick* OR short* OR “point of care”)) OR (“id now” OR accula OR “xpert xpress” OR “covid nudge” OR “samba ii” OR binaxnow OR covid-viro OR panbio OR veritor OR nowcheck OR biosynex OR respi-strip OR dart OR espline OR innova OR strongstep OR sofia OR biocredit OR “standard q” OR “standard f” OR bioeasy))<br>)<br>AND<br>(HAS_FT:n OR BODY:(child* OR pediatric OR paediatric))<br>AND<br>SRC:PPR |

### 3) Study registries

#### ClinicalTrials.gov

- Provided by the U.S. National Library of Medicine (NLM)
- Search interface: ClinicalTrials.gov Expert Search
- Filter: Eligibility Criteria: Age Group: Child (birth-17)
- Last Search 07/05/2021

| **Search** |
|---|
| (antigen OR antigenic OR molecular OR nucleic acid OR isothermal OR crispr) AND (test OR diagnose OR detect OR assay) AND AREA[ConditionSearch] COVID-19 AND AREA[StdAge] EXPAND[Term] COVER[FullMatch] "Child" |

#### International Clinical Trials Registry Platform (ICTRP)

- Provided by the World Health Organisation (WHO)
- Due to access issues, download of COVID-19 trials repository from https://www.who.int/clinical-trials-registry-platform (accessed online 07/05/2021)

Use R (tidyverse package) to search for entries in column “Public title” or “Scientific title” which include:

> (antigen* OR molecular OR nucleic acid OR isothermal OR crispr) AND (test* OR diagnos* OR detect* OR assay*)

### 4) Further information sources

#### NICE Evidence Search

- Provided by the National Institute for Health and Care Excellence (NICE)
- https://www.evidence.nhs.uk/
- Filter: Evidence Type: Policy and Strategy; Date: From 01/01/2020 to 24/05/2021
- Last search: 24/05/2021

| Search |
|---|
| COVID-19 rapid test |

#### NICE Guidance

- Provided by the National Institute for Health and Care Excellence (NICE)
- Guidance and advice list
- Filter: Area of interest: Covid-19; Status: Published
- Last search: 24/05/2021

#### Foundation for Innovative New Diagnostics (FIND) Website

- https://www.finddx.org/test-directory/
  - Filter: Laboratory/Point-of-care = Point-of-care; FIND evaluation = Yes
- https://www.finddx.org/sarscov2-eval-antigen/
  - Table 1: Antigen(Ag)-detection RDTs undergoing evaluation
- Last search: 24/05/2021
