## Appendix 2: QUADAS-2 Guidance for "Diagnostic accuracy of rapid point-of-care tests for diagnosis of current SARS-CoV-2 infections in children: A systematic review and meta-analysis"

**Appendix 2: Quality Assessment – Review-specific guidance to signaling questions of QUADAS-2 tool.**

### DOMAIN 1: Patient selection

#### A: Risk of Bias

*1) Was a consecutive or random sample of individuals enrolled?*

| | |
|---|---|
| Yes | If it is clear that all eligible individuals were asked to participate in the study within a certain recruitment period or if it is stated that the study enrolled a consecutive or random sample of eligible individuals. |
| No | If a different enrolment method (e.g. via convenience sampling) is described. |
| Unclear | If the enrolment is not described adequately. |

*2) Was a case-control design avoided?*

| | |
|---|---|
| Yes | If the study employed a cross-sectional or cohort study design. |
| No | Not applicable, as “No” would lead to an exclusion of the study. |
| Unclear | If the design of the study is not described clearly enough, so that a definite judgment cannot be made. |

*3) Did the study avoid inappropriate exclusions?*

| | |
|---|---|
| Yes | If most (≥90%) eligible individuals were included without unreasonable selection (see applicability). |
| No | If inappropriate exclusions (>10%) were made that cannot be justified by the test’s instructions for use (IFU) (see applicability) |
| Unclear | If it is not clear whether inappropriate exclusions were made. |

***Could the selection of individuals have introduced bias?***

| | |
|---|---|
| Low | If all questions are answered “Yes”. |
| High | If ≥ 1 question is answered “No”. |
| Unclear | Either if all questions are answered “Unclear” or if ≥ 1 question is answered “Unclear”, and ≥ 1 question is answered “Yes”. |

#### B. Concerns regarding applicability

***Is there concern that the included individuals do not match the review question?***

| | |
|---|---|
| Low | If at least 80% of the included paediatric study participants are below the age of 18 and match the review question and no exclusions are made to adhere to intended use in IFU (e.g. due to symptom onset > 7 days) |

| | |
|---|---|
| High | If paediatric study participants are selected in accordance with the described intended use in the instructions for use (IFU), for example symptom onset ≤ 7 days. The presence of exclusions based on IFU does not increase the risk of bias but may affect applicability. |
| Unclear | If no sufficient information is provided to make a judgment. |

**DOMAIN 2: Index test**

**A: Risk of Bias**

*1) Were the index test results interpreted without knowledge of the results of the reference standard?*

| | |
|---|---|
| Yes | If blinding is explicitly stated or if it is clear that index test was carried out first and the result was read and reported prior to the availability of the result of the reference standard and the process of the read out was described. |
| No | If no blinding was implemented. |
| Unclear | If no sufficient information is provided to make a judgment. |

*2) If a threshold was used, was it pre-specified?*

| | |
|---|---|
| Yes | If a pre-specified threshold was reported in the methods section or if is stated that the test was performed according to the test's IFU. |
| No | If no pre-specified threshold was used. |
| Unclear | If no sufficient information is provided to make a judgment. |

***Could the conduct or interpretation of the index test have introduced bias?***

| | |
|---|---|
| Low | If all questions are answered "Yes". |
| High | If ≥ 1 question is answered "No". |
| Unclear | Either if all questions are answered "Unclear" or if ≥ 1 question is answered "Unclear", and ≥ 1 question is answered "Yes". |

**B. Concerns regarding applicability**

***Is there concern that the index test, its conduct, or interpretation differ from the review question?***

| | |
|---|---|
| Low | If the test kit was used and if the test was performed according to the IFU. |
| High | If the test kit was not used (different swab) or if deviations from the IFU occurred (e.g. usage outside defined temperature range or specimen handling not in accordance with IFU) |
| Unclear | If no sufficient information is provided to make a judgment. |

**DOMAIN 3: Reference Standard**

**A: Risk of Bias**

*1) Is the reference standard likely to correctly classify the target condition?*

| | |
|---|---|
| Yes | If a validated laboratory-based RT-PCR was used. |

| | |
|---|---|
| No | Not applicable, as “No” would lead to an exclusion of the study. |
| Unclear | If no sufficient information is provided to make a judgment. |

*2) Was the general threshold and any additional thresholds that were used pre-specified?*

| | |
|---|---|
| Yes | If all thresholds that were used are reported in the methods section or if is stated that the test was performed according to the laboratory’s protocol or the manufacturer’s IFU. |
| No | If no pre-specified threshold(s) was/were used. |
| Unclear | If no sufficient information is provided to make a judgment. |

*3) Were the reference standard results interpreted without knowledge of the results of the index test?*

| | |
|---|---|
| Yes | If blinding is explicitly stated. |
| No | If no blinding was implemented. |
| Unclear | If no sufficient information is provided to make a judgment. |

***Could the reference standard, its conduct, or its interpretation have introduced bias?***

| | |
|---|---|
| Low | If all questions are answered “Yes”. |
| High | If ≥ 1 question is answered “No”. |
| Unclear | Either if all questions are answered “Unclear” or if ≥ 1 question is answered “Unclear”, and ≥ 1 question is answered “Yes”. |

**B. Concerns regarding applicability**

*Is there concern that the target condition as defined by the reference standard does not match the review question?*

| | |
|---|---|
| Low | If a SARS-CoV-2 RT-PCR assay was used. |
| High | Not applicable. |
| Unclear | If it is not clear, whether a SARS-CoV-2 RT-PCR assay was used. |

**DOMAIN 4: Flow and timing**

**A: Risk of Bias**

*1) Was there an appropriate interval between index test(s) and reference standard?*

| | |
|---|---|
| Yes | If both samples are taken at the same time (concurrently or consecutively) or if it is clear that specimen collection occurred during the same participant encounter in an ambulatory setting. |
| No | If it is clear that both samples are not taken at the same time (time interval > 12 hours) or during one ambulatory visit. |
| Unclear | If no sufficient information is provided to make a judgment. |

*2) Did all study participants receive a reference standard?*

| | |
|---|---|
| Yes | If it is clear that all participants received a reference standard. |
| No | If it is clear that not all participants received a reference standard. |
| Unclear | If no sufficient information is provided to make a judgment. |

*3) Did study participants receive the same reference standard?*

| | |
|---|---|
| Yes | If all participants were tested with a RT-PCR assay. |
| No | Not applicable, as there is only one reference standard defined. |
| Unclear | If no sufficient information is provided to make a judgment. |

4) Were all *study participants* included in the analysis?

| | |
|---|---|
| Yes | If the number of enrolled participants matches the total number of included participants reported in the 2x2 table (for cohort studies with repeat testing over time: only initial test is included in the analysis) |
| No | If number of enrolled participants does not match total number in 2x2 table. “No” does not increase the risk of bias, if difference is ≤ 5 % and reasons for exclusion are reported such as missing data on symptom onset, missing clinical data, missing symptom data, inconclusive/missing index test/reference standard. |
| Unclear | If it is not possible to determine whether all participants were included in the analysis |

***Could the patient flow have introduced bias?***

| | |
|---|---|
| Low | If all questions are answered “Yes”. |
| High | If ≥ 1 question is answered “No”. |
| Unclear | Either if all questions are answered “Unclear” or if ≥ 1 question is answered “Unclear”, and ≥ 1 question is answered “Yes”. |

**FUNDING / CONFLICTS OF INTEREST (COI)**

*Independent evaluation?*

| | |
|---|---|
| Yes | If authors declare no COI and no financial support was provided from industry or other private sources. |
| No | If authors declare relevant COI and/or financial support was provided from industry or other private sources. |
| Unclear | If statement about funding and/or COI is missing. |
