## Appendix 3: List of excluded studies for "Diagnostic accuracy of rapid point-of-care tests for diagnosis of current SARS-CoV-2 infections in children: A systematic review and meta-analysis"

**Appendix 3: List of excluded studies and reasons for exclusion**

**Ineligible Population**

Basso D, Aita A, Padoan A, Cosma C, Navaglia F, Moz S, et al. Salivary SARS-CoV-2 antigen rapid detection: A prospective cohort study. Clin Chim Acta 2021;517:54–9. https://doi.org/10.1016/j.cca.2021.02.014.

Blairon L, Wilmet A, Beukinga I, Tré-Hardy M. Implementation of rapid SARS-CoV-2 antigenic testing in a laboratory without access to molecular methods: Experiences of a general hospital. J Clin Virol 2020;129:104472. https://doi.org/10.1016/j.jcv.2020.104472.

Bokelmann L, Nickel O, Maricic T, Pääbo S, Meyer M, Borte S, et al. Rapid, reliable, and cheap point-of-care bulk testing for SARS-CoV-2 by combining hybridization capture with improved colorimetric LAMP (Cap-iLAMP). MedRxiv 2020:2020.08.04.20168617. https://doi.org/10.1101/2020.08.04.20168617.

Bruzzone B, De Pace V, Caligiuri P, Ricucci V, Guarona G, Pennati BM, et al. Comparative diagnostic performance of rapid antigen detection tests for COVID-19 in a hospital setting. Int J Infect Dis 2021;107:215–8. https://doi.org/10.1016/j.ijid.2021.04.072.

Caruana G, Croxatto A, Kampouri E, Kritikos A, Opota O, Foerster M, et al. Implementing SARS-CoV-2 Rapid Antigen Testing in the Emergency Ward of a Swiss University Hospital: The INCREASE Study. Microorganisms 2021;9. https://doi.org/10.3390/microorganisms9040798.

Cheah PK, Ongkili DF, Zaharuddin FS, Hashim MI, Ho CV, Lee HG, et al. Discrepancy in Screening Performances of Different Rapid Test Kits for SARS-CoV-2; a Letter to Editor. Arch Acad Emerg Med 2021;9:e9.

Ciotti M, Nicolai E, Marcuccilli F, Bernardini S. Use of a rapid point-of-care molecular test in the triage of suspected COVID-19 cases. J Med Virol 2021;93:4088–9. https://doi.org/10.1002/jmv.26944.

Donato LJ, Trivedi VA, Stransky AM, Misra A, Pritt BS, Binnicker MJ, et al. Evaluation of the Cue Health point-of-care COVID-19 (SARS-CoV-2 nucleic acid amplification) test at a community drive through collection center. Diagn Microbiol Infect Dis 2021;100:115307. https://doi.org/10.1016/j.diagmicrobio.2020.115307.

Fenollar F, Bouam A, Ballouche M, Fuster L, Prudent E, Colson P, et al. Evaluation of the Panbio COVID-19 Rapid Antigen Detection Test Device for the Screening of Patients with COVID-19. J Clin Microbiol 2021;59. https://doi.org/10.1128/JCM.02589-20.

FIND Evaluation of Coris BioConcept COVID-19 Ag Respi-Strip, External Report, Version 1.2. 2020.

FIND Evaluation of Shenzhen Bioeasy Biotechnology Co. Ltd. 2019-nCoV Ag Rapid Test Kit (Fluorescence), External Report, Version 1.0. 2021.

FIND Evaluation of Abbott Panbio COVID-19 Ag Rapid Test Device (NASAL) External Report, Version 1.0. 2021.

FIND Evaluation of Fujirebio Inc. Espline SARS-CoV-2, External Report, Version 1.0. 2021.

FIND Evaluation of Bionote, Inc. NowCheck COVID-19 Ag Test, nasal swab, External Report, Version 1.0. 2021.

FIND Evaluation of SD Biosensor, Inc. STANDARD Q COVID-19 Ag Test, nasal swab, External Report, Version 2.0. 2021.

FIND Evaluation of Mologic Ltd, COVID 19 RAPID ANTIGEN TEST, External Report, Version 1.0. 2021.

FIND Evaluation of Edinburgh Genetics ActivXpress+ COVID-19 Antigen Complete Testing Kit, External Report, Version 1.0. 2021.

FIND Evaluation of Premier Medical Corporation Private Limited. Sure Status COVID-19 Card Test, External Report, Version 1.0. 2021.

García-Fiñana M, Hughes DM, Cheyne CP, Burnside G, Stockbridge M, Fowler TA, et al. Performance of the Innova SARS-CoV-2 Antigen Rapid Lateral Flow Test in the Liverpool Asymptomatic Testing Pilot. Rochester, NY: Social Science Research Network; 2021. https://doi.org/10.2139/ssrn.3798558.

Gupta A, Khurana S, Das R, Srigyan D, Singh A, Mittal A, et al. Rapid chromatographic immunoassay-based evaluation of COVID-19: A cross-sectional, diagnostic test accuracy study & its implications for COVID-19 management in India. Indian J Med Res 2021;153:126–31. https://doi.org/10.4103/ijmr.IJMR_3305_20.

Igloi Z, Velzing J, van Beek J, van de Vijver D, Aron G, Ensing R, et al. Clinical Evaluation of Roche SD Biosensor Rapid Antigen Test for SARS-CoV-2 in Municipal Health Service Testing Site, the Netherlands. Emerg Infect Dis 2021;27:1323–9. https://doi.org/10.3201/eid2705.204688.

Klein JAF, Krüger LJ, Tobian F, Gaeddert M, Lainati F, Schnitzler P, et al. Head-to-head performance comparison of self-collected nasal versus professional-collected nasopharyngeal swab for a WHO-listed SARS-CoV-2 antigen-detecting rapid diagnostic test. MedRxiv 2021:2021.03.17.21253076. https://doi.org/10.1101/2021.03.17.21253076.

Kohmer N, Toptan T, Pallas C, Karaca O, Pfeiffer A, Westhaus S, et al. The Comparative Clinical Performance of Four SARS-CoV-2 Rapid Antigen Tests and Their Correlation to Infectivity In Vitro. J Clin Med 2021;10. https://doi.org/10.3390/jcm10020328.

Krüger LJ, Gaeddert M, Tobian F, Lainati F, Gottschalk C, Klein J a. F, et al. Evaluation of the accuracy and ease-of-use of Abbott PanBio - A WHO emergency use listed, rapid, antigen-detecting point-of-care diagnostic test for SARS-CoV-2. MedRxiv 2020:2020.11.27.20239699. https://doi.org/10.1101/2020.11.27.20239699.

Krüger LJ, Gaeddert M, Tobian F, Lainati F, Gottschalk C, Klein JAF, et al. The Abbott PanBio WHO emergency use listed, rapid, antigen-detecting point-of-care diagnostic test for SARS-CoV-2—Evaluation of the accuracy and ease-of-use. PLOS ONE 2021;16:e0247918. https://doi.org/10.1371/journal.pone.0247918.

Li J, Hu X, Wang X, Yang J, Zhang L, Deng Q, et al. A novel One-pot rapid diagnostic technology for COVID-19. Anal Chim Acta 2021;1154:338310. https://doi.org/10.1016/j.aca.2021.338310.

Lindner AK, Nikolai O, Kausch F, Wintel M, Hommes F, Gertler M, et al. Head-to-head comparison of SARS-CoV-2 antigen-detecting rapid test with self-collected nasal swab versus professional-collected nasopharyngeal swab. Eur Respir J 2021;57:2003961. https://doi.org/10.1183/13993003.03961-2020.

Lindner AK, Nikolai O, Rohardt C, Burock S, Hülso C, Bölke A, et al. Head-to-head comparison of SARS-CoV-2 antigen-detecting rapid test with professional-collected nasal versus nasopharyngeal swab. Eur Respir J 2021;57. https://doi.org/10.1183/13993003.04430-2020.

Mahmoud SA, Ibrahim E, Ganesan S, Thakre B, Teddy JG, Raheja P, et al. Evaluation of seven different rapid methods for nucleic acid detection of SARS-COV-2 virus. MedRxiv 2021:2021.04.15.21255533. https://doi.org/10.1101/2021.04.15.21255533.

Moran A, Beavis KG, Matushek SM, Ciaglia C, Francois N, Tesic V, et al. Detection of SARS-CoV-2 by Use of the Cepheid Xpert Xpress SARS-CoV-2 and Roche cobas SARS-CoV-2 Assays. J Clin Microbiol 2020;58. https://doi.org/10.1128/JCM.00772-20.

Nalumansi A, Lutalo T, Kayiwa J, Watera C, Balinandi S, Kiconco J, et al. Field evaluation of the performance of a SARS-CoV-2 antigen rapid diagnostic test in Uganda using nasopharyngeal samples. Int J Infect Dis 2021;104:282–6. https://doi.org/10.1016/j.ijid.2020.10.073.

Oh SM, Jeong H, Chang E, Choe PG, Kang CK, Park WB, et al. Clinical Application of the Standard Q COVID-19 Ag Test for the Detection of SARS-CoV-2 Infection. J Korean Med Sci 2021;36:e101. https://doi.org/10.3346/jkms.2021.36.e101.

Osmanodja B, Budde K, Zickler D, Naik MG, Hofmann J, Gertler M, et al. Diagnostic accuracy of a novel SARS-CoV-2 antigen-detecting rapid diagnostic test from standardized self-collected anterior nasal swabs. MedRxiv 2021:2021.04.20.21255797. https://doi.org/10.1101/2021.04.20.21255797.

Pekosz A, Parvu V, Li M, Andrews JC, Manabe YC, Kodsi S, et al. Antigen-Based Testing but Not Real-Time Polymerase Chain Reaction Correlates With Severe Acute Respiratory Syndrome Coronavirus 2 Viral Culture. Clin Infect Dis 2021. https://doi.org/10.1093/cid/ciaa1706.

Plebani M, Aita A, Cattelan AM, Bonfante F, Padoan A, Giaquinto C, et al. Frequent testing regimen based on salivary samples for an effective COVID-19 containment strategy. MedRxiv 2020:2020.10.13.20210013. https://doi.org/10.1101/2020.10.13.20210013.

Prado JAN del, Reyes AQ, Torre JBL, Loli RG, Olejua AP, Chirinos ERC, et al. Clinical validation of RCSMS: a rapid and sensitive CRISPR-Cas12a test for the molecular detection of SARS-CoV-2 from saliva. MedRxiv 2021:2021.04.26.21256081. https://doi.org/10.1101/2021.04.26.21256081.

Rastawicki W, Gierczyński R, Juszczyk G, Mitura K, Henry BM. Evaluation of PCL rapid point of care antigen test for detection of SARS-CoV-2 in nasopharyngeal swabs. J Med Virol 2021;93:1920–2. https://doi.org/10.1002/jmv.26765.

Schwob JM, Miauton A, Petrovic D, Perdrix J, Senn N, Jaton K, et al. Antigen rapid tests, nasopharyngeal PCR and saliva PCR to detect SARS-CoV-2: a prospective comparative clinical trial. MedRxiv 2020:2020.11.23.20237057. https://doi.org/10.1101/2020.11.23.20237057.

Serei VD, Cristelli R, Joho K, Salaru G, Kirn T, Carayannopoulous MO, et al. Comparison of abbott ID NOW COVID-19 rapid molecular assay to cepheid xpert xpress SARS-CoV-2 assay in dry nasal swabs. Diagn Microbiol Infect Dis 2021;99:115208. https://doi.org/10.1016/j.diagmicrobio.2020.115208.

Smith RL, Gibson LL, Martinez PP, Ke R, Mirza A, Conte M, et al. Longitudinal assessment of diagnostic test performance over the course of acute SARS-CoV-2 infection. MedRxiv 2021:2021.03.19.21253964. https://doi.org/10.1101/2021.03.19.21253964.

SoRelle JA, Mahimainathan L, McCormick-Baw C, Cavuoti D, Lee F, Bararia A, et al. Evaluation of symptomatic patient saliva as a sample type for the Abbott ID NOW COVID-19 assay. MedRxiv 2020:2020.06.01.20119198. https://doi.org/10.1101/2020.06.01.20119198.

Stohr JJJM, Zwart VF, Goderski G, Meijer A, Nagel-Imming CRS, Bergh MFQK den, et al. Self-testing for the detection of SARS-CoV-2 infection with rapid antigen tests. MedRxiv 2021:2021.02.21.21252153. https://doi.org/10.1101/2021.02.21.21252153.

Sundah NR, Natalia A, Liu Y, Ho NRY, Zhao H, Chen Y, et al. Catalytic amplification by transition-state molecular switches for direct and sensitive detection of SARS-CoV-2. Sci Adv 2021;7. https://doi.org/10.1126/sciadv.abe5940.

Thakur P, Saxena S, Manchanda V, Rana N, Goel R, Arora R. Utility of Antigen-Based Rapid Diagnostic Test for Detection of SARS-CoV-2 Virus in Routine Hospital Settings. Lab Med 2021. https://doi.org/10.1093/labmed/lmab033.

Tu Y-P, Iqbal J, O'Leary T. Sensitivity of ID NOW and RT-PCR for detection of SARS-CoV-2 in an ambulatory population. Elife 2021;10. https://doi.org/10.7554/eLife.65726.

Tworek JA, Khan F, Sekedat MD, Scheidel C, Malani AN. The Utility of Rapid Nucleic Acid Amplification Testing to Triage Symptomatic Patients and to Screen Asymptomatic Preprocedure Patients for SARS-CoV-2. Open Forum Infect Dis 2021;8:ofaa607. https://doi.org/10.1093/ofid/ofaa607.

Uprety P, Mathew J, Weinstein MP, Kirn TJ. Comparison of 4 molecular assays for detection of severe acute respiratory syndrome coronavirus 2 (SARS-CoV-2). Diagn Microbiol Infect Dis 2021;99:115196. https://doi.org/10.1016/j.diagmicrobio.2020.115196.

Van J-CN, Gerlier C, Pilmis B, Mizrahi A, Ponfilly GP de, Khaterchi A, et al. Prospective evaluation of ID NOW COVID-19 assay used as point-of-care test in an Emergency Department. MedRxiv 2021:2021.03.29.21253909. https://doi.org/10.1101/2021.03.29.21253909.

Wang X, Zhong M, Liu Y, Ma P, Dang L, Meng Q, et al. Rapid and sensitive detection of COVID-19 using CRISPR/Cas12a-based detection with naked eye readout, CRISPR/Cas12a-NER. Sci Bull (Beijing) 2020;65:1436–9. https://doi.org/10.1016/j.scib.2020.04.041.

Yin N, Debuysschere C, Decroly M, Bouazza F-Z, Collot V, Martin C, et al. SARS-CoV-2 Diagnostic Tests: Algorithm and Field Evaluation From the Near Patient Testing to the Automated Diagnostic Platform. Front Med (Lausanne) 2021;8:650581. https://doi.org/10.3389/fmed.2021.650581.

Yong Chua PE, Gwee SXW, Wang MX, Hao G, Pang J. SARS-CoV-2 Diagnostic Tests for Reopening of Borders: A Systematic Review and Meta-Analysis. Rochester, NY: Social Science Research Network; 2021. https://doi.org/10.2139/ssrn.3774144.

Yoshimi K, Takeshita K, Yamayoshi S, Shibumura S, Yamauchi Y, Yamamoto M, et al. Rapid and Accurate Detection of Novel Coronavirus SARS-CoV-2 Using CRISPR-Cas3. Rochester, NY: Social Science Research Network; 2020. https://doi.org/10.2139/ssrn.3640844.

Yu L, Wu S, Hao X, Dong X, Mao L, Pelechano V, et al. Rapid Detection of COVID-19 Coronavirus Using a Reverse Transcriptional Loop-Mediated Isothermal Amplification (RT-LAMP) Diagnostic Platform. Clin Chem 2020;66:975–7. https://doi.org/10.1093/clinchem/hvaa102.

**Ineligible index test**

Alemany A, Baró B, Ouchi D, Rodó P, Ubals M, Corbacho-Monné M, et al. Analytical and clinical performance of the panbio COVID-19 antigen-detecting rapid diagnostic test. J Infect 2021;82:186–230. https://doi.org/10.1016/j.jinf.2020.12.033.

Aoki K, Nagasawa T, Ishii Y, Yagi S, Kashiwagi K, Miyazaki T, et al. Evaluation of clinical utility of novel coronavirus antigen detection reagent, Espline® SARS-CoV-2. J Infect Chemother 2021;27:319–22. https://doi.org/10.1016/j.jiac.2020.11.015.

Barauna VG, Singh MN, Barbosa LL, Marcarini WD, Vassallo PF, Mill JG, et al. Ultrarapid On-Site Detection of SARS-CoV-2 Infection Using Simple ATR-FTIR Spectroscopy and an Analysis Algorithm: High Sensitivity and Specificity. Anal Chem 2021;93:2950–8. https://doi.org/10.1021/acs.analchem.0c04608.

Baro B, Rodo P, Ouchi D, Bordoy AE, Amaro ENS, Salsench SV, et al. Performance characteristics of five antigen-detecting rapid diagnostic test (Ag-RDT) for SARS-CoV-2 asymptomatic infection: a head-to-head benchmark comparison. MedRxiv 2021:2021.02.11.21251553. https://doi.org/10.1101/2021.02.11.21251553.

Ben-Assa N, Naddaf R, Gefen T, Capucha T, Hajjo H, Mandelbaum N, et al. SARS-CoV-2 On-the-Spot Virus Detection Directly from Patients. MedRxiv 2020:2020.04.22.20072389. https://doi.org/10.1101/2020.04.22.20072389.

Brandsma E, Verhagen HJMP, van de Laar TJW, Claas ECJ, Cornelissen M, van den Akker E. Rapid, Sensitive, and Specific Severe Acute Respiratory Syndrome Coronavirus 2 Detection: A Multicenter Comparison Between Standard Quantitative Reverse-Transcriptase Polymerase Chain Reaction and CRISPR-Based DETECTR. J Infect Dis 2021;223:206–13. https://doi.org/10.1093/infdis/jiaa641.

Brotons P, Perez-Argüello A, Launes C, Torrents F, Saucedo J, Claverol J, et al. Validation and implementation of a direct RT-qPCR method for rapid screening of SARS-CoV-2 infection by using non-invasive saliva samples. MedRxiv 2020:2020.11.19.20234245. https://doi.org/10.1101/2020.11.19.20234245.

Cameron A, Pecora ND, Pettengill MA. Extraction-Free Methods for the Detection of SARS-CoV-2 by Reverse Transcription-PCR: a Comparison with the Cepheid Xpert Xpress SARS-CoV-2 Assay across Two Medical Centers. J Clin Microbiol 2021;59. https://doi.org/10.1128/JCM.02643-20.

Cerutti F, Burdino E, Milia MG, Allice T, Gregori G, Bruzzone B, et al. Urgent need of rapid tests for SARS CoV-2 antigen detection: Evaluation of the SD-Biosensor antigen test for SARS-CoV-2. J Clin Virol 2020;132:104654. https://doi.org/10.1016/j.jcv.2020.104654.

Chaimayo C, Kaewnaphan B, Tanlieng N, Athipanyasilp N, Sirijatuphat R, Chayakulkeeree M, et al. Rapid SARS-CoV-2 antigen detection assay in comparison with real-time RT-PCR assay for laboratory diagnosis of COVID-19 in Thailand. Virol J 2020;17:177. https://doi.org/10.1186/s12985-020-01452-5.

Diao B, Wen K, Zhang J, Chen J, Han C, Chen Y, et al. Accuracy of a nucleocapsid protein antigen rapid test in the diagnosis of SARS-CoV-2 infection. Clin Microbiol Infect 2021;27:289.e1-289.e4. https://doi.org/10.1016/j.cmi.2020.09.057.

Egerer R, Edel B, Löffler B, Henke A, Rödel J. Performance of the RT-LAMP-based eazyplex® SARS-CoV-2 as a novel rapid diagnostic test. J Clin Virol 2021;138:104817. https://doi.org/10.1016/j.jcv.2021.104817.

Fitoussi F, Dupont R, Tonen-Wolyec S, Bélec L. Performances of the VitaPCR™ SARS-CoV-2 Assay during the second wave of the COVID-19 epidemic in France. J Med Virol 2021;93:4351–7. https://doi.org/10.1002/jmv.26950.

Ghofrani M, Casas MT, Pelz RK, Kroll C, Blum N, Foster SD. Performance characteristics of the ID NOW COVID-19 assay: A regional health care system experience. MedRxiv 2020:2020.06.03.20116327. https://doi.org/10.1101/2020.06.03.20116327.

Gouilh MA, Cassier R, Maille E, Schanen C, Rocque L-M, Vabret A. An easy, reliable and rapid SARS-CoV2 RT-LAMP based test for Point-of-Care and diagnostic lab. MedRxiv 2020:2020.09.25.20200956. https://doi.org/10.1101/2020.09.25.20200956.

Gremmels H, Winkel BMF, Schuurman R, Rosingh A, Rigter NAM, Rodriguez O, et al. Real-life validation of the Panbio™ COVID-19 antigen rapid test (Abbott) in community-dwelling subjects with symptoms of potential SARS-CoV-2 infection. EClinicalMedicine 2021;31:100677. https://doi.org/10.1016/j.eclinm.2020.100677.

Gupta N, Rana S, Singh H. Innovative point-of-care molecular diagnostic test for COVID-19 in India. Lancet Microbe 2020;1:e277. https://doi.org/10.1016/S2666-5247(20)30164-6.

Hansen G, Marino J, Wang Z-X, Beavis KG, Rodrigo J, Labog K, et al. Clinical Performance of the Point-of-Care cobas Liat for Detection of SARS-CoV-2 in 20 Minutes: a Multicenter Study. J Clin Microbiol 2021;59. https://doi.org/10.1128/JCM.02811-20.

Haq F, Sharif S, Khurshid A, Ikram A, Shabbir I, Salman M, et al. Reverse transcriptase loop-mediated isothermal amplification (RT-LAMP)-based diagnosis: A potential alternative to quantitative real-time PCR based detection of the novel SARS-COV-2 virus. Saudi J Biol Sci 2021;28:942–7. https://doi.org/10.1016/j.sjbs.2020.10.064.

Harrington A, Cox B, Snowdon J, Bakst J, Ley E, Grajales P, et al. Comparison of Abbott ID Now and Abbott m2000 Methods for the Detection of SARS-CoV-2 from Nasopharyngeal and Nasal Swabs from Symptomatic Patients. Journal of Clinical Microbiology n.d.;58:e00798-20. https://doi.org/10.1128/JCM.00798-20.

Ishii T, Sasaki M, Yamada K, Kato D, Osuka H, Aoki K, et al. Immunochromatography and chemiluminescent enzyme immunoassay for COVID-19 diagnosis. J Infect Chemother 2021;27:915–8. https://doi.org/10.1016/j.jiac.2021.02.025.

Kim D, Lee J, Bal J, Seo SK, Chong C-K, Lee JH, et al. Development and Clinical Evaluation of an Immunochromatography-Based Rapid Antigen Test (GenBody™ COVAG025) for COVID-19 Diagnosis. Viruses 2021;13. https://doi.org/10.3390/v13050796.

Kitagawa Y, Orihara Y, Kawamura R, Imai K, Sakai J, Tarumoto N, et al. Evaluation of rapid diagnosis of novel coronavirus disease (COVID-19) using loop-mediated isothermal amplification. J Clin Virol 2020;129:104446. https://doi.org/10.1016/j.jcv.2020.104446.

Krause E, Puyskens A, Bourquain D, Brinkmann A, Biere B, Schaade L, et al. Sensitive on-site detection of SARS-CoV-2 by ID NOW COVID-19. MedRxiv 2021:2021.04.18.21255688. https://doi.org/10.1101/2021.04.18.21255688.

Lambert-Niclot S, Cuffel A, Le Pape S, Vauloup-Fellous C, Morand-Joubert L, Roque-Afonso A-M, et al. Evaluation of a Rapid Diagnostic Assay for Detection of SARS-CoV-2 Antigen in Nasopharyngeal Swabs. J Clin Microbiol 2020;58. https://doi.org/10.1128/JCM.00977-20.

Lephart PR, Bachman MA, LeBar W, McClellan S, Barron K, Schroeder L, et al. Comparative study of four SARS-CoV-2 Nucleic Acid Amplification Test (NAAT) platforms demonstrates that ID NOW performance is impaired substantially by patient and specimen type. Diagn Microbiol Infect Dis 2021;99:115200. https://doi.org/10.1016/j.diagmicrobio.2020.115200.

Lu R, Wu X, Wan Z, Li Y, Zuo L, Qin J, et al. Development of a Novel Reverse Transcription Loop-Mediated Isothermal Amplification Method for Rapid Detection of SARS-CoV-2. Virol Sin 2020;35:344–7. https://doi.org/10.1007/s12250-020-00218-1.

Mboumba Bouassa R-S, Veyer D, Péré H, Bélec L. Analytical performances of the point-of-care SIENNA™ COVID-19 Antigen Rapid Test for the detection of SARS-CoV-2 nucleocapsid protein in nasopharyngeal swabs: A prospective evaluation during the COVID-19 second wave in France. Int J Infect Dis 2021;106:8–12. https://doi.org/10.1016/j.ijid.2021.03.051.

McCormick-Baw C, Morgan K, Gaffney D, Cazares Y, Jaworski K, Byrd A, et al. Saliva as an Alternate Specimen Source for Detection of SARS-CoV-2 in Symptomatic Patients Using Cepheid Xpert Xpress SARS-CoV-2. J Clin Microbiol 2020;58. https://doi.org/10.1128/JCM.01109-20.

Mohon AN, Oberding L, Hundt J, van Marle G, Pabbaraju K, Berenger BM, et al. Optimization and clinical validation of dual-target RT-LAMP for SARS-CoV-2. J Virol Methods 2020;286:113972. https://doi.org/10.1016/j.jviromet.2020.113972.

Nawattanapaiboon K, Pasomsub E, Prombun P, Wongbunmak A, Jenjitwanich A, Mahasupachai P, et al. Colorimetric reverse transcription loop-mediated isothermal amplification (RT-LAMP) as a visual diagnostic platform for the detection of the emerging coronavirus SARS-CoV-2. Analyst 2021;146:471–7. https://doi.org/10.1039/d0an01775b.

Porte L, Legarraga P, Iruretagoyena M, Vollrath V, Pizarro G, Munita JM, et al. Rapid SARS-CoV-2 antigen detection by immunofluorescence – a new tool to detect infectivity. MedRxiv 2020:2020.10.04.20206466. https://doi.org/10.1101/2020.10.04.20206466.

Public Health England. COVID-19: paediatric surveillance. GOVUK n.d. https://www.gov.uk/guidance/covid-19-paediatric-surveillance (accessed June 20, 2021).

Rearigh LM, Hewlett AL, Fey PD, Broadhurst MJ, Brett-Major DM, Rupp ME, et al. Utility of repeat testing for COVID-19: Laboratory stewardship when the stakes are high. Infect Control Hosp Epidemiol 2021;42:338–40. https://doi.org/10.1017/ice.2020.397.

Renzoni A, Perez F, Ngo Nsoga MT, Yerly S, Boehm E, Gayet-Ageron A, et al. Analytical Evaluation of Visby Medical RT-PCR Portable Device for Rapid Detection of SARS-CoV-2. Diagnostics (Basel) 2021;11. https://doi.org/10.3390/diagnostics11050813.

Sakai J, Tarumoto N, Orihara Y, Kawamura R, Kodana M, Matsuzaki N, et al. Evaluation of a high-speed but low-throughput RT-qPCR system for detection of SARS-CoV-2. J Hosp Infect 2020;105:615–8. https://doi.org/10.1016/j.jhin.2020.05.025.

Scohy A, Anantharajah A, Bodéus M, Kabamba-Mukadi B, Verroken A, Rodriguez-Villalobos H. Low performance of rapid antigen detection test as frontline testing for COVID-19 diagnosis. J Clin Virol 2020;129:104455. https://doi.org/10.1016/j.jcv.2020.104455.

Tham JWM, Ng SC, Chai CN, Png S, Tan EJM, Ng LJ, et al. Parallel testing of 241 clinical nasopharyngeal swabs for the detection of SARS-CoV-2 virus on the Cepheid Xpert Xpress SARS-CoV-2 and the Roche cobas SARS-CoV-2 assays. Clin Chem Lab Med 2020;59:e45–8. https://doi.org/10.1515/cclm-2020-1338.

Toptan T, Eckermann L, Pfeiffer AE, Hoehl S, Ciesek S, Drosten C, et al. Evaluation of a SARS-CoV-2 rapid antigen test: Potential to help reduce community spread? J Clin Virol 2021;135:104713. https://doi.org/10.1016/j.jcv.2020.104713.

Trobajo-Sanmartín C, Navascués A, Miqueleiz A, Ezpeleta C. Evaluation of the rapid antigen test CerTest SARS-CoV-2 as an alternative COVID-19 diagnosis technique. Infect Dis (Lond) 2021:1–3. https://doi.org/10.1080/23744235.2021.1902563.

Vaz SN, Santana DS de, Netto EM, Wang W-K, Brites C. Validation of the GeneXpert Xpress SARS-CoV-2 PCR assay using saliva as biological specimen. The Brazilian Journal of Infectious Diseases 2021;25:101543. https://doi.org/10.1016/j.bjid.2021.101543.

Wang F, Yang J, He R, Yu X, Chen S, Liu Y, et al. PfAgo-based detection of SARS-CoV-2. Biosensors and Bioelectronics 2021;177:112932. https://doi.org/10.1016/j.bios.2020.112932.

Watanabe R, Asai S, Kakizoe H, Saeki H, Masukawa A, Miyazawa M, et al. Evaluation of the basic assay performance of the GeneSoc® rapid PCR testing system for detection of severe acute respiratory syndrome coronavirus 2. PLoS One 2021;16:e0248397. https://doi.org/10.1371/journal.pone.0248397.

Wei S, Kohl E, Djandji A, Morgan S, Whittier S, Mansukhani M, et al. Direct diagnostic testing of SARS-CoV-2 without the need for prior RNA extraction. Sci Rep 2021;11:2402. https://doi.org/10.1038/s41598-021-81487-y.

Wei S, Suryawanshi H, Djandji A, Kohl E, Morgan S, Hod EA, et al. Field-deployable, rapid diagnostic testing of saliva for SARS-CoV-2. Scientific Reports 2021;11:5448. https://doi.org/10.1038/s41598-021-84792-8.

Wen D, Yang S, Li G, Xuan Q, Guo W, Wu W. Sample-to-Answer and Routine Real-Time rRT-PCR: A Comparison of Different Platforms for SARS-CoV-2 Detection. J Mol Diagn 2021. https://doi.org/10.1016/j.jmoldx.2021.02.010.

Yamazaki W, Matsumura Y, Thongchankaew-Seo U, Yamazaki Y, Nagao M. Development of a point-of-care test to detect SARS-CoV-2 from saliva which combines a simple RNA extraction method with colorimetric reverse transcription loop-mediated isothermal amplification detection. J Clin Virol 2021;136:104760. https://doi.org/10.1016/j.jcv.2021.104760.

Yan C, Cui J, Huang L, Du B, Chen L, Xue G, et al. Rapid and visual detection of 2019 novel coronavirus (SARS-CoV-2) by a reverse transcription loop-mediated isothermal amplification assay. Clin Microbiol Infect 2020;26:773–9. https://doi.org/10.1016/j.cmi.2020.04.001.

Yoshikawa R, Abe H, Igasaki Y, Negishi S, Goto H, Yasuda J. Development and evaluation of a rapid and simple diagnostic assay for COVID-19 based on loop-mediated isothermal amplification. PLoS Negl Trop Dis 2020;14:e0008855. https://doi.org/10.1371/journal.pntd.0008855.

Zheng Y-Z, Chen J-T, Li J, Wu X-J, Wen J-Z, Liu X-Z, et al. Reverse Transcription Recombinase-Aided Amplification Assay With Lateral Flow Dipstick Assay for Rapid Detection of 2019 Novel Coronavirus. Front Cell Infect Microbiol 2021;11:613304. https://doi.org/10.3389/fcimb.2021.613304.

Zwirglmaier K, Weyh M, Krüger C, Ehmann R, Müller K, Wölfel R, et al. Rapid detection of SARS-CoV-2 by pulse-controlled amplification (PCA). Journal of Virological Methods 2021;290:114083. https://doi.org/10.1016/j.jviromet.2021.114083.

**Ineligible reference standard**

Beck ET, Paar W, Fojut L, Serwe J, Jahnke RR. Comparison of the Quidel Sofia SARS FIA Test to the Hologic Aptima SARS-CoV-2 TMA Test for Diagnosis of COVID-19 in Symptomatic Outpatients. J Clin Microbiol 2021;59. https://doi.org/10.1128/JCM.02727-20.

**Ineligible study design**

Anahtar MN, McGrath GEG, Rabe BA, Tanner NA, White BA, Lennerz JKM, et al. Clinical Assessment and Validation of a Rapid and Sensitive SARS-CoV-2 Test Using Reverse Transcription Loop-Mediated Isothermal Amplification Without the Need for RNA Extraction. Open Forum Infectious Diseases 2021;8. https://doi.org/10.1093/ofid/ofaa631.

Annamalai P, Kanta M, Ramu P, Ravi B, Veerapandian K, Srinivasan R. A Simple Colorimetric Molecular Detection of Novel Coronavirus (COVID-19), an Essential Diagnostic Tool for Pandemic Screening. MedRxiv 2020:2020.04.10.20060293. https://doi.org/10.1101/2020.04.10.20060293.

Arizti-Sanz J, Freije CA, Stanton AC, Boehm CK, Petros BA, Siddiqui S, et al. Integrated sample inactivation, amplification, and Cas13-based detection of SARS-CoV-2. BioRxiv 2020:2020.05.28.119131. https://doi.org/10.1101/2020.05.28.119131.

Basawarajappa SG, Rangaiah A, Padukone S, Yadav PD, Gupta N, Shankar SM. Performance evaluation of Truenat$^{TM}$ Beta CoV & Truenat$^{TM}$ SARS-CoV-2 point-of-care assays for coronavirus disease 2019. Indian J Med Res 2021;153:144–50. https://doi.org/10.4103/ijmr.IJMR_2363_20.

Behrmann O, Bachmann I, Hufert F, Dame G. Schnellnachweis von SARS-CoV-2 mit recombinase polymerase amplification. Biospektrum (Heidelb) 2020;26:624–7. https://doi.org/10.1007/s12268-020-1458-3.

Carpenter CR. Rapid antigen and molecular tests had varied sensitivity and ≥97% specificity for detecting SARS-CoV-2 infection. Ann Intern Med 2020;173:JC69. https://doi.org/10.7326/ACPJ202012150-069.

Castro R, Luz PM, Wakimoto MD, Veloso VG, Grinsztejn B, Perazzo H. COVID-19: a meta-analysis of diagnostic test accuracy of commercial assays registered in Brazil. Braz J Infect Dis 2020;24:180–7. https://doi.org/10.1016/j.bjid.2020.04.003.

Chauhan N, Soni S, Gupta A, Jain U. New and developing diagnostic platforms for COVID-19: A systematic review. Expert Rev Mol Diagn 2020;20:971–83. https://doi.org/10.1080/14737159.2020.1816466.

Cole KH, Bouin A, Ruiz C, Semler BL, Inlay MA, Lupták A. Single-tube collection and nucleic acid analysis of clinical samples: a rapid approach for SARS-CoV-2 saliva testing. MedRxiv 2021:2021.04.29.21256345. https://doi.org/10.1101/2021.04.29.21256345.

Ebrahimi M, Harmooshi NN, Rahim F. Diagnostic Utility of Antigen Detection Rapid Diagnostic Tests for Covid- 19: A Systematic Review and Meta-Analysis. MedRxiv 2021:2021.04.02.21254714. https://doi.org/10.1101/2021.04.02.21254714.

Farfour E, Asso-Bonnet M, Vasse M, SARS-CoV-2 Foch Hospital study group. The ID NOW COVID-19, a high-speed high-performance assay. Eur J Clin Microbiol Infect Dis 2021. https://doi.org/10.1007/s10096-021-04243-0.

Hou H, Chen J, Wang Y, Lu Y, Zhu Y, Zhang B, et al. Multicenter Evaluation of the Cepheid Xpert Xpress SARS-CoV-2 Assay for the Detection of SARS-CoV-2 in Oropharyngeal Swab Specimens. J Clin Microbiol 2020;58. https://doi.org/10.1128/JCM.01288-20.

Huang L, Ding L, Zhou J, Chen S, Chen F, Zhao C, et al. One-step rapid quantification of SARS-CoV-2 virus particles via low-cost nanoplasmonic sensors in generic microplate reader and point-of-care device. Biosensors and Bioelectronics 2021;171:112685. https://doi.org/10.1016/j.bios.2020.112685.

Jamshaid H, Zahid F, Din I ud, Zeb A, Choi HG, Khan GM, et al. Diagnostic and Treatment Strategies for COVID-19. AAPS PharmSciTech 2020;21:222. https://doi.org/10.1208/s12249-020-01756-3.

Joung J, Ladha A, Saito M, Kim N-G, Woolley AE, Segel M, et al. Detection of SARS-CoV-2 with SHERLOCK One-Pot Testing. N Engl J Med 2020;383:1492–4. https://doi.org/10.1056/NEJMc2026172.

Kari Broder, Ahmed Babiker, Charles Myers, Terri White, Heather Jones, John Cardella, et al. Test Agreement between Roche Cobas 6800 and Cepheid GeneXpert Xpress SARS-CoV-2 Assays at High Cycle Threshold Ranges. Journal of Clinical Microbiology 2020;58:e01187-20. https://doi.org/doi:10.1128/JCM.01187-20.

Kellner MJ, Ross JJ, Schnabl J, Dekens MPS, Heinen R, Grishkovskaya I, et al. A rapid, highly sensitive and open-access SARS-CoV-2 detection assay for laboratory and home testing. BioRxiv 2020:2020.06.23.166397. https://doi.org/10.1101/2020.06.23.166397.

Lau YL, Ismail I, Mustapa NI, Lai MY, Tuan Soh TS, Hassan A, et al. Real-time reverse transcription loop-mediated isothermal amplification for rapid detection of SARS-CoV-2. PeerJ 2020;8:e9278. https://doi.org/10.7717/peerj.9278.

Lee L, Liu F, Chen Y, Roma G. Quantitative and Ultrasensitive In-situ Immunoassay Technology for SARS-CoV-2 Detection in Saliva. Res Sq 2021. https://doi.org/10.21203/rs.3.rs-138025/v1.

Liotti FM, Menchinelli G, Lalle E, Palucci I, Marchetti S, Colavita F, et al. Performance of a novel diagnostic assay for rapid SARS-CoV-2 antigen detection in nasopharynx samples. Clin Microbiol Infect 2021;27:487–8. https://doi.org/10.1016/j.cmi.2020.09.030.

Lu R, Wu X, Wan Z, Li Y, Jin X, Zhang C. A Novel Reverse Transcription Loop-Mediated Isothermal Amplification Method for Rapid Detection of SARS-CoV-2. Int J Mol Sci 2020;21. https://doi.org/10.3390/ijms21082826.

Mahendiratta S, Batra G, Sarma P, Kumar H, Bansal S, Kumar S, et al. Molecular diagnosis of COVID-19 in different biologic matrix, their diagnostic validity and clinical relevance: A systematic review. Life Sci 2020;258:118207. https://doi.org/10.1016/j.lfs.2020.118207.

Olearo F, Nörz D, Heinrich F, Sutter JP, Roedl K, Schultze A, et al. Handling and accuracy of four rapid antigen tests for the diagnosis of SARS-CoV-2 compared to RT-qPCR. J Clin Virol 2021;137:104782. https://doi.org/10.1016/j.jcv.2021.104782.

Public Health England. SARS-CoV-2 lateral flow antigen tests: evaluation of VOC1 and VOC2. GOVUK n.d. https://www.gov.uk/government/publications/sars-cov-2-lateral-flow-antigen-tests-evaluation-of-voc1-and-voc2 (accessed June 20, 2021).

Public Health England. SARS-CoV-2 lateral flow antigen tests: evaluation of VUI-202012/01. GOVUK n.d. https://www.gov.uk/government/publications/sars-cov-2-lateral-flow-antigen-tests-evaluation-of-vui-20201201 (accessed June 20, 2021).

Ridgway JP, Pisano J, Landon E, Beavis KG, Robicsek A. Clinical Sensitivity of Severe Acute Respiratory Syndrome Coronavirus 2 Nucleic Acid Amplification Tests for Diagnosing Coronavirus Disease 2019. Open Forum Infectious Diseases 2020;7. https://doi.org/10.1093/ofid/ofaa315.

Shirato K, Nao N, Matsuyama S, Takeda M, Kageyama T. An Ultra-Rapid Real-Time RT-PCR Method Using the PCR1100 to Detect Severe Acute Respiratory Syndrome Coronavirus-2. Jpn J Infect Dis 2021;74:29–34. https://doi.org/10.7883/yoken.JJID.2020.324.

Shirvani A, Azimi L, Ghanaie RM, Alebouyeh M, Fallah F, Tabatabaei SR, et al. Utility of available methods for diagnosing SARS-CoV-2 in clinical samples. Archives of Pediatric Infectious Diseases 2020;8. https://doi.org/10.5812/pedinfect.103677.

Varlamov DA, Blagodatskikh KA, Smirnova EV, Kramarov VM, Ignatov KB. Combinations of PCR and Isothermal Amplification Techniques Are Suitable for Fast and Sensitive Detection of SARS-CoV-2 Viral RNA. Front Bioeng Biotechnol 2020;8:604793. https://doi.org/10.3389/fbioe.2020.604793.

Woo CH, Jang S, Shin G, Jung GY, Lee JW. Sensitive fluorescence detection of SARS-CoV-2 RNA in clinical samples via one-pot isothermal ligation and transcription. Nat Biomed Eng 2020;4:1168–79. https://doi.org/10.1038/s41551-020-00617-5.

Xu Y, Rather A, Song S, Fang J-C, Dupont RL, Kara UI, et al. Ultrasensitive and Selective Detection of SARS-CoV-2 Using Thermotropic Liquid Crystals and Image-Based Machine Learning. SSRN Journal 2020. https://doi.org/10.2139/ssrn.3682267.

**Inadequate unit of analysis**

Bonde J, Ejegod D, Pedersen H, Smith B, Cortes D, Leding C, et al. Clinical validation of point-of-care SARS-COV-2 BD Veritor antigen test by a single throat swab for rapid COVID-19 status on hospital patients predominantly without overt COVID symptoms. MedRxiv 2021:2021.04.12.21255299. https://doi.org/10.1101/2021.04.12.21255299.

**Inadequate sample size**

Abdulrahman A, Mustafa F, AlAwadhi AI, Alansari Q, AlAlawi B, AlQahtani M. Comparison of SARS-COV-2 nasal antigen test to nasopharyngeal RT-PCR in mildly symptomatic patients. MedRxiv 2020:2020.11.10.20228973. https://doi.org/10.1101/2020.11.10.20228973.

Agulló V, Fernández-González M, Ortiz de la Tabla V, Gonzalo-Jiménez N, García JA, Masiá M, et al. Evaluation of the rapid antigen test Panbio COVID-19 in saliva and nasal swabs in a population-based point-of-care study. J Infect 2021;82:186–230. https://doi.org/10.1016/j.jinf.2020.12.007.

Albert E, Torres I, Bueno F, Huntley D, Molla E, Fernández-Fuentes MÁ, et al. Field evaluation of a rapid antigen test (Panbio™ COVID-19 Ag Rapid Test Device) for COVID-19 diagnosis in primary healthcare centres. Clin Microbiol Infect 2021;27:472.e7-472.e10. https://doi.org/10.1016/j.cmi.2020.11.004.

Berger A, Nsoga MTN, Perez-Rodriguez FJ, Aad YA, Sattonnet-Roche P, Gayet-Ageron A, et al. Diagnostic accuracy of two commercial SARS-CoV-2 antigen-detecting rapid tests at the point of care in community-based testing centers. PLoS One 2021;16:e0248921. https://doi.org/10.1371/journal.pone.0248921.

Ciotti M, Maurici M, Pieri M, Andreoni M, Bernardini S. Performance of a rapid antigen test in the diagnosis of SARS-CoV-2 infection. J Med Virol 2021;93:2988–91. https://doi.org/10.1002/jmv.26830.

Favresse J, Gillot C, Oliveira M, Cadrobbi J, Elsen M, Eucher C, et al. Head-to-Head Comparison of Rapid and Automated Antigen Detection Tests for the Diagnosis of SARS-CoV-2 Infection. J Clin Med 2021;10. https://doi.org/10.3390/jcm10020265.

FIND Evaluation of Joysbio (Tianjin) Biotechnology Co., Ltd., SARS-CoV-2 Antigen Rapid Test Kit (Colloidal Gold), External Report, Version 1.0. 2021.

FIND Evaluation of Boditech Medical, Inc. iChroma COVID-19 Ag Test, External Report, Version 1.0. 2021.

FIND Evaluation of Guangzhou Wondfo Biotech Co., Ltd Wondfo 2019-nCoV Antigen Test (Lateral Flow Method), Public Report, Version 1.0. 2021.

Fournier P-E, Zandotti C, Ninove L, Prudent E, Colson P, Gazin C, et al. Contribution of VitaPCR SARS-CoV-2 to the emergency diagnosis of COVID-19. J Clin Virol 2020;133:104682. https://doi.org/10.1016/j.jcv.2020.104682.

Gibani MM, Toumazou C, Sohbati M, Sahoo R, Karvela M, Hon T-K, et al. CovidNudge: diagnostic accuracy of a novel lab-free point-of-care diagnostic for SARS-CoV-2. MedRxiv 2020:2020.08.13.20174193. https://doi.org/10.1101/2020.08.13.20174193.

Kilic A, Hiestand B, Palavecino E. Evaluation of Performance of the BD Veritor SARS-CoV-2 Chromatographic Immunoassay Test in Patients with Symptoms of COVID-19. J Clin Microbiol 2021;59. https://doi.org/10.1128/JCM.00260-21.

Kogoj R, Rus KR, Uršič T. Shortening turnaround time for high-priority patients during the COVID-19 epidemic: evaluation of the Xpert Xpress SARS-CoV-2 test. Slovenian Medical Journal 2020;89:614–25. https://doi.org/10.6016/ZdravVestn.3103.

Korenkov M, Poopalasingam N, Madler M, Vanshylla K, Eggeling R, Wirtz M, et al. Reliable assessment of in vitro SARS-CoV-2 infectivity by a Rapid Antigen Detection Test. MedRxiv 2021:2021.03.30.21254624. https://doi.org/10.1101/2021.03.30.21254624.

Landaas ET, Storm ML, Tollånes MC, Barlinn R, Kran A-MB, Bragstad K, et al. Diagnostic performance of a SARS-CoV-2 rapid antigen test in a large, Norwegian cohort. J Clin Virol 2021;137:104789. https://doi.org/10.1016/j.jcv.2021.104789.

Linares M, Pérez-Tanoira R, Carrero A, Romanyk J, Pérez-García F, Gómez-Herruz P, et al. Panbio antigen rapid test is reliable to diagnose SARS-CoV-2 infection in the first 7 days after the onset of symptoms. J Clin Virol 2020;133:104659. https://doi.org/10.1016/j.jcv.2020.104659.

Marti JLG, Gribschaw J, McCullough M, Mallon A, Acero J, Kinzler A, et al. Differences in detected viral loads guide use of SARS-CoV-2 antigen-detection assays towards symptomatic college students and children. MedRxiv 2021:2021.01.28.21250365. https://doi.org/10.1101/2021.01.28.21250365.

Masiá M, Fernández-González M, Sánchez M, Carvajal M, García JA, Gonzalo-Jiménez N, et al. Nasopharyngeal Panbio COVID-19 Antigen Performed at Point-of-Care Has a High Sensitivity in Symptomatic and Asymptomatic Patients With Higher Risk for Transmission and Older Age. Open Forum Infect Dis 2021;8:ofab059. https://doi.org/10.1093/ofid/ofab059.

Matsuda EM, de Campos IB, de Oliveira IP, Colpas DR, Carmo AMDS, Brígido LF de M. Field evaluation of COVID-19 antigen tests versus RNA based detection: Potential lower sensitivity compensated by immediate results, technical simplicity, and low cost. J Med Virol 2021;93:4405–10. https://doi.org/10.1002/jmv.26985.

McDonald S, Courtney DM, Clark AE, Muthukumar A, Lee F, Balani J, et al. Diagnostic Performance of a Rapid Point-of-care Test for SARS-CoV-2 in an Urban Emergency Department Setting. Acad Emerg Med 2020;27:764–6. https://doi.org/10.1111/acem.14039.

Merino P, Guinea J, Muñoz-Gallego I, González-Donapetry P, Galán JC, Antona N, et al. Multicenter evaluation of the Panbio™ COVID-19 rapid antigen-detection test for the diagnosis of SARS-CoV-2 infection. Clin Microbiol Infect 2021. https://doi.org/10.1016/j.cmi.2021.02.001.

Muhi S, Tayler N, Hoang T, Ballard SA, Graham M, Rojek A, et al. Multi-site assessment of rapid, point-of-care antigen testing for the diagnosis of SARS-CoV-2 infection in a low-prevalence setting: A validation and implementation study. Lancet Reg Health West Pac 2021;9:100115. https://doi.org/10.1016/j.lanwpc.2021.100115.

Nsoga MTN, Kronig I, Rodriguez FJP, Sattonnet-Roche P, Silva DD, Helbling J, et al. Diagnostic accuracy of Panbio™ rapid antigen tests on oropharyngeal swabs for detection of SARS-CoV-2. MedRxiv 2021:2021.01.30.21250314. https://doi.org/10.1101/2021.01.30.21250314.

Peña M, Ampuero M, Garcés C, Gaggero A, García P, Velasquez MS, et al. Performance of SARS-CoV-2 rapid antigen test compared with real-time RT-PCR in asymptomatic individuals. Int J Infect Dis 2021;107:201–4. https://doi.org/10.1016/j.ijid.2021.04.087.

Pilarowski G, Lebel P, Sunshine S, Liu J, Crawford E, Marquez C, et al. Performance Characteristics of a Rapid Severe Acute Respiratory Syndrome Coronavirus 2 Antigen Detection Assay at a Public Plaza Testing Site in San Francisco. J Infect Dis 2021;223:1139–44. https://doi.org/10.1093/infdis/jiaa802.

Ristić M, Nikolić N, Čabarkapa V, Turkulov V, Petrović V. Validation of the STANDARD Q COVID-19 antigen test in Vojvodina, Serbia. PLoS One 2021;16:e0247606. https://doi.org/10.1371/journal.pone.0247606.

Salvagno GL, Gianfilippi G, Bragantini D, Henry BM, Lippi G. Clinical assessment of the Roche SARS-CoV-2 rapid antigen test. Diagnosis (Berl) 2021. https://doi.org/10.1515/dx-2020-0154.

Seitz T, Schindler S, Winkelmeyer P, Zach B, Wenisch C, Zoufaly A, et al. Evaluation of rapid antigen tests based on saliva for the detection of SARS-CoV-2. J Med Virol 2021;93:4161–2. https://doi.org/10.1002/jmv.26983.

Shrestha B, Neupane AK, Pant S, Shrestha A, Bastola A, Rajbhandari B, et al. Sensitivity and Specificity of Lateral Flow Antigen Test Kits for COVID-19 in Asymptomatic Population of Quarantine Centre of Province 3. Kathmandu Univ Med J (KUMJ) 2020;18:36–9.

Torres I, Poujois S, Albert E, Álvarez G, Colomina J, Navarro D. Point-of-care evaluation of a rapid antigen test (CLINITEST® Rapid COVID-19 Antigen Test) for diagnosis of SARS-CoV-2 infection in symptomatic and asymptomatic individuals. J Infect 2021;82:e11–2. https://doi.org/10.1016/j.jinf.2021.02.010.

Turcato G, Zaboli A, Pfeifer N, Ciccariello L, Sibilio S, Tezza G, et al. Clinical application of a rapid antigen test for the detection of SARS-CoV-2 infection in symptomatic and asymptomatic patients evaluated in the emergency department: A preliminary report. J Infect 2021;82:e14–6. https://doi.org/10.1016/j.jinf.2020.12.012.

Wagenhäuser I, Knies K, Rauschenberger V, Eisenmann M, McDonogh M, Petri N, et al. Clinical performance evaluation of SARS-CoV-2 rapid antigen testing in point of care usage in comparison to RT-qPCR. MedRxiv 2021:2021.03.27.21253966. https://doi.org/10.1101/2021.03.27.21253966.

Wang D, He S, Wang X, Yan Y, Liu J, Wu S, et al. Rapid lateral flow immunoassay for the fluorescence detection of SARS-CoV-2 RNA. Nat Biomed Eng 2020;4:1150–8. https://doi.org/10.1038/s41551-020-00655-z.

**Insufficient reported results**

Drain PK, Ampajwala M, Chappel C, Gvozden AB, Hoppers M, Wang M, et al. A Rapid, High-Sensitivity SARS-CoV-2 Nucleocapsid Immunoassay to Aid Diagnosis of Acute COVID-19 at the Point of Care: A Clinical Performance Study. Infect Dis Ther 2021;10:753–61. https://doi.org/10.1007/s40121-021-00413-x.

Kernéis S, Elie C, Fourgeaud J, Choupeaux L, Delarue SM, Alby M-L, et al. Accuracy of antigen and nucleic acid amplification testing on saliva and naopharyngeal samples for detection of SARS-CoV-2 in ambulatory care. MedRxiv 2021:2021.04.08.21255144. https://doi.org/10.1101/2021.04.08.21255144.

Leber W, Lammel O, Monika R-F, Panovska-Griffiths J, Czypionka T. Comparing the Diagnostic Accuracy of Point-of-Care Lateral Flow Antigen Testing for SARS-CoV-2 with RT-PCR in Primary Care (REAP-2). Rochester, NY: Social Science Research Network; 2021. https://doi.org/10.2139/ssrn.3796103.

Peto T, Team UC-19 LFO. COVID-19: Rapid Antigen detection for SARS-CoV-2 by lateral flow assay: a national systematic evaluation for mass-testing. MedRxiv 2021:2021.01.13.21249563. https://doi.org/10.1101/2021.01.13.21249563.

**Ineligible publication type**

Berger U. Schnell-Tests auf Covid-19: Was gibt es da zu diskutieren? Psychother Psychosom Med Psychol 2020;70:308–10.

Boodman C, Lagacé-Wiens P, Bullard J. Diagnostic testing for SARS-CoV-2. CMAJ 2020;192:E713. https://doi.org/10.1503/cmaj.200858.

Buella Parivallal P, Prasanna Kumar MK, Vasanthakumar KC, Thavuroollah FF, Manoharan Y, Muthu S. LAMP Assay: Could it be a Boon for the Molecular Diagnosis of COVID-19? Indian J Clin Biochem 2020:1–2. https://doi.org/10.1007/s12291-020-00915-4.

Cabinet Office and Department of Health and Social Care. Community testing for people without symptoms of coronavirus. GOVUK n.d. https://www.gov.uk/government/publications/community-testing-explainer (accessed June 20, 2021).

Carvalho LF das CES de, Nogueira MS. Optical techniques for fast screening - Towards prevention of the coronavirus COVID-19 outbreak. Photodiagnosis Photodyn Ther 2020;30:101765. https://doi.org/10.1016/j.pdpdt.2020.101765.

Colavita F, Vairo F, Meschi S, Valli MB, Lalle E, Castilletti C, et al. COVID-19 Rapid Antigen Test as Screening Strategy at Points of Entry: Experience in Lazio Region, Central Italy, August-October 2020. Biomolecules 2021;11. https://doi.org/10.3390/biom11030425.

Crozier A, Rajan S, Buchan I, McKee M. Put to the test: use of rapid testing technologies for covid-19. BMJ 2021;372:n208. https://doi.org/10.1136/bmj.n208.

Department for Education and Department of Health and Social Care. Mass asymptomatic testing: schools and colleges. GOVUK n.d. https://www.gov.uk/guidance/asymptomatic-testing-in-schools-and-colleges (accessed June 20, 2021).

Department of Health and Social Care. Assessment and procurement of coronavirus (COVID-19) tests. GOVUK n.d. https://www.gov.uk/government/publications/assessment-and-procurement-of-coronavirus-covid-19-tests (accessed June 20, 2021).

FIND Evaluation of Abbott Panbio COVID-19 Ag Rapid Test Device, External Report, Version 2.1. 2020.

FIND Evaluation of RapiGEN Inc. BIOCREDIT COVID-19 Ag, External Report, Version 2.1. 2020.

FIND Evaluation of SD Biosensor, Inc. STANDARD Q COVID-19 Ag Test, External Report, Version 2.1. 2020.

FIND Evaluation of SD Biosensor, Inc. STANDARD™ F COVID-19 Ag FIA, External Report, Version 2.1. 2020.

FIND Evaluation of Joysbio (Tianjin) Biotechnology Co., Ltd., SARS-CoV-2 Antigen Rapid Test Kit (Colloidal Gold), External Report, Version 1.0. 2021.

FIND Evaluation of Bionote, Inc. NowCheck COVID-19 Ag Test, External Report, Version 1.5. 2021.

FIND Evaluation of NADAL COVID-19 Ag Rapid Test, External Report, Version 1.0. 2021.

García-Díaz G, Montalvo-Varela E, Cano-Pérez E, da Silva Francisco Junior R, Rodriguez-Morales A. Usefulness and limitations of implementing rapid tests for the diagnosis of COVID-19 in Latin America. Infez Med 2020;28:642–4.

Hogan CA, Sahoo MK, Huang C, Garamani N, Stevens B, Zehnder J, et al. Five-minute point-of-care testing for SARS-CoV-2: Not there yet. J Clin Virol 2020;128:104410. https://doi.org/10.1016/j.jcv.2020.104410.

Johnson-León M, Caplan AL, Kenny L, Buchan I, Fesi L, Olhava P, et al. Executive summary: It's wrong not to test: The case for universal, frequent rapid COVID-19 testing. EClinicalMedicine 2021;33:100759. https://doi.org/10.1016/j.eclinm.2021.100759.

Liu M, Arora RK, Krajden M. Rapid antigen tests for SARS-CoV-2. CMAJ 2021;193:E447. https://doi.org/10.1503/cmaj.202827.

McDermott A. Inner Workings: Researchers race to develop in-home testing for COVID-19, a potential game changer. Proc Natl Acad Sci U S A 2020;117:25956–9. https://doi.org/10.1073/pnas.2019062117.

McDermott JH, Stoddard D, Ellingford JM, Gokhale D, Reynard C, Black G, et al. Utilizing point-of-care diagnostics to minimize nosocomial infection in the 2019 novel coronavirus (SARS-CoV-2) pandemic. QJM 2020;113:851–3. https://doi.org/10.1093/qjmed/hcaa185.

Mertz L. CRISPR Tech Behind Super-Sensitive, Smartphone COVID Test. IEEE Pulse 2021;12:8–11. https://doi.org/10.1109/MPULS.2021.3066716.

Mina MJ, Andersen KG. COVID-19 testing: One size does not fit all. Science 2021;371:126–7. https://doi.org/10.1126/science.abe9187.

Mina MJ, Peto TE, García-Fiñana M, Semple MG, Buchan IE. Clarifying the evidence on SARS-CoV-2 antigen rapid tests in public health responses to COVID-19. Lancet 2021;397:1425–7. https://doi.org/10.1016/S0140-6736(21)00425-6.

Nimmo C, Agbetile J, Bhowmik A, Capocci S, Rajakulasingam RK. Implementing rapid diagnostics for COVID-19. Lancet Respir Med 2021;9:e7. https://doi.org/10.1016/S2213-2600(20)30526-9.

Patriquin G, LeBlanc JJ. SARS-CoV-2 sensitivity limbo - How low can we go? Int J Infect Dis 2021;103:23–4. https://doi.org/10.1016/j.ijid.2020.11.138.

Peeling RW, Olliaro P. Rolling out COVID-19 antigen rapid diagnostic tests: the time is now. Lancet Infect Dis 2021. https://doi.org/10.1016/S1473-3099(21)00152-3.

Royal College of Pathologists. Briefing : LAMP assays for SARS-CoV-2. Royal College of Pathologists 2020.

Sheridan C. Fast, portable tests come online to curb coronavirus pandemic. Nat Biotechnol 2020;38:515–8. https://doi.org/10.1038/d41587-020-00010-2.

Shuren J, Stenzel T. Covid-19 Molecular Diagnostic Testing - Lessons Learned. N Engl J Med 2020;383:e97. https://doi.org/10.1056/NEJMp2023830.

Stovitz SD. In suspected SARS-CoV-2, rapid antigen detection tests had 67% to 73% sensitivity and 98% to 100% specificity. Ann Intern Med 2021;174:JC56. https://doi.org/10.7326/ACPJ202105180-056.

Thampi N, Sander B, Science M. Preventing the introduction of SARS-CoV-2 into school settings. CMAJ 2021;193:E24–5. https://doi.org/10.1503/cmaj.202568.

Thornton HV, Khalid T, Hay AD. Point-of-care testing for respiratory infections during and after COVID-19. Br J Gen Pract 2020;70:574–5. https://doi.org/10.3399/bjgp20X713561.

Ujiie M, Ohmagari N, Inoue H. Testing for COVID-19 at travel clinics in Japan. J Travel Med 2020;27. https://doi.org/10.1093/jtm/taaa107.

Usherwood T, Zhang L, Tripathi A. The Path Forward for COVID-19 Diagnostics. Mol Diagn Ther 2020;24:637–9. https://doi.org/10.1007/s40291-020-00492-5.

Walbert H. Vergessen Sie die Schnellteste auf Coronavirus! MMW Fortschritte Der Medizin 2020;162. https://doi.org/10.1007/s15006-020-0337-7.

Weiss G, Bellmann-Weiler R. Rapid antigen testing and non-infectious shedding of SARS-Cov2. Infection 2021. https://doi.org/10.1007/s15010-020-01570-w.

Yang T, Wang Y-C, Shen C-F, Cheng C-M. Point-of-Care RNA-Based Diagnostic Device for COVID-19. Diagnostics (Basel) 2020;10. https://doi.org/10.3390/diagnostics10030165.

**Multiple publication**

Albert E, Torres I, Bueno F, Huntley D, Molla E, Fernández-Fuentes MÁ, et al. Field evaluation of a rapid antigen test (Panbio™ COVID-19 Ag Rapid Test Device) for the diagnosis of COVID-19 in primary healthcare centers. MedRxiv 2020:2020.10.16.20213850. https://doi.org/10.1101/2020.10.16.20213850.

Basso D, Aita A, Padoan A, Cosma C, Navaglia F, Moz S, et al. Salivary SARS-CoV-2 antigen rapid detection: a prospective cohort study. MedRxiv 2020:2020.12.24.20248825. https://doi.org/10.1101/2020.12.24.20248825.

Bokelmann L, Nickel O, Maricic T, Pääbo S, Meyer M, Borte S, et al. Rapid, reliable, and cheap point-of-care bulk testing for SARS-CoV-2 by combining hybridization capture with improved colorimetric LAMP (Cap-iLAMP). MedRxiv 2020:2020.08.04.20168617. https://doi.org/10.1101/2020.08.04.20168617.

Caruana G, Croxatto A, Kampouri E, Kritikos A, Opota O, Foerster M, et al. ImplemeNting SARS-CoV-2 Rapid antigen testing in the Emergency wArd of a Swiss univErsity hospital: the INCREASE study. MedRxiv 2021:2021.02.10.21250915. https://doi.org/10.1101/2021.02.10.21250915.

Dinnes J., Deeks J.J., Adriano A., Berhane S., Davenport C., Dittrich S., et al. Rapid, point-of-care antigen and molecular-based tests for diagnosis of SARS-CoV-2 infection. Cochrane Database Syst Rev 2020;2020:CD013705. https://doi.org/10.1002/14651858.CD013705.

Drain PK, Ampajwala M, Chappel C, Gvozden AB, Hoppers M, Wang M, et al. A rapid, high-sensitivity SARS-CoV-2 nucleocapsid immunoassay to aid diagnosis of acute COVID-19 at the point of care. MedRxiv 2020:2020.12.11.20238410. https://doi.org/10.1101/2020.12.11.20238410.

Gremmels H, Winkel BMF, Schuurman R, Rosingh A, Rigter NAM, Rodriguez O, et al. Real-life validation of the Panbio COVID-19 Antigen Rapid Test (Abbott) in community-dwelling subjects with symptoms of potential SARS-CoV-2 infection. MedRxiv 2020:2020.10.16.20214189. https://doi.org/10.1101/2020.10.16.20214189.

Iglὀi Z, Velzing J, Beek J van, Vijver D van de, Aron G, Ensing R, et al. Clinical evaluation of the Roche/SD Biosensor rapid antigen test with symptomatic, non-hospitalized patients in a municipal health service drive-through testing site. MedRxiv 2020:2020.11.18.20234104. https://doi.org/10.1101/2020.11.18.20234104.

Lephart PR, Bachman M, LeBar W, McClellan S, Barron K, Schroeder L, et al. Comparative study of four SARS-CoV-2 Nucleic Acid Amplification Test (NAAT) platforms demonstrates that ID NOW performance is impaired substantially by patient and specimen type. BioRxiv 2020:2020.06.04.135616. https://doi.org/10.1101/2020.06.04.135616.

Lippi G, Gianfillipi G, Bragantini D, Henry B, Salvagno GL. Clinical Assessment of the Roche SARS-CoV-2 Rapid Antigen Test. SSRN 2020. https://doi.org/10.2139/ssrn.3746520.

Masiá M, Fernández-González M, Sánchez M, Carvajal M, García JA, Gonzalo N, et al. Nasopharyngeal Panbio COVID-19 antigen performed at point-of-care has a high sensitivity in symptomatic and asymptomatic patients with higher risk for transmission and older age. MedRxiv 2020:2020.11.16.20230003. https://doi.org/10.1101/2020.11.16.20230003.

Merino-Amador P, Guinea J, Muñoz-Gallego I, González-Donapetry P, Galán J-C, Antona N, et al. Multicenter evaluation of the Panbio™ COVID-19 Rapid Antigen-Detection Test for the diagnosis of SARS-CoV-2 infection. MedRxiv 2020:2020.11.18.20230375. https://doi.org/10.1101/2020.11.18.20230375.

Muhi S, Tayler N, Hoang T, Ballard S, Graham M, Rojek A, et al. Multi-Site Assessment of Rapid, Point-of-Care Antigen Testing for the Diagnosis of SARS-CoV-2 Infection in a Low-Prevalence Setting: A Validation and Implementation Stud. SSRN 2021. https://doi.org/10.2139/ssrn.3750715.

Olearo F, Nörz D, Heinrich F, Sutter JP, Rödel K, Schultze A, et al. Handling and accuracy of four rapid antigen tests for the diagnosis of SARS-CoV-2 compared to RT-qPCR. MedRxiv 2020:2020.12.05.20244673. https://doi.org/10.1101/2020.12.05.20244673.

Peña M, Ampuero M, Garcés C, Gaggero A, García P, Velasquez MS, et al. Performance of SARS-CoV-2 rapid antigen test compared with real-time RT-PCR in asymptomatic individuals. MedRxiv 2021:2021.02.12.21251643. https://doi.org/10.1101/2021.02.12.21251643.

Pollock NR, Jacobs JR, Tran K, Cranston A, Smith S, Kane CO, et al. Performance and Implementation Evaluation of the Abbott BinaxNOW Rapid Antigen Test in a High-throughput Drive-through Community Testing Site in Massachusetts. MedRxiv 2021:2021.01.09.21249499. https://doi.org/10.1101/2021.01.09.21249499.

Shah MM, Salvatore PP, Ford L, Kamitani E, Whaley MJ, Mitchell K, et al. Performance of Repeat BinaxNOW SARS-CoV-2 Antigen Testing in a Community Setting, Wisconsin, November-December 2020. MedRxiv 2021:2021.04.05.21254834. https://doi.org/10.1101/2021.04.05.21254834.

Torres I, Poujois S, Albert E, Colomina J, Navarro D. Real-life evaluation of a rapid antigen test (Panbio™ COVID-19 Ag Rapid Test Device) for SARS-CoV-2 detection in asymptomatic close contacts of COVID-19 patients. MedRxiv 2020:2020.12.01.20241562. https://doi.org/10.1101/2020.12.01.20241562.

Wei S, Kohl E, Djandji A, Morgan S, Whittier S, Mansukhani M, et al. Field-deployable, rapid diagnostic testing of saliva samples for SARS-CoV-2. MedRxiv 2020:2020.06.13.20129841. https://doi.org/10.1101/2020.06.13.20129841.

Zwirglmaier K, Weyh M, Krüger C, Ehmann R, Müller K, Wölfel R, et al. Rapid detection of SARS-CoV-2 by pulse-controlled amplification (PCA). MedRxiv 2020:2020.07.29.20154104. https://doi.org/10.1101/2020.07.29.20154104.
